# Gut microbiome-derived metabolic remodeling and the butyrate–IL-18 inflammatory axis after transcatheter aortic valve implantation

**DOI:** 10.64898/2026.08.30.26361742

**Authors:** Caroline Chong-Nguyen, Cyril Ferro, Bahtiyar Yilmaz, Daijiro Tomii, Camille Dupuy, Lydie Nadal-Desbarats, Pamela Nicholson, Aparna Pandey, Thomas Pilgrim, Yvonne Döring

**Affiliations:** Department of Angiology, Inselspital, Bern University Hospital, University of Bern, Bern, Switzerland; MIDHOS: Metabolism I Inflammation I Digital Health I Osteology, Faculty of Medicine, University of Bern, Bern, Switzerland; Department of Cardiology, Biel/Bienne Hospital, Switzerland; Department of BioMedical Research, University of Bern, Switzerland; Department of Cardiology, Inselspital, Bern University Hospital, University of Bern, Freiburgstrasse 20, Bern, CH-3010, Switzerland; Department of Visceral Surgery and Medicine, Bern University Hospital, University of Bern, 3010, Bern, Switzerland; Maurice Müller Laboratories, Department for Biomedical Research, University of Bern, 3008, Bern, Switzerland; Plateforme de Métabolomique et d’Analyses Chimiques, US61 ASB, Université de Tours, CHRU Tours, Inserm, Tours, France; Université de Tours, INSERM, Imaging Brain & Neuropsychiatry iBraiN U1253, 37032, Tours, France; Next Generation Sequencing Platform, University of Bern, Bremgartenstrasse 109a, 3012 Bern, Switzerland; Interfaculty Bioinformatics Unit and Swiss Institute of Bioinformatics, University of Bern, Bern, Switzerland; Institute for Cardiovascular Prevention, Ludwig-Maximilians-Universität, Munich, Germany

**Keywords:** transcatheter aortic valve implantation (TAVI), aortic stenosis (AS), gut microbiome, short-chain fatty acids (SCFA), butyrate, interleukin-18 (IL-18)

## Abstract

**Background:** Severe aortic stenosis is associated with systemic and splanchnic hemodynamic disturbances that may alter gut microbial metabolism and host inflammatory responses.

**Objectives:** We aimed to determine whether TAVI remodels the gut microbiome-derived metabolome and whether post-procedural SCFA dynamics are associated with the inflammatory cytokine response.

**Methods:** We conducted a prospective paired single-center study of patients undergoing elective TAVI at Bern University Hospital. Stool and blood samples were collected before and three months after the procedure. Gut microbial composition was profiled by full-length 16S rRNA sequencing, circulating short-chain fatty acids (SCFAs) by targeted metabolomics, and inflammatory mediators by multiplex cytokine analysis, and integrated with hemodynamic and clinical data.

**Results:** Forty patients were enrolled. Following TAVI, microbial richness declined without significant restructuring of overall community composition. In contrast, circulating SCFA profiles were significantly remodeled, driven by selective reductions in butyrate and isovalerate. A greater decline in circulating butyrate was inversely associated with IL-18 elevation (ρ⍰=⍰−0.668, p⍰<⍰0.001, n⍰=⍰36), independent of aortic valve calcification burden, hemodynamic improvement, and cardiovascular medications. Baseline isovalerate was nominally associated with 1-month adjudicated adverse events (AUC⍰0.77; exploratory).

**Conclusions:** TAVI is associated with selective changes in gut microbiome-derived metabolic output rather than broad alterations in microbial community structure. Declining circulating butyrate identifies a gut–metabolite–immune axis linked to IL-18 dynamics and represents a potential biomarker of inflammatory recovery following valve intervention.

**Condensed Abstract:** Whether TAVI remodels the gut microbiome-derived metabolome and whether short-chain fatty acid (SCFA) dynamics couple to the post-procedural inflammatory response has not been examined. In 40 patients undergoing elective TAVI, paired stool and serum samples collected before and three months after the procedure were analyzed by full-length 16S rRNA sequencing, targeted SCFA metabolomics, and multiplex cytokine profiling. Gut microbial richness declined modestly without significant restructuring of community composition. Circulating butyrate and isovalerate were selectively reduced after TAVI. Post-procedural butyrate decline was specifically and inversely associated with IL-18 elevation (ρ⍰=⍰−0.668, p⍰<⍰0.001), independent of hemodynamic response, calcification burden, and cardiovascular risk profile. These findings identify a gut-metabolite-immune axis that may represent a modifiable biological dimension of post-procedural recovery after cardiac valve intervention.

## INTRODUCTION

Gut microbial composition is associated with cardiovascular risk across disease phenotypes including atherosclerosis, coronary artery disease, and heart failure. (1–3) Gut-derived metabolites are key mechanistic intermediates in this relationship: trimethylamine N-oxide (TMAO), produced by microbial fermentation of dietary phosphatidylcholine and carnitine, is an established mediator of atherogenesis.(4) Among the most abundant gut-derived circulating metabolites are the short-chain fatty acids (SCFAs), acetate, propionate, and butyrate, generated by anaerobic fermentation of dietary fiber by colonic bacteria. SCFAs regulate intestinal barrier integrity, modulate immune cell differentiation, and attenuate systemic inflammatory signaling through receptor-mediated and epigenetic mechanisms.8/30/2026 2:32:00 AM.

Severe aortic stenosis reduces cardiac output and impairs splanchnic perfusion, compromising intestinal mucosal integrity through the same hemodynamic axis that underlies Heyde syndrome.(5,6) Transcatheter aortic valve implantation (TAVI) corrects the hemodynamic obstruction but does not uniformly resolve its systemic sequelae: residual systemic inflammation is common in the post-procedural period,(7) and the procedural context itself (anesthesia, iodinated contrast, and peri-procedural antibiotic exposure) may represent an additional stressor to which the gut ecosystem responds, as suggested by observations of gut mucosal hypoperfusion during cardiac procedures.(8) Whether TAVI remodels the gut microbiome-derived metabolome, and whether individual variability in post-procedural SCFA dynamics is coupled to the post-procedural inflammatory response, has not previously been examined.

We conducted a prospective longitudinal study of patients undergoing TAVI, in whom paired 16S rRNA sequencing, targeted circulating SCFA metabolomics, and a cytokine panel were performed before and after the procedure, to test the hypothesis that TAVI is associated with remodeling of the gut microbiome-derived metabolome coupled to differences in inflammatory cytokine trajectory.

## METHODS

All methodology details concerning metabolomics, metagenomics, cytokine panel assessment and adjudicated adverse events are described in the **Supplementary Data Methods document**.

### Study Design and Patient Population

This was a prospective observational, single-center cohort study conducted at Bern University Hospital, Switzerland. Patients with significant aortic valve disease scheduled for elective transcatheter aortic valve implantation (TAVI) were enrolled and studied in a paired longitudinal design, with assessments before TAVI (PRE) and at 3-month after TAVI (POST).

The study was nested within the Swiss-TAVI registry framework, with stool and blood biospecimens collected at both time points together with standardized clinical, echocardiographic, laboratory, and imaging data.

Eligible patients were adults aged ≥18 years who were hospitalized for TAVI or for pre-interventional assessment before TAVI for significant aortic valve disease, including severe calcific aortic stenosis or combined severe aortic stenosis and aortic regurgitation requiring valve intervention. Severe aortic stenosis was defined according to hemodynamic criteria, including high-gradient severe aortic stenosis, classical low-flow low-gradient aortic stenosis, or paradoxical low-gradient aortic stenosis, as specified in the study protocol.

Key exclusion criteria were treatments known to interfere with intestinal microbiota composition within the preceding 3 months, including systemic or local corticosteroids, antibiotics, antiretroviral therapy, bile acid sequestrants, HIV-targeted antiretroviral therapies, and selective serotonin reuptake inhibitors. Additional exclusion criteria included history of cholecystectomy, chronic liver disease, inflammatory bowel disease, failure to fast on the day of blood sampling, valve-in-valve TAVI, left ventricular ejection fraction <20%, emergency intervention, rheumatic aortic stenosis, and infective endocarditis. Study was approved by the local Ethics Committee, and all participants provided written informed consent.

### TAVI Procedure

All participants were scheduled for elective TAVI after the indication for transcatheter treatment of severe aortic stenosis had been reviewed and approved by the interdisciplinary Heart Team. Procedural characteristics, including access route, prosthesis type, valve sizing approach, and anesthesia strategy, were determined according to the clinical and anatomical requirements of the individual patient. Access was categorized as transfemoral or alternative access. Peri-procedural antibiotic prophylaxis was standardized across the cohort: all patients received Amoxicillin 2.2 g and Clavulanic acid intravenously as a single dose immediately before the intervention.

### Biospecimen Collection

Stool and blood samples were collected before TAVI and at the 3-month post-TAVI follow-up visit. Baseline stool samples could be obtained within 4 weeks before the intervention, whereas follow-up stool samples were collected at the routine 3-month visit to minimize interference from acute procedural stress and peri-procedural antibiotic exposure. Participants collected stool using a dedicated stool sampling kit. Samples were processed by homogenization, aliquoted, and stored at −80°C until analysis.

Blood samples were obtained at the pre-interventional clinical assessment, during hospitalization before TAVI, or at the follow-up visit, preferably in conjunction with routine blood sampling to avoid additional patient burden. Patients were instructed to fast for at least 6 hours before blood collection (typically from midnight); compliance was 100%. Two 5-mL blood tubes were collected per patient per timepoint. Blood was centrifuged by the study team; serum was isolated, aliquoted, and stored at −80°C until analysis. Cytokine analytes are stable at this storage temperature.(9) Paired pre-procedural and post-procedural samples from the same patient were processed and analyzed within the same analytical batch to minimize inter-batch variability. Samples were thawed once for analysis. No aortic valve tissue was obtained, consistent with the transcatheter nature of the procedure.

### Circulating Short-Chain Fatty Acid Metabolomics

Circulating short-chain fatty acids were quantified in serum using targeted mass-spectrometry-based metabolomics. The SCFA panel comprised acetate (C2), propionate (C3), butyrate (C4), isobutyrate (branched C4), isovalerate (branched C5), and valerate (C5). Metabolite assays were performed according to standardized and validated chromatographic mass-spectrometry methods.

### Cytokine Panel

Systemic inflammatory mediators were quantified in serum using multiplex electrochemiluminescence immunoassays (V-PLEX, Meso Scale Discovery [MSD], Rockville, MD, USA) according to the manufacturer’s instructions.

The cytokine panel comprised IL-18, IFN-γ, IL-17A(10), IL-1β (11), TNF-α (12), IL-5, IL-17B, and IL-13. Prior to downstream analyses, analytes were evaluated for detectability and analytical performance. IL-13 was excluded because 72.9% of measurements were below the lower limit of quantification.

### Statistical Analysis

All analyses were performed using Python (numpy, scipy, statsmodels). Continuous variables are expressed as median and interquartile range (IQR); categorical variables as counts and percentages. Given the paired study design, pre- to post-procedural changes were assessed using paired Wilcoxon signed-rank tests as the primary test, complemented by paired t-tests. This approach was applied to alpha diversity metrics (Chao1, observed ASVs, Shannon entropy, Simpson index), individual circulating metabolite concentrations, individual cytokine concentrations, and functional guild scores. Where multiple comparisons were performed within a pre-specified family of tests, p-values were adjusted for the false discovery rate using the Benjamini-Hochberg (BH) method; a q-value < 0.05 was the confirmatory significance threshold; an adjusted p-value < 0.25 was considered a suggestive signal warranting evaluation in a future prospective cohort. Correction was applied separately within each pre-specified family of tests; no cross-family adjustment was performed.

For multivariate pre-to-post comparisons, microbial community composition was assessed by Bray-Curtis dissimilarity and tested by PERMANOVA in two configurations: unpaired (group as fixed factor) and paired (stratified permutation by patient, the primary test). The global SCFA metabolome was assessed by paired PERMANOVA on squared Euclidean distances of z-scored log_2_-transformed concentrations, complemented by a Hotelling’s T^2^ sign-permutation test on delta concentration vectors; both analyses used 9,999 permutations. The cytokine panel was assessed by the same paired PERMANOVA approach. Ordination was visualized by principal coordinate analysis (PCoA). Within-patient distances across time points were compared to between-patient distances to characterize the relative contribution of inter-individual variability versus procedural effect.

Bivariate associations between continuous variables were assessed using Spearman’s rank correlation coefficient (ρ), computed as Pearson’s r on rank-transformed data. Two-tailed permutation p-values were derived from 5000 random permutations and 95% confidence intervals by the bootstrap percentile method (B=5000 resamples). This framework was applied to delta SCFA × delta cytokine associations, delta SCFA × hemodynamic parameter correlations, and cross-omics correlations between functional guild scores and circulating metabolite changes.

### Exploratory Analyses

To explore the potential clinical relevance of post-procedural SCFA remodeling, patients were classified into a SCFA responder phenotype. For each SCFA that changed significantly at the cohort level in the pre-post analysis, individual delta values were standardized (z-scored; mean 0, SD 1 across the cohort). A composite z-score was computed as the mean of the standardized delta values across the significant SCFAs; patients with a composite z-score > 0 were classified as SCFA Responders and those ≤ 0 as Non-Responders. This classification was defined a priori based on the pre-post comparison results and was not optimized for the outcome. Post-procedural adverse events were ascertained from the study database at the 3-month follow-up visit; patients with incomplete follow-up were excluded from this analysis. Between-group adverse event rates were compared using Fisher’s exact test (two-sided).

### Adverse Event Ascertainment and Classification

Adverse events were ascertained across four timepoints: periprocedural (Day 0), 1-month (Day 1–35), 3-month, and 1-year. Data sources were: (i) the SwissTAVI registry discharge and 30-day follow-up forms for periprocedural and 1-month events; (ii) REDCap 3-month follow-up visit data; (iii) the SwissTAVI 1-year follow-up registry sheet for conduction variables and cumulative atrial fibrillation.

## RESULTS

### Clinical Cohort

A total of 40 TAVI patients (median age 81 years [IQR 78.5-85.3], 52.5% females) were enrolled between March 2024 and July 2024. Complete paired datasets were available for circulating metabolomics in 39 patients, microbiome profiling in 38 patients, and the primary SCFA–cytokine correlation cohort in 36 patients (**Figure 1**). Baseline characteristics are summarized in **Table 1**. Most patients had established cardiovascular risk profiles: hypertension in 33 (82.5%), dyslipidemia in 21 (52.5%), diabetes mellitus in 9 (22.5%), and prior coronary artery disease in 8 (20.0%). Echocardiographic parameters confirmed severe aortic stenosis with a median mean transvalvular gradient of 40.0 mmHg [32.3–47.0], aortic valve area of 0.80 cm^2^ [0.70–0.98], and preserved left ventricular ejection fraction (60.5% [54.8–66.8]). Aortic valve calcification as assessed by computed tomography was severe in the majority of patients (29/33, 87.9% Agatston grade 3), with a median aortic valve calcium score of 2819 Agatston units [2129-3342] (n=38).

**Figure 1.**
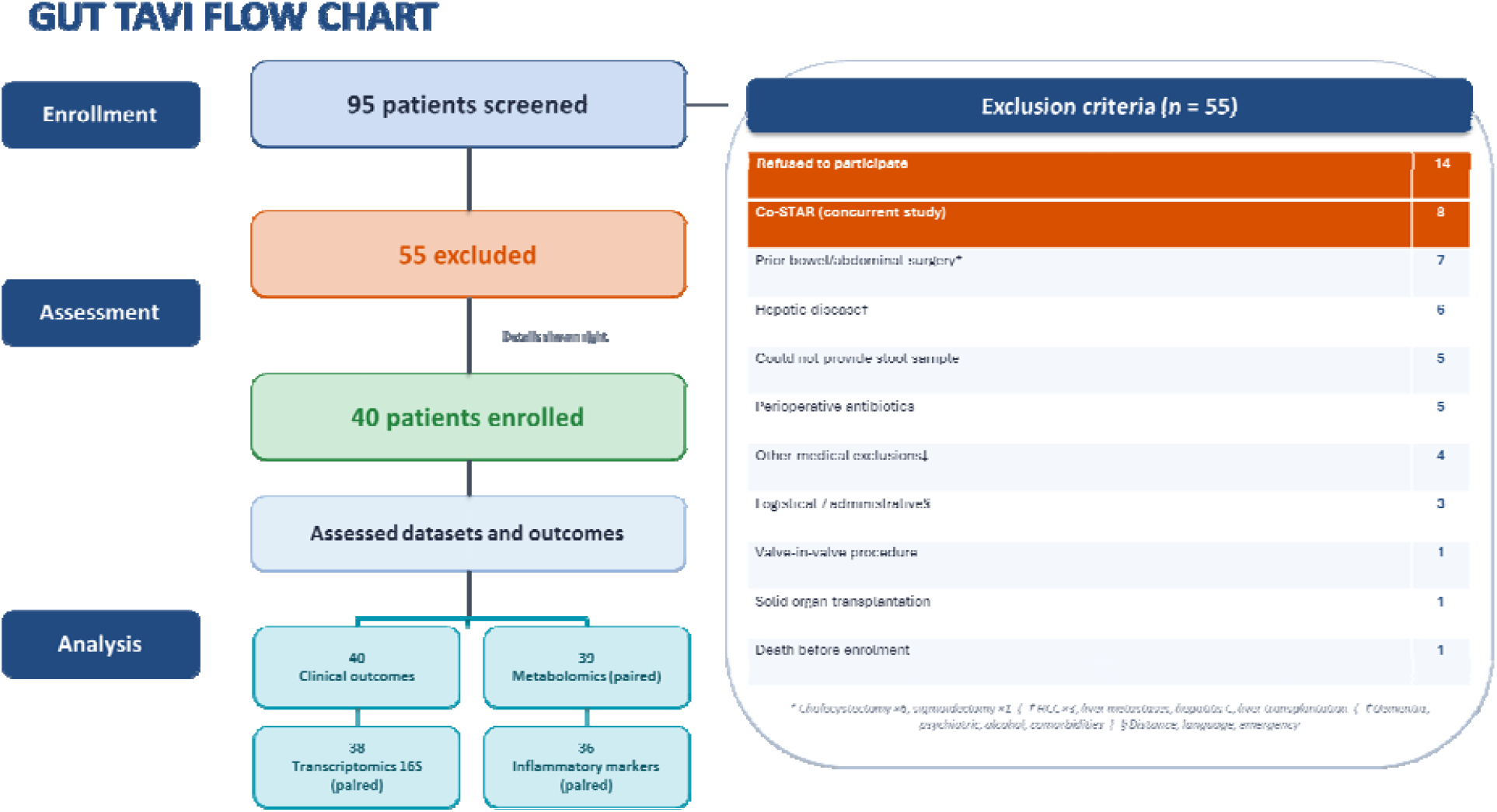
Flow chart.

Clinical outcomes across four follow-up windows are summarized in **Table 2**. Periprocedural adverse events, predominantly conduction disturbances and early structural valve complications, were observed in a minority of patients. At 1 month, the total number of events remained low, with conduction-related events predominating. The pattern at 3 months and 1 year was consistent with the expected post-TAVI trajectory in an elderly high-risk cohort, encompassing conduction system complications, hemodynamic leaflet thrombosis, new-onset atrial fibrillation, and adjudicated serious adverse events. A consistent directional pattern was observed across timepoints, without a single category of event dominating the follow-up period.

### Gut Microbial Diversity and Community Structure Following TAVI

Three months after TAVI, gut microbial richness was modestly but significantly reduced. Both ASV richness declined compared with baseline (**Figure 2A; Figure S1; Table S1**), whereas Shannon and Simpson diversity also declined numerically, although these changes were not statistically significant, indicating preservation of overall community diversity despite the loss of low-abundance taxa (**Figure S2**). Individual responses were heterogeneous, with some patients exhibiting stable or increased richness; nevertheless, the overall cohort demonstrated a consistent reduction in richness (**Figure 2B**).

**Figure 2A.**
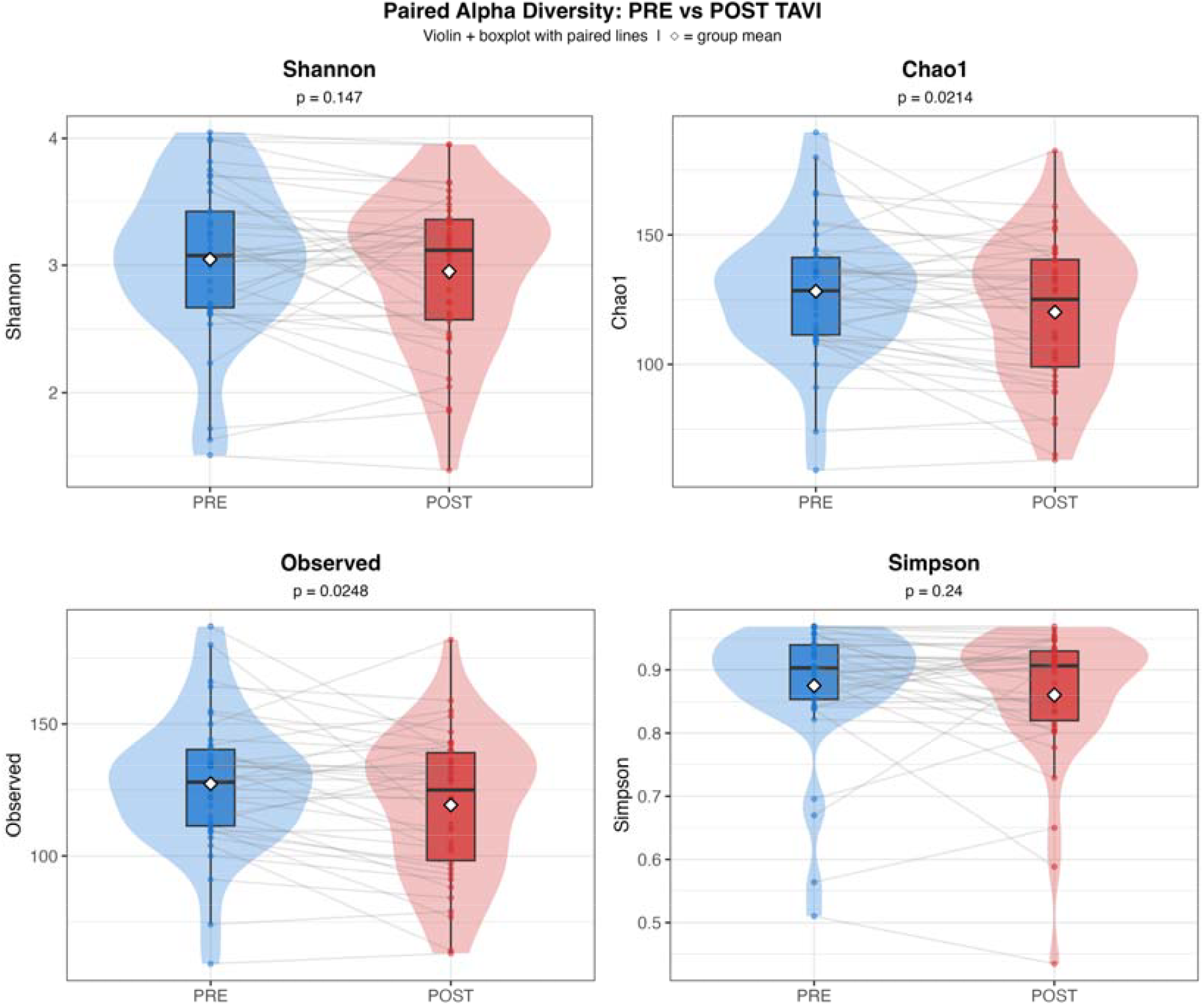
Paired alpha diversity before and after TAVI. Paired violin and boxplots compare alpha-diversity metrics before and after TAVI. Observed species counts and Chao1 richness declined after TAVI, while Shannon and Simpson diversity remained unchanged, indicating a reduction in microbial richness without a global loss of diversity.

**Figure 2B.**
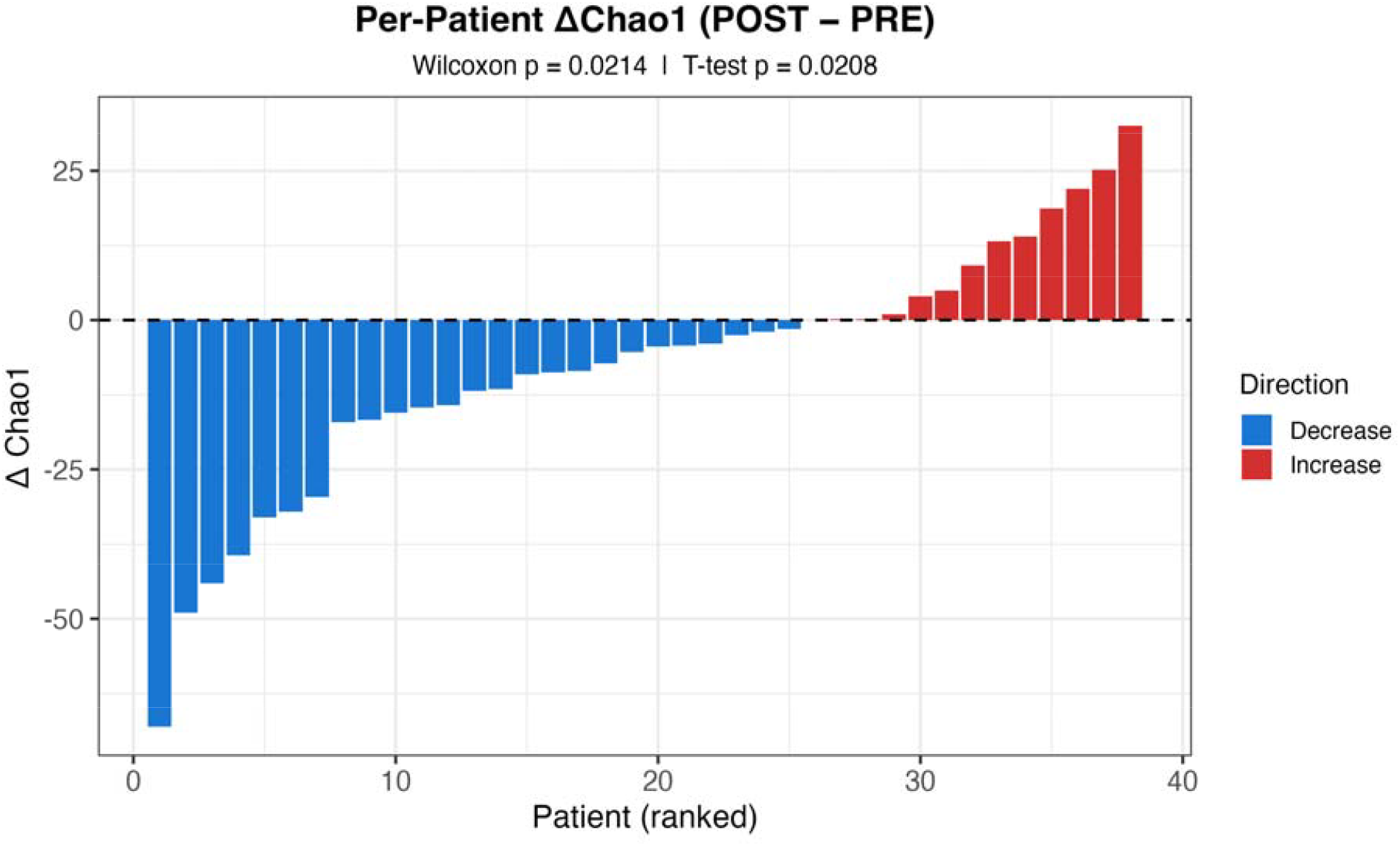
Per-patient Chao1 richness changes after TAVI. The ranked per-patient Chao1 change plot shows heterogeneous individual trajectories: some patients had stable or increasing richness, but the cohort-level pattern showed decreased Chao1 and observed species counts after TAVI.

**Figure 2C.**
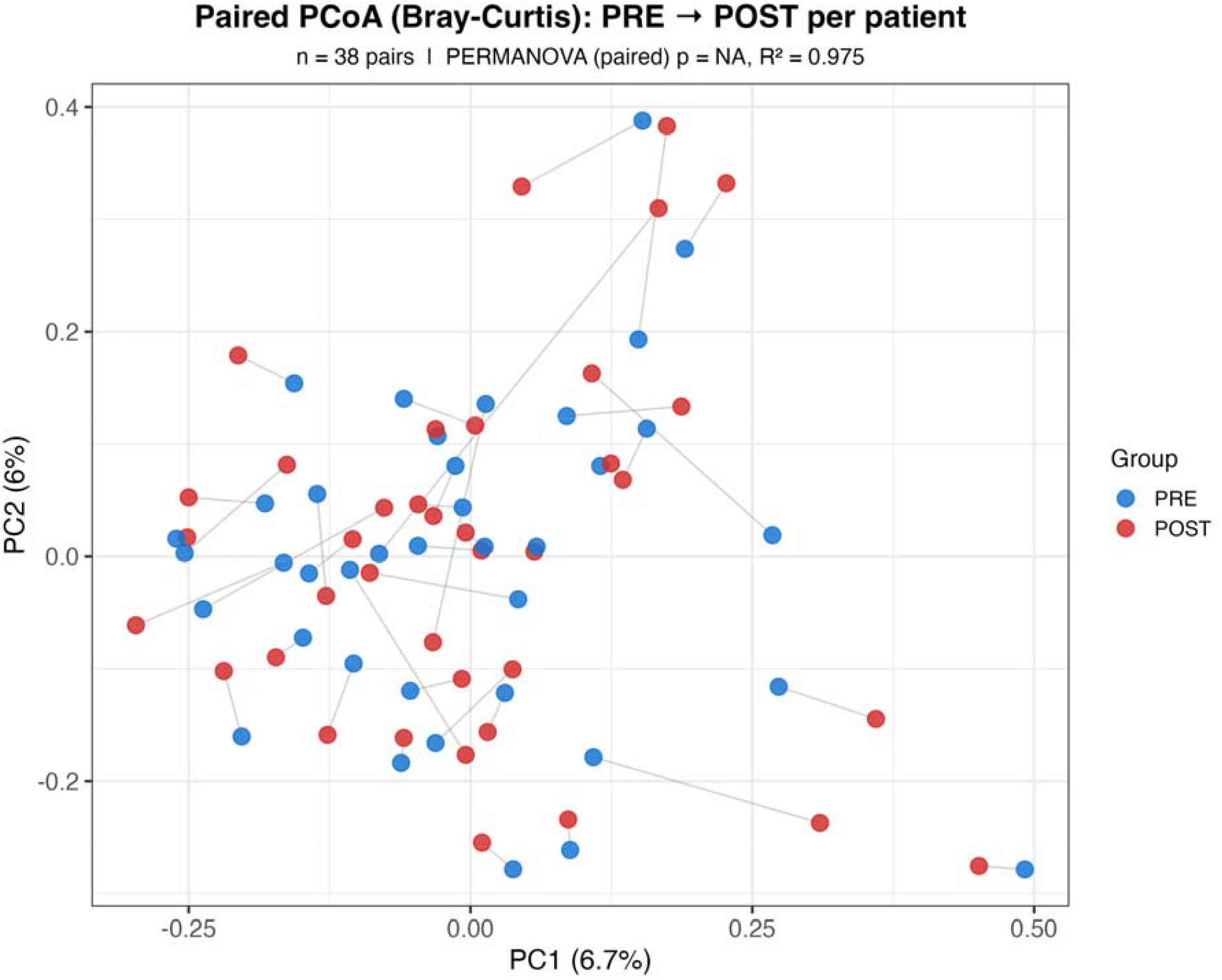
Paired Bray-Curtis PCoA before and after TAVI. The plot visualizes PRE-to-POST microbiome community shifts within patients. Overall community composition was not significantly restructured after TAVI, and within-patient PRE-POST distances were smaller than between-patient distances, supporting preservation of overall microbiome structure.

Despite these changes, global microbial community composition remained largely preserved after TAVI. Bray-Curtis principal coordinate analysis showed substantial overlap between pre- and post-procedural samples, and paired PERMANOVA detected no significant shift in overall community structure (R^2^ = 0.025, p = 0.692; **Figure 2C; Figure S3**). Consistent with this finding, within-patient microbiome dissimilarities between baseline and follow-up were considerably smaller than differences observed between individuals (**Figure S4**), indicating that inter-individual variation remained the dominant determinant of microbial community structure following TAVI.

### Circulating SCFA Remodeling Following TAVI

TAVI was associated with a significant shift in the circulating SCFA metabolome at the class level (paired PERMANOVA pseudo-F = 2.97, R^2^ = 0.038, p = 0.014; Hotelling’s T^2^ p < 0.001; **Figure 3A; Table S2**). The R^2^ of 0.038 indicates that the PRE-to-POST transition accounted for approximately 4% of total SCFA metabolome variance, consistent with a modest cohort-level effect in the context of pronounced inter-individual variability. At the individual metabolite level, butyric acid (q = 0.020) and isovaleric acid (q = 0.012) were the only two SCFA to decline significantly after correction for multiple testing (**Figure 3B; Table S3**). Acetate, propionate, and isobutyrate did not change significantly. Microbial functional guild scores showed heterogeneous pre-to-post trajectories without significant cohort-level change after BH-FDR correction (**Figure S5**); the 16S-derived butyrate-producing guild score was not correlated with circulating butyrate (**Figure S6**). No SCFA delta correlated with delta aortic mean gradient, delta maximum gradient, or delta Vmax after BH-FDR correction (**Figure S7**).

**Figure 3A.**
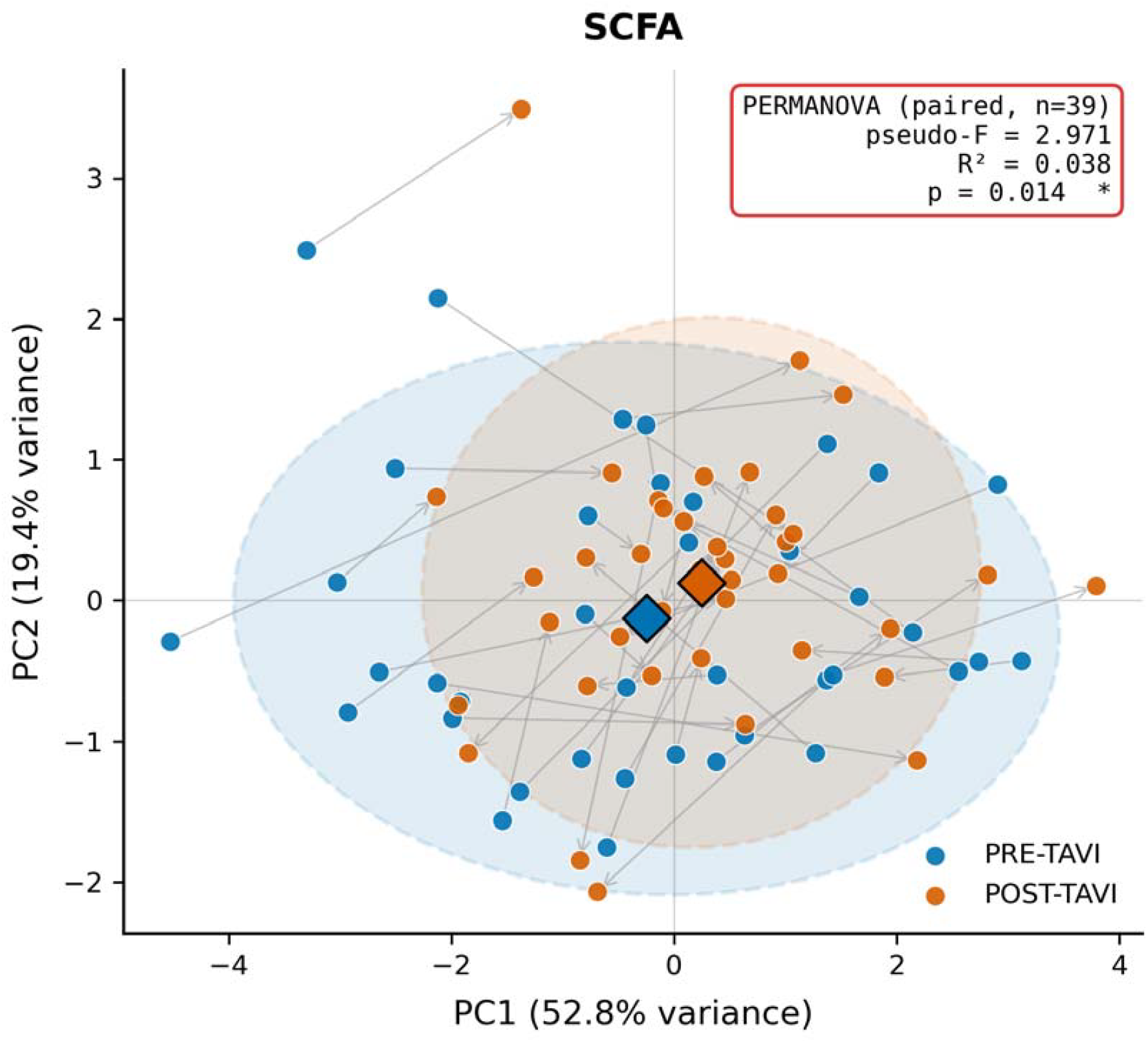
Circulating SCFA metabolome remodeling after TAVI. PCoA of circulating SCFA profiles shows a significant class-level shift after TAVI (paired PERMANOVA pseudo-F = 2.97, R^2^ = 0.038, p = 0.014; Hotelling’s T^2^ p < 0.001), indicating remodeling of the SCFA metabolome.

**Figure 3B.**
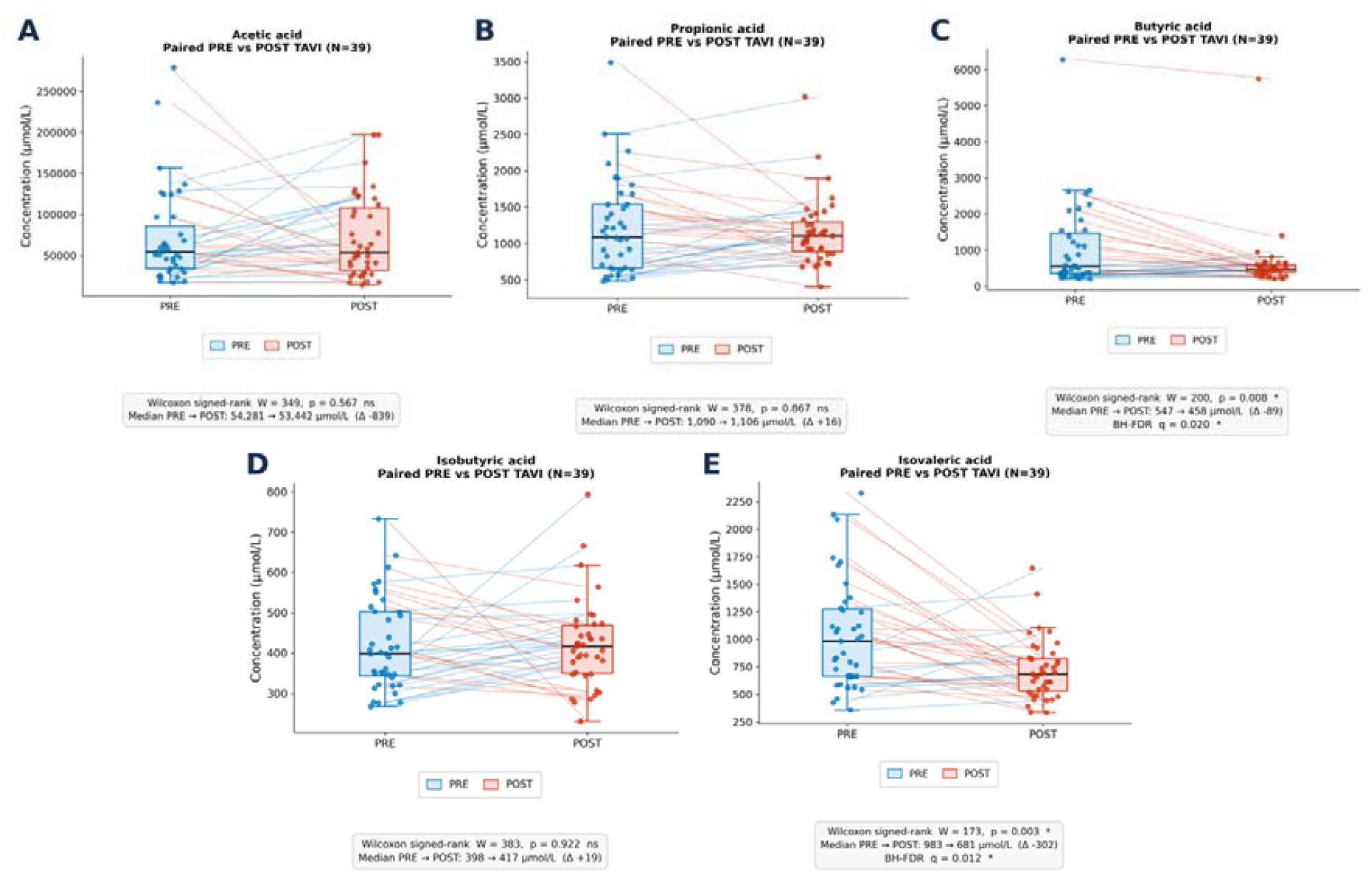
Paired PRE-to-POST concentrations. Paired plasma concentrations at baseline (PRE) and 3-month follow-up (POST) for the five quantifiable short-chain fatty acids: (A) acetic acid, (B) propionic acid, (C) butyric acid, (D) isobutyric acid, (E) isovaleric acid. Valeric acid was excluded a priori (84.6% of values below the lower limit of quantification). Boxes show median and interquartile range; lines connect PRE→POST values within the same patient (n = 39 paired samples). Comparisons by paired Wilcoxon signed-rank test (two-sided); p-values corrected for multiple testing using the Benjamini-Hochberg method within the SCFA class (5 tests).

### Post-TAVI Reduction in Circulating Butyrate Is Specifically Associated with Elevation of IL-18

The cytokine panel was not significantly restructured at the class level following TAVI (paired PERMANOVA R^2^ = 0.011, p = 0.630, n = 35; Table S4). We examined Spearman correlations between delta-SCFA and delta-cytokine values across the paired cohort (n = 36). No SCFA other than butyrate showed a consistent directional relationship with any cytokine after correction for multiple testing (**Figure 4A**). Post-TAVI reduction in circulating butyrate was inversely correlated with ΔIL-18 (ρ = −0.668, 95% CI [−0.820, −0.412], p < 0.0002; **Figure 4B**). Among seven cytokines tested with BH-FDR correction, only ΔIL-18 (q < 0.001) and ΔIFN-γ (ρ = −0.631, q < 0.001; **Figure 4C; Figure S8**) were significantly associated with ΔButyrate; IL-1β, TNF-α, IL-17A, IL-17B, and IL-5 were not. At the cohort level, no cytokine changed significantly following TAVI after correction for multiple testing (**Table S4; Figure S9**); IL-18 concentrations were stable (median Δ = −51.0 pg/mL, p = 0.706, q = 0.983), and a nominal decrease in IL-1β did not survive FDR correction (p = 0.018, q = 0.123). IL-1β was not associated with ΔButyrate.

**Figure 4A.**
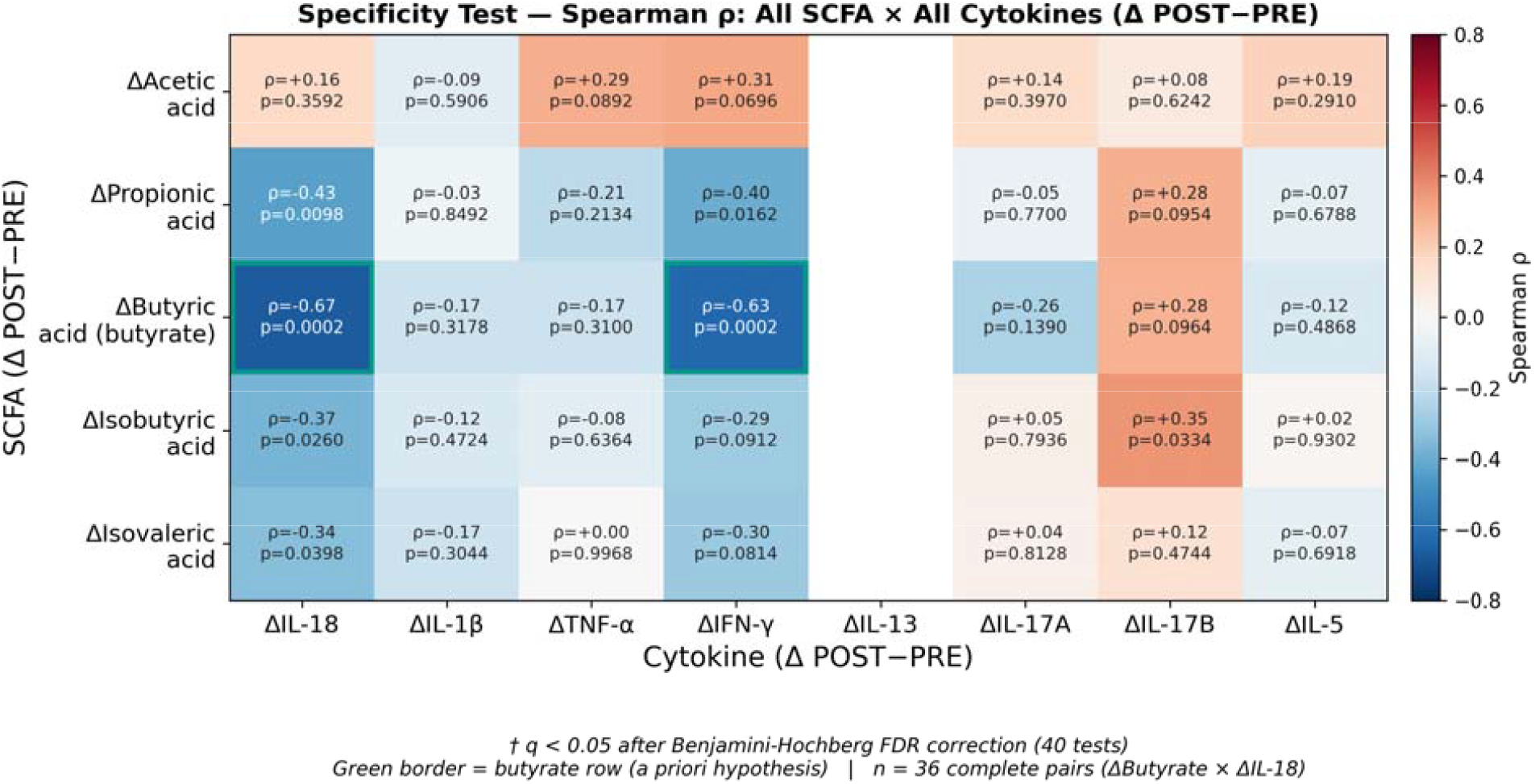
Delta-SCFA and delta-cytokine association matrix. The heatmap summarizes Spearman correlations between changes in SCFAs and cytokines after TAVI. No SCFA other than butyrate showed a consistent directional relationship with cytokines after multiple-testing correction, highlighting the specificity of the butyrate-cytokine signal.

**Figure 4B.**
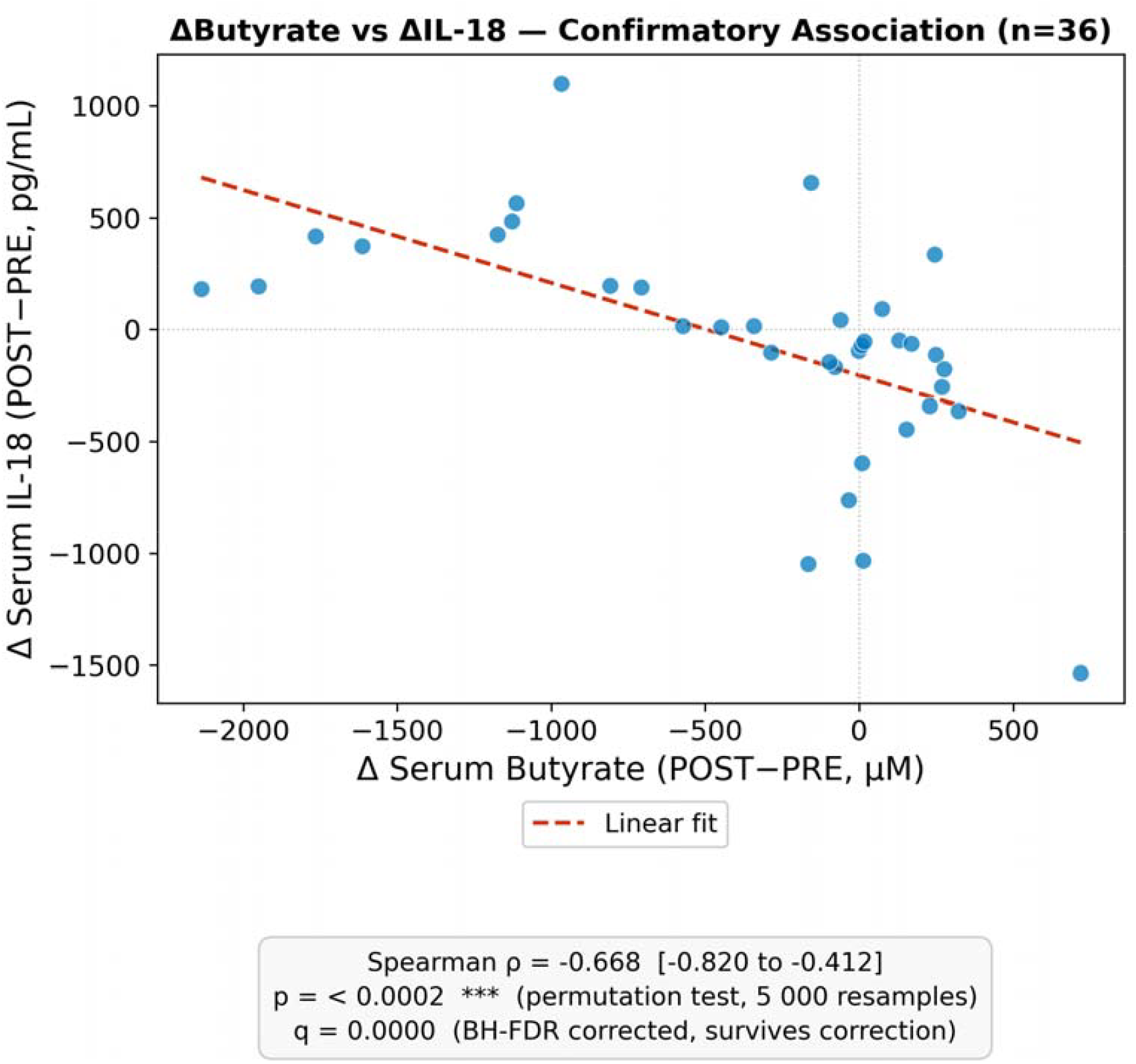
Association between butyrate decline and IL-18 elevation. The scatter plot shows the confirmatory inverse association between change in serum butyrate and changes in IL-18: greater post-TAVI butyrate decline was associated with IL-18 elevation (ρ = -0.668, 95% CI -0.820 to -0.412, p < 0.0002).

**Figure 4C.**
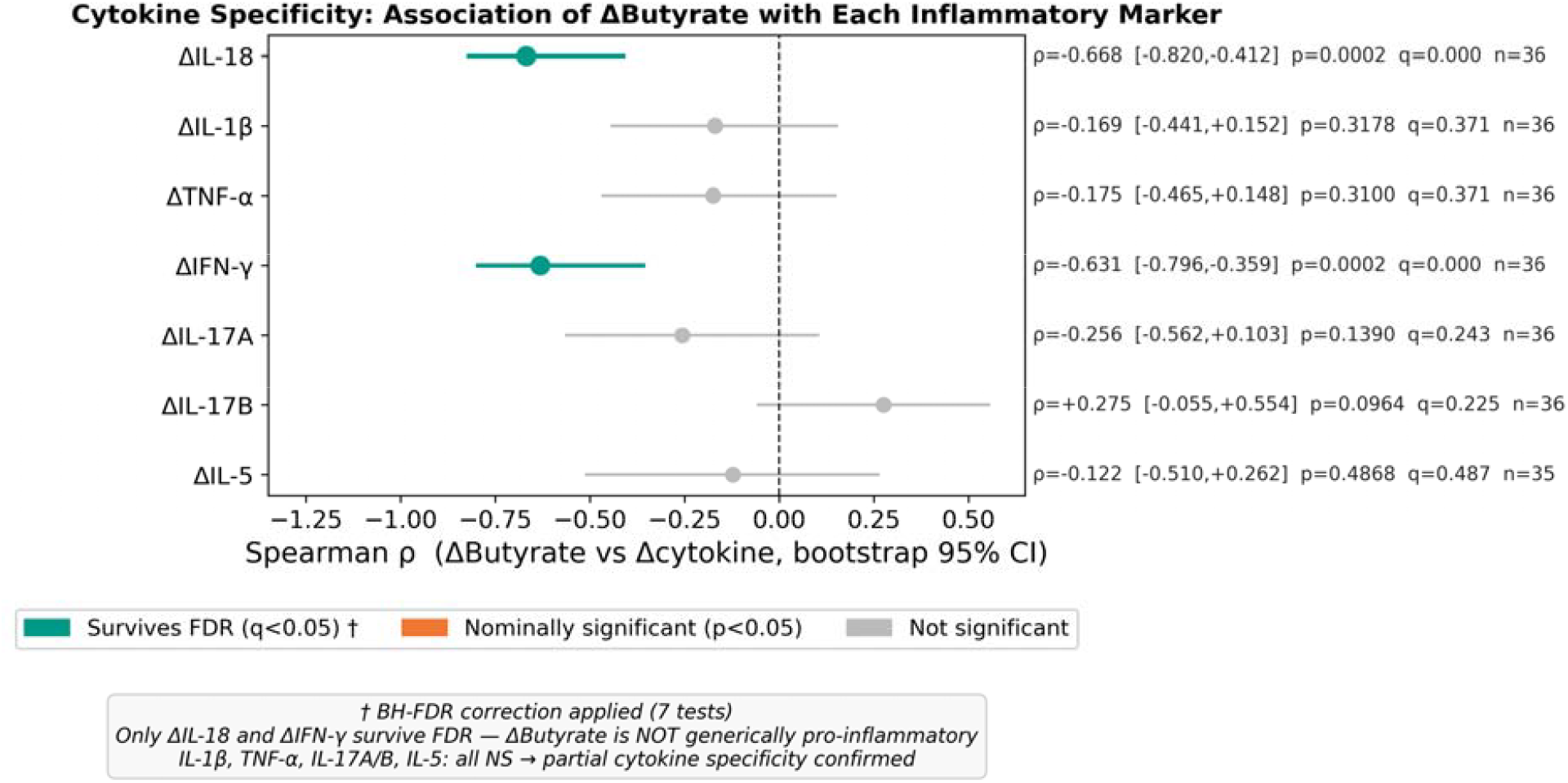
Cytokine-specificity of the delta-butyrate association. Among seven cytokines tested with BH-FDR correction, only ΔIL-18 and ΔIFN-γ were significantly associated with ΔButyrate; IL-1β, TNF-α, IL-17A, IL-17B, and IL-5 were not.

**Figure 4D.**
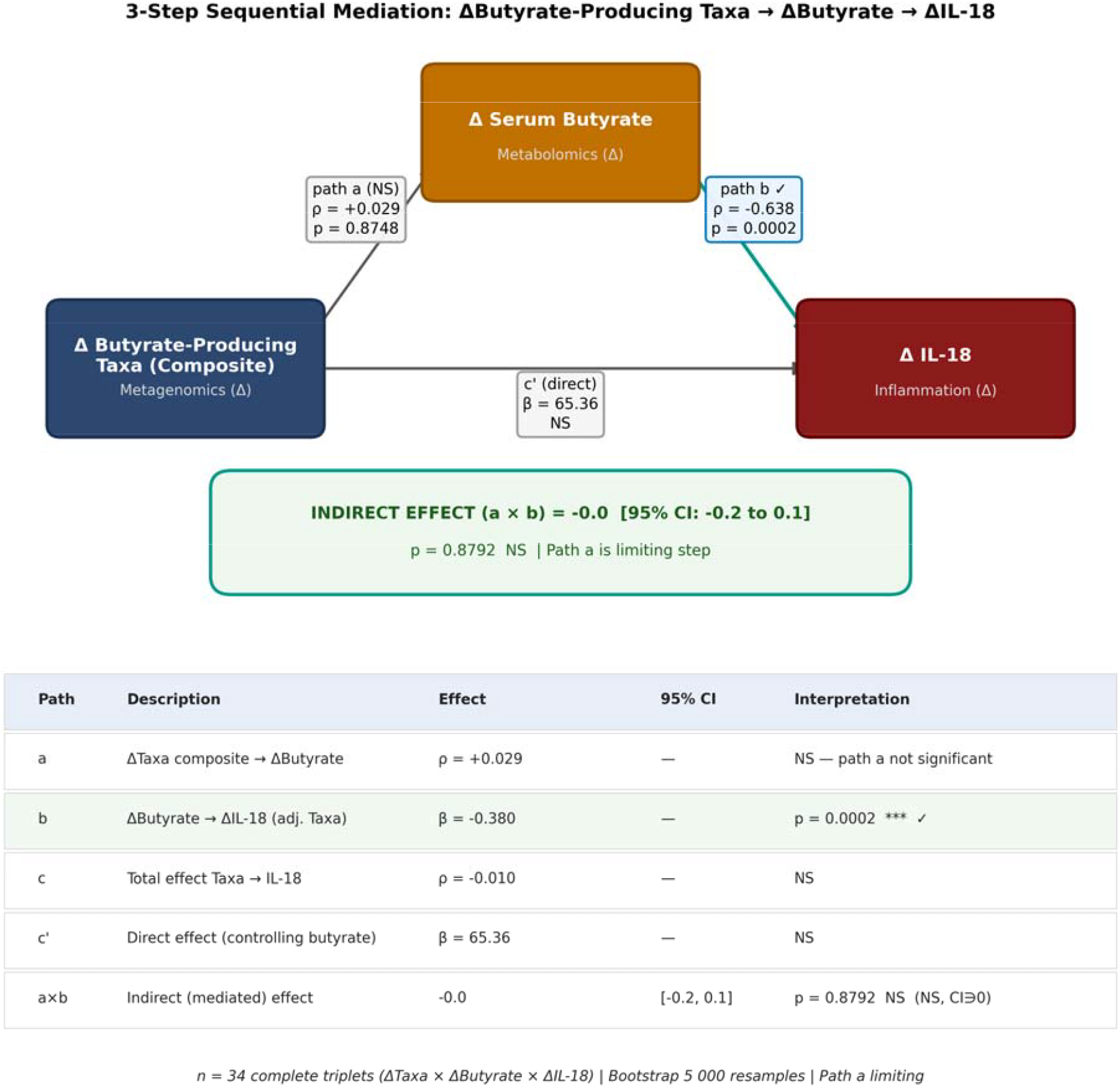
Sequential association model for butyrate-producing taxa, butyrate, and IL-18. The model tests whether changes in butyrate-producing taxa statistically link taxon-level change to the ΔButyrate–ΔIL-18 association. The butyrate-to-IL-18 path remained significant, but no butyrate-producing taxon independently predicted butyrate change, indicating that the taxon-level pathway did not account for the butyrate–IL-18 association. Because all variables were derived from the same PRE-to-POST interval, this model cannot establish temporal precedence.

In a sequential association model (n = 34), the butyrate→IL-18 association (ρ = −0.649, p = 0.0002) was significant, while no butyrate-producing taxon independently predicted butyrate change (**Figure 4D**). When kynurenine was included as a parallel covariate, ΔButyrate remained inversely associated with ΔIL-18 (partial ρ = −0.591, p = 0.0002); ΔKynurenine was not independently associated with ΔIL-18 after adjustment for ΔButyrate (**Figure S10**).

### The ΔButyrate–ΔIL-18 axis is independent of baseline patient characteristics and procedural hemodynamic response

The inverse association between post-TAVI butyrate reduction and IL-18 elevation was not explained by the degree of hemodynamic improvement, aortic valve calcification burden, peri-procedural hematological stress, or baseline demographics and cardiovascular medications. Partial Spearman correlations after adjustment for aortic valve calcification burden (n = 34), peri-procedural hemoglobin decline, cardiovascular medications (statin, ACEI, aspirin individually and jointly), and demographic variables (BMI, age, sex individually and jointly) ranged from −0.602 to −0.740, all p ≤ 0.0004 (**Figures S11-14; Table S5**). ΔButyrate was not associated with peri-procedural changes in hemoglobin or creatinine (**Figure S15**). Across all pre-specified sensitivity analyses, the signal was preserved, and in several instances strengthened, indicating that the ΔButyrate–ΔIL-18 relationship may work independently of procedural success and patient profile. Whether this independence reflects an intrinsic property of the gut–immune axis or inter-patient variability in microbiome-derived metabolic resilience cannot be determined from the available data.

### Post-TAVI SCFA Trajectory and Adverse Events

To explore the potential clinical relevance of post-TAVI SCFA remodeling, patients were stratified according to their composite SCFA trajectory. Individuals demonstrating an overall increase in butyrate and isovalerate (the two SCFAs that declined significantly at the cohort level) were classified as SCFA Responders (composite standardized z-score above zero; n=19), and those with stable or declining concentrations as Non-Responders (n=20). Responders and Non-Responders were clinically balanced at baseline: no demographic, comorbidity, procedural-risk, echocardiographic, or ECG variable differed significantly between groups (**Table S6**), indicating that the SCFA classification reflects post-procedural trajectory rather than pre-existing clinical risk.

Because the Responder/Non-Responder classification requires the 3-month post-TAVI SCFA sample, it cannot be applied to 1-month events, which precede this measurement. At 1 month, six adjudicated inflammatory and conduction events occurred in the cohort (**Table 2**). Among baseline serum SCFAs (all measured before the procedure), isovalerate was the only metabolite with a nominal association with 1-month adjudicated event status (Mann-Whitney U, p=0.037): patients who experienced a 1-month event had numerically higher baseline isovalerate concentrations than those who did not (**Figure S16**). Receiver operating characteristic analysis yielded an AUC of 0.77 (95% bootstrap CI 0.53-0.94; Youden threshold 1,263 nM; sensitivity 0.83, specificity 0.82; n=6 events; **Figure S17**). Given only 6 events and the number of baseline predictors tested, this association requires prospective validation. No SCFA measurement at 1 month was obtained in this study, precluding change-score analyses; all isovalerate analyses use baseline (pre-procedural) concentrations exclusively. In a sensitivity analysis excluding the sole structural valve deterioration (SVD) case from the 1-month event composite (5 events, n=40), the AUC for baseline isovalerate was materially unchanged (0.74, 95% bootstrap CI 0.46–0.94), indicating the association is not driven by the SVD classification.

At 3 months, inflammatory events (hemodynamic leaflet thrombosis and complete atrioventricular block) occurred numerically less frequently in Responders than Non-Responders (1/19 [5.3%] vs 4/20 [20.0%]; p=0.34; **Figure S18**), consistent with the directional pattern of the original 3-month adverse event comparison previously reported. At 1 year (N=37; two patients without 1-year follow-up excluded), Responders continued to show numerically fewer adjudicated serious adverse events and conduction disturbances across all composites, though no comparison reached statistical significance (**Figure 5; Table 2)**. Across both post-discharge Responder-stratified timepoints (3 months and 1 year), the direction of effect was consistently unfavorable in Non-Responders. All adverse event comparisons are exploratory, no multiplicity correction was applied, and no causal inference is drawn.

**Figure 5.**
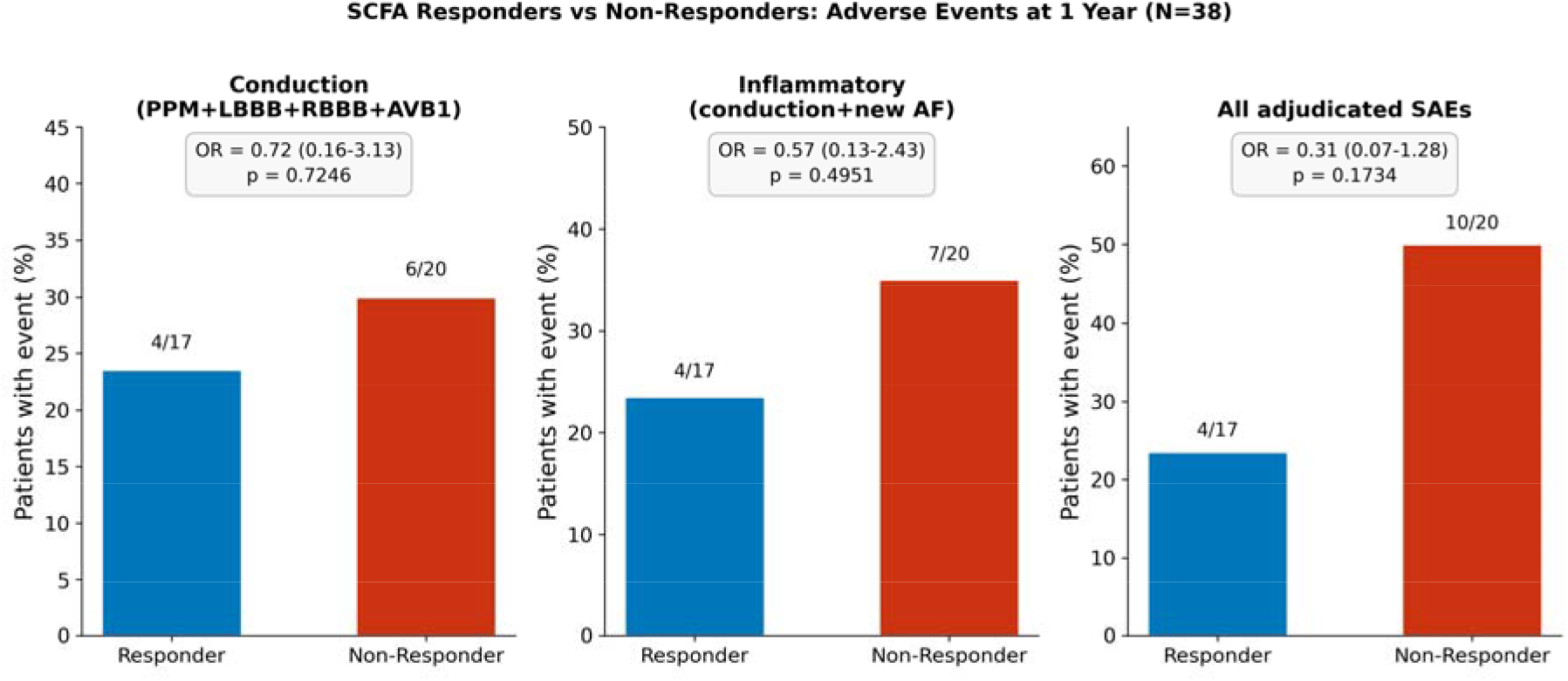
Adverse event rates at 1-year follow-up by SCFA Responder status. Adverse event rates at 1-year follow-up in SCFA Responders (n=17) and Non-Responders (n=20). SCFA Responders were defined as patients with a composite standardised z-score above zero for the pre-to-post TAVI change in butyrate and isovalerate; Non-Responders had a composite z-score at or below zero. Two patients without 1-year follow-up data and one patient without a post-TAVI SCFA sample were excluded (total N=37). Three composites are shown. Conduction composite: new permanent pacemaker implantation, left bundle branch block, right bundle branch block, or first-degree atrioventricular block present at 1-year follow-up. Inflammatory composite: conduction composite plus new-onset atrial fibrillation. All-cause SAE composite: any adjudicated serious adverse event at 1-year follow-up. p-values from Fisher’s exact test, two-sided. Odds ratios and 95% confidence intervals computed with Haldane–Anscombe continuity correction (+0.5 to all cells) where any cell contained zero observations; p-values from the uncorrected exact test. All comparisons are exploratory; no multiplicity correction was applied. *SCFA, short-chain fatty acid; TAVI, transcatheter aortic valve implantation; PPM, permanent pacemaker; LBBB, left bundle branch block; RBBB, right bundle branch block; AVB1, first-degree atrioventricular block; AF, atrial fibrillation; SAE, serious adverse event; OR, odds ratio; CI, confidence interval*.

## DISCUSSION

In this prospective paired study of patients undergoing TAVI, we report three principal findings. First, TAVI was associated with contraction of gut microbial richness without restructuring of overall community composition, indicating selective pressure against rare taxa in the absence of global dysbiosis. Second, circulating SCFAs were significantly remodeled as a class, with selective reduction in butyrate and isovalerate while acetate, propionate, and isobutyrate were unchanged. Third, and most centrally, individual post-procedural butyrate decline was specifically and inversely coupled to IL-18 elevation in a relationship independent of hemodynamic response, aortic valve calcification burden, peri-procedural stress, and cardiovascular medication profile. Taken together, these findings identify a gut–metabolite–immune axis that operates through inter-individual variation in circulating butyrate dynamics, rather than generalized gut dysbiosis, and is consistent with butyrate-associated modulation of inflammasome-linked inflammatory biology following cardiovascular intervention.

### The observed SCFA–IL-18 coupling is consistent with a suppressive inflammasome model

The most central finding of this study was a specific inverse association between individual post-procedural butyrate decline and IL-18 elevation, in the absence of a cohort-level IL-18 shift. The specificity of this coupling, absent at the population level but present across individual trajectories, is consistent with the possibility that the gut–immune relationship in this context operates through inter-individual variation in circulating butyrate dynamics rather than a generalized post-procedural inflammatory response. The present study did not, however, directly measure NLRP3 activity, caspase-1 cleavage, or the ratio of pro-to mature IL-18, and the mechanistic interpretation of this association therefore remains speculative.

The association between butyrate decline and IL-18 elevation is consistent with experimental evidence linking butyrate to regulation of the NLRP3 inflammasome, the primary processing machinery for mature IL-18. IL-18 is not constitutively secreted: its bioactive form requires NLRP3 inflammasome assembly, caspase-1 activation, and enzymatic cleavage of biologically inert pro-IL-18.(13,14) In experimental models, butyrate has been associated with suppression of NLRP3 inflammasome activation through HDAC inhibition and GPR109A receptor signaling.(15,16) Whether reduced butyrate availability after TAVI attenuates this suppressive signal and thereby contributes to the observed IL-18 elevation cannot, however, be determined from the present data. A recent multi-omics study provides contextual support: gut-derived butyric acid modulates CAVD pathogenesis through butyrylation-mediated inhibition of osteogenic differentiation in valvular interstitial cells, with serum butyrate inversely correlating with calcification severity.(17) These findings are nonetheless consistent with the hypothesis that butyrate depletion may shape the inflammatory environment in aortic valve disease through NLRP3-dependent pathways, a question that requires direct experimental testing.

A secondary association with IFN-γ (ρ = −0.631, q < 0.001) suggests that butyrate remodeling may influence additional immune pathways beyond the inflammasome axis.(18,19)

### Functional remodeling occurs despite preservation of microbiome structure

A notable finding of this study is the dissociation between microbiome structure and metabolic function following TAVI. Although overall community composition remained remarkably stable, circulating SCFAs underwent significant remodeling, with selective reductions in butyrate and isovalerate. This observation suggests that preservation of microbial structure does not necessarily imply preservation of microbiome-derived metabolic activity. In elderly patients, whose microbiomes are typically characterized by substantial inter-individual variability and relative temporal stability, biologically relevant changes may occur at the functional level without major shifts in taxonomic composition.(20) More broadly, these observations align with emerging evidence that microbial ecosystems can maintain structural stability through functional redundancy while still exhibiting substantial variability in metabolic activity, particularly at the level of short-chain fatty acid production.(21) The absence of association between circulating butyrate trajectories and 16S-derived butyrate-producing guild scores further supports this interpretation and highlights a recognized limitation of amplicon-based approaches: microbiome-derived metabolic activity is determined by strain-level gene content and metabolic activity that cannot be resolved through taxonomic profiling alone (22). Together, these findings suggest that metabolite-level measurements provide information that is complementary to, and potentially more sensitive than, community composition when assessing gut ecosystem responses to cardiovascular interventions.

The accompanying reduction in richness may indicate selective loss of low-abundance taxa that contribute disproportionately to ecosystem function (23,24). While the specific microbial drivers of the observed SCFA changes could not be identified in this dataset, the findings are consistent with the concept that functional resilience and taxonomic stability represent distinct properties of the gut ecosystem.

### Recovery of gut microbiome-derived metabolic activity may drive long-term butyrate trajectories

The mechanism underlying the cohort-level butyrate decline remains uncertain. Peri-procedural antibiotic prophylaxis, all patients received co-amoxiclav 2.2 g at induction, may have contributed to an initial ecosystem perturbation, but single-dose beta-lactam effects on butyrate-producing Firmicutes are transient (24–48 hours) and are unlikely to account for the inter-individual variability in butyrate trajectory observed at 3 months.(25) A more plausible interpretation of the 3-month assessment is that the observed variability reflects differences in recovery of gut microbiome-derived metabolic activity following the procedural episode, rather than the direct acute effects of antibiotic exposure or hospitalization. Post-procedural nutritional intake, physical recovery trajectory, frailty status, and restoration of intestinal perfusion, all of which vary substantially across TAVI patients and are plausible determinants of gut ecosystem recovery, were not formally captured in the present study and warrant prospective investigation in future cohorts.

### The gut–immune axis may contribute to heterogeneity of post-TAVI recovery

Post-procedural systemic inflammation after TAVI is heterogeneous: two patients with identical procedural success and equivalent residual valve gradients can diverge substantially in their inflammatory recovery. Individual variability in circulating butyrate dynamics, independent of hemodynamic correction, calcification burden, peri-procedural stress, and cardiovascular medication profile, constitutes a distinct biological dimension of this response, consistent with the recognition from the CANTOS trial that inflammasome-derived cytokines are mechanistically relevant to cardiovascular outcomes and amenable to modulation.(26) Prior work has demonstrated that TAVI modifies circulating gut-derived metabolites, with pre-procedural indoxyl sulfate independently predicting MACE at approximately 400 days; the present study extends this evidence to the SCFA metabolome and identifies coupling between SCFA dynamics and an inflammasome-derived cytokine, a dimension not captured by uremic toxin profiling and not previously reported after any cardiac valvular intervention.(27,28)

### SCFA response profiles may enable biological stratification of patients

The adverse event categories observed to occur numerically more frequently in Non-Responders, specifically hemodynamic leaflet thrombosis and conduction system disturbances including complete atrioventricular block and new-onset atrial fibrillation, are not incidental to the biology described above. Both are established inflammatory sequelae of the peri-procedural immune response to TAVI and both implicate the NLRP3 inflammasome axis.(29) Independent experimental evidence for this mechanistic link comes from the Co-STAR randomized trial, which demonstrated that peri-procedural colchicine, an agent that suppresses NLRP3 inflammasome assembly and downstream IL-1β-mediated cascades, reduced new-onset arrhythmias requiring pacemaker implantation, new-onset atrial fibrillation, and subclinical leaflet thrombosis compared with placebo in elderly patients undergoing TAVI. Although the Co-STAR trial was terminated early due to an excess stroke rate in the colchicine group, it establishes the principle that pharmacological attenuation of peri-procedural NLRP3 activity is sufficient to reduce precisely the categories of complication that were directionally enriched in Non-Responders in the present study.(30) In this framework, persistent circulating SCFA depletion after TAVI may represent an endogenous analogue of impaired anti-inflammatory signaling: failure to recover butyrate production after the procedural episode maintains a state of relative NLRP3 permissibility, in which the peri-procedural inflammatory milieu remains conducive to leaflet thrombosis and conduction system injury. Because the post-procedural SCFA sample and 3-month adverse event ascertainment are contemporaneous, the direction of this association cannot be confirmed, and reverse causation (an adverse event altering subsequent SCFA levels through hospitalization, altered nutritional intake, or secondary inflammation) remains equally plausible.

Responders and Non-Responders were clinically indistinguishable at baseline across all measured covariates, supporting the interpretation that post-TAVI SCFA trajectory captures a dimension of post-procedural recovery that is independent of pre-existing clinical risk factors. The absence of significant group-level cytokine differences between Responders and Non-Responders does not undermine this interpretation: the continuous ΔButyrate– ΔIL-18 correlation in the full cohort reflects individual-level coupling between SCFA dynamics and inflammatory signaling, and such relationships are expected to be attenuated when patients are dichotomized by a composite classifier in a small sample. The two observations are therefore complementary rather than contradictory, and together support the hypothesis that post-TAVI SCFA remodeling constitutes a modifiable dimension of the post-procedural inflammatory milieu, warranting prospective evaluation in adequately powered cohorts.

Plasma butyrate is quantifiable by targeted metabolomics at pre- and post-procedural time points and lends itself to a simple, biologically grounded binary responder classification.

Interestingly, baseline isovalerate concentrations were higher among patients experiencing 1-month adverse events. Isovalerate is a branched-chain fatty acid produced through microbial catabolism of branched-chain amino acids, particularly leucine, and is considered a marker of amino-acid fermentation pathways.(31) Unlike canonical SCFAs such as butyrate, the clinical relevance of circulating isovalerate remains incompletely understood, with experimental studies suggesting context-dependent effects on host physiology.(32,33) Thus, the observed association may reflect differences in microbial metabolic adaptation rather than a direct detrimental effect of isovalerate itself.

Whether circulating SCFA profiling can serve as a biomarker of post-TAVI inflammatory recovery warrants prospective evaluation. Future studies should validate the SCFA responder phenotype in independent cohorts and determine whether gut-directed interventions such as dietary fiber optimization or synbiotic supplementation can modulate the butyrate–IL-18 axis and improve post-procedural recovery.

## LIMITATIONS

The reported findings need to be interpreted in light of several limitations. The modest sample size (n=40) constrains statistical power for all exploratory analyses, and adverse event comparisons are additionally underpowered and partly data-adaptive: the inflammatory composite was defined in part by the observed event distribution, which increases the risk of false-positive findings, and all adverse event comparisons should be regarded as hypothesis-generating pending pre-specified replication in an independent cohort. As a single-center observational study conducted at a tertiary referral center, the findings may not generalize to other settings or less selected patient populations; the absence of an untreated or medically managed control group further means that TAVI-specific effects on the gut microbiome-SCFA axis cannot be distinguished from temporal variation, natural disease progression, or concurrent medication changes. The observational design also precludes establishing directionality or causality for the ΔButyrate–ΔIL-18 association: dietary fiber intake, the primary determinant of colonic butyrate production, was not captured, limiting attribution of SCFA changes to the procedural event specifically. Because ΔButyrate and ΔIL-18 (and, in exploratory models, ΔIFN-γ and taxon-level deltas) were all derived from the same PRE-to-POST interval, temporal precedence between these variables cannot be established, and the sequential association models presented here should not be interpreted as confirming a causal mediation pathway; reverse or bidirectional influence cannot be excluded. Interval antibiotic use, dietary changes, and medication modifications between timepoints were not systematically recorded and could independently alter SCFA dynamics; moreover, although paired samples were processed within the same analytical batch, the storage interval between pre-procedural and post-procedural samples was not identical across patients, and differential biospecimen storage-time effects cannot be excluded. From a technical standpoint, 16S rRNA amplicon sequencing provides taxonomic rather than functional resolution; strain-level butyrate-producing capacity (butyryl-CoA:acetate CoA-transferase gene burden) is not captured, which explains the cross-omics gap between guild scores and plasma butyrate. Inflammasome activity was not directly measured; the mechanistic interpretation linking butyrate reduction to IL-18 elevation via NLRP3 is supported by existing experimental literature but not confirmed in this dataset. Finally, the Responder/Non-Responder classification is derived from the pre-to-post TAVI SCFA delta and therefore cannot be applied to periprocedural or 1-month events, which precede the 3-month post-TAVI measurement; adverse event analyses for these early windows instead use baseline SCFA concentrations as prospective predictors.

## CONCLUSION

In patients with severe AS undergoing TAVI, the procedure was associated with selective remodeling of the gut microbiome-derived metabolome, characterized by contraction of microbial richness, selective reduction in circulating butyrate and isovalerate, and a specific inverse coupling between individual butyrate dynamics and IL-18 trajectory that was independent of hemodynamic response, calcification burden, peri-procedural hematological stress, and cardiovascular risk factors. These findings identify a gut–metabolite–immune axis with properties consistent with inter-individual modulation of inflammasome-linked biological recovery following cardiovascular intervention, and support further investigation of the gut microbiome-derived metabolic activity as a candidate determinant of post-procedural immune adaptation.

## HIGHLIGHTS

- Post-TAVI systemic inflammation is heterogeneous, and gut microbiome-derived metabolic contributions remain unexplored.
- TAVI selectively remodels circulating SCFAs without restructuring overall gut microbial community composition.
- Post-TAVI butyrate decline inversely associates with IL-18 elevation, independent of hemodynamic correction.
- Prospective SCFA profiling and gut-directed intervention trials are needed to establish clinical utility.

## CLINICAL PERSPECTIVES

Competencies Addressed: Medical Knowledge; Patient Care and Procedural Skills; Practice-Based Learning and Improvement

### Clinical Competencies

Post-procedural systemic inflammation following TAVI is a recognized clinical challenge with consequences ranging from hemodynamic leaflet thrombosis to conduction system injury and new-onset atrial fibrillation. The present study demonstrates that individual variability in this inflammatory trajectory is associated with gut microbiome-derived metabolic changes, specifically a selective reduction in circulating butyrate, that are not explained by procedural success, hemodynamic improvement, or established cardiovascular risk factors. Clinicians involved in the peri-procedural management of TAVI patients should recognize that post-procedural inflammatory heterogeneity may reflect biological dimensions of recovery not captured by current assessment paradigms. While circulating SCFA profiling is not yet available in standard clinical practice, awareness of the gut-metabolite-immune axis as a candidate contributor to post-TAVI inflammatory risk broadens the conceptual framework for individualized post-procedural monitoring and positions this pathway as a target for future therapeutic development.

### Translational Outlook

Several steps are required before circulating SCFA profiling can enter the clinical translation pathway. First, the SCFA responder phenotype identified in this prospective cohort must be validated in independent, larger, and multicenter cohorts with pre-specified event ascertainment to confirm its predictive value for post-procedural inflammatory adverse events. Second, the mechanistic basis of the butyrate-IL-18 association has not been directly established in this dataset: experimental evidence linking butyrate to NLRP3 inflammasome suppression through histone deacetylase inhibition and GPR109A receptor signaling provides a plausible biological framework, but direct confirmation in human cardiovascular tissue is required. Third, analytical standardization of circulating SCFA measurement across clinical laboratories will be necessary before any biomarker application is feasible. Future research should prioritize prospective evaluation of the SCFA responder phenotype as a predictive biomarker of post-TAVI inflammatory risk, determine whether gut-directed interventions, including dietary fiber optimization and synbiotic supplementation, can modulate the butyrate-IL-18 axis after cardiac valve intervention, and assess whether such strategies translate to measurable reductions in post-procedural inflammatory complications.

## Supporting information

Main Tables

Supplementary data metagenomics

## DECLARATIONS

### Ethics approval and consent to participate

The study was approved by the local ethics committee (BASEC). All participants provided written informed consent prior to inclusion.

### Clinical Trial number

This study is registered at ClinicalTrials.gov (NCT07565077).

### Data Availability Statement

The datasets generated and/or analyzed during the current study are not publicly available due to patient privacy considerations but are available from the corresponding author on reasonable request.

### Conflicts of interest

Caroline Chong-Nguyen reports research grants from the French Society of Cardiology and the MIDHOS group, research funding from the Swiss Life Foundation, the Sana Foundation and the Peter Bockhoff Foundation, travel and educational grants from Abbott as well as consulting and proctoring for Abbott without personal remuneration.

Cyril Ferro reports travel expenses to the institution from Boston Scientific without personal remuneration.

Thomas Pilgrim reports research grants from the Swiss National Science Foundation, the Swiss Heart Foundation, the Swiss Polar Institute, the Bangerter-Rhyner Foundation, the Mach-Gaensslen Foundation, and the Monsol Foundation. Research, travel or educational grants to the institution without personal remuneration from Biotronik, Boston Scientific, Edwards Lifesciences, and ATSens; speaker fees and consultancy fees to the institution from Biotronik, Boston Scientific, Edwards Lifesciences, Abbott, Medtronic, Biosensors, and Highlife.

All other co authors declared no conflict of interest for this manuscript.

### Author contributions

Caroline Chong-Nguyen conceptualized and designed the study, supervised the project, performed data analysis and interpretation, and drafted the manuscript.

Cyril Ferro contributed to patient recruitment, clinical data collection, and manuscript revision.

Bahtiyar Yilmaz contributed to data interpretation, particularly in microbiome analyses.

Daijiro Tomii contributed to biological sample collection and biobanking

Camille Dupuy developed and performed metabolomics analyses.

Lydie Nadal-Desbarats developed and performed metabolomics analyses.

Pamela Nicholson performed metagenomics experiments.

Aparna Pandey performed statistical and bioinformatic analyses, particularly for metabolomics data.

Thomas Pilgrim contributed to study design, critically revised the manuscript, and provided senior supervision.

Yvonne Döring contributed to study design, critically revised the manuscript, and provided senior supervision.

All authors critically revised the manuscript and approved the final version.

## Acknowledgement

The authors gratefully acknowledge financial support from the Peter Bockhoff Foundation, the Sana Foundation, and the Swiss Life Foundation for the metabolomics and metagenomics analyses.

The summary Figure was created with BioRender.com

## ABBREVIATIONS

16S rRNA: 16S ribosomal RNA
ACEI: angiotensin-converting enzyme inhibitor
ARB: angiotensin receptor blocker
AS: aortic stenosis
ASC: apoptosis-associated speck-like protein
BMI: body mass index
CANTOS: Canakinumab Anti-inflammatory Thrombosis Outcomes Study
CAVD: calcific aortic valve disease
CT: computed tomography
FFAR: free fatty acid receptor
GPR: G-protein-coupled receptor
GPR109A: G-protein-coupled receptor 109A
HDAC: histone deacetylase
HIV: human immunodeficiency virus
IFN-γ: interferon-gamma
IL-1β: interleukin-1 beta
IL-5: interleukin-5
IL-13: interleukin-13
IL-17A: interleukin-17A
IL-17B: interleukin-17B
IL-18: interleukin-18
ILC: innate lymphoid cell
LC-MS/MS: liquid chromatography–tandem mass spectrometry
MACE: major adverse cardiovascular events
NF-κB: nuclear factor kappa B
NLRP3: NOD-like receptor protein 3
NYHA: New York Heart Association
PERMANOVA: permutational multivariate analysis of variance
QIIME2: Quantitative Insights Into Microbial Ecology 2
SC: straight-chain
SCFA: short-chain fatty acid
TAVI: transcatheter aortic valve implantation
TMAO: trimethylamine N-oxide
TNF-α: tumour necrosis factor-alpha
tnaA: tryptophanase gene
TRP: tryptophan

## Summary Figure

**Figure S1.**
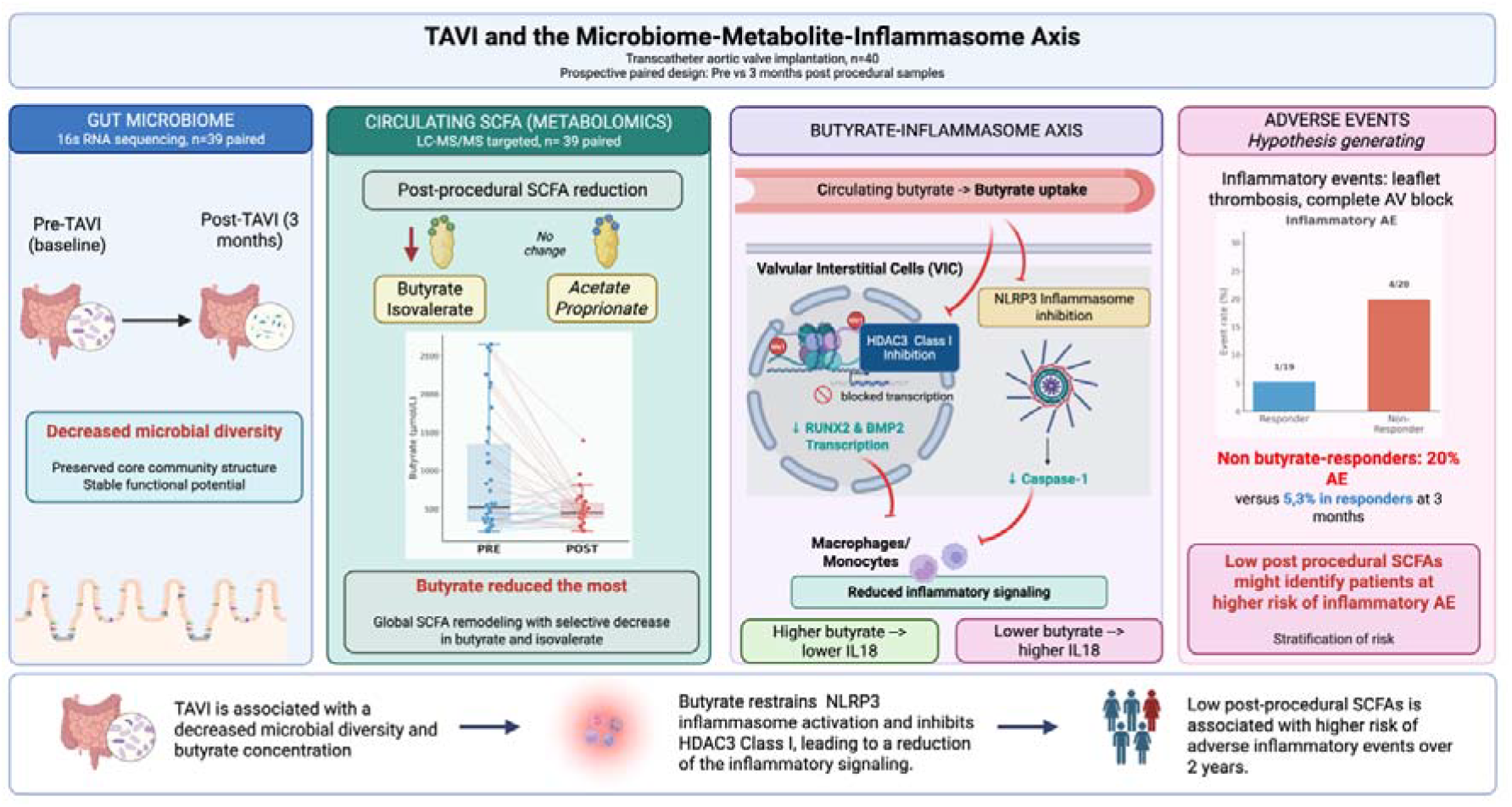
TAVI and the microbiome–metabolite–inflammasome axis. Blue panel — Gut microbiome. Stool samples collected before TAVI and three months after the procedure were analyzed by 16S rRNA gene sequencing. Post-procedural analysis demonstrated a reduction in microbial diversity, while the overall community structure and inferred functional potential remained largely preserved, indicating decreased richness without major disruption of community composition. Green panel — Circulating SCFAs. Targeted metabolomic profiling by LC-MS/MS revealed a selective post-procedural reduction in circulating butyrate and isovalerate concentrations, whereas acetate and propionate remained largely unchanged. Global SCFA remodeling was therefore primarily driven by decreased butyrate availability, with butyrate showing the largest magnitude of change. Purple panel — Butyrate–inflammasome axis (hypothesis-generating). This panel illustrates a proposed mechanism by which circulating butyrate may modulate inflammatory signaling. Based on experimental evidence from the literature, butyrate inhibits class I HDACs in valvular interstitial cells (VICs) and monocytes/macrophages, suppressing downstream transcriptional targets including RUNX2 and BMP2, and attenuating NLRP3 inflammasome activation. This results in reduced caspase-1 activity and lower IL-18 production. Consistent with this proposed framework, higher post-procedural butyrate concentrations were associated with lower circulating IL-18 levels, whereas lower butyrate concentrations were associated with higher IL-18 levels in the present study. The molecular interactions depicted were not directly assessed in this cohort and should be considered hypothesis-generating. Pink panel — Adverse events (hypothesis-generating). Patients with lower post-procedural butyrate concentrations (non-responders) exhibited a numerically higher incidence of inflammatory adverse events at three months compared with patients with preserved or increased butyrate levels (responders; 20% vs. 5.3%). Given the limited number of events, these findings are exploratory and hypothesis-generating, but suggest a potential link between post-procedural SCFA depletion and susceptibility to inflammatory adverse events. **Bottom panel** — Integrated summary. Collectively, these findings support an association between TAVI and reduced microbial diversity alongside decreased circulating butyrate concentrations. Reduced butyrate availability may contribute to persistent inflammatory activation through impaired suppression of NLRP3 inflammasome signaling in monocytes/macrophages and VICs, potentially increasing susceptibility to inflammatory adverse events. This proposed microbiome–metabolite–inflammation axis warrants further mechanistic and clinical investigation. *Abbreviations: AE, adverse event; AV, atrioventricular; BMP2, bone morphogenetic protein 2; HDAC, histone deacetylase; IL, interleukin; LC-MS/MS, liquid chromatography–tandem mass spectrometry; NLRP3, nucleotide-binding oligomerization domain-like receptor family pyrin domain-containing 3; RUNX2, runt-related transcription factor 2; SCFA, short-chain fatty acid; TAVI, transcatheter aortic valve implantation; VIC, valvular interstitial cell*.

