## Supplementary Tables for "Gut microbiome-derived metabolic remodeling and the butyrate–IL-18 inflammatory axis after transcatheter aortic valve implantation"

**Table of Contents**

1. Table S1. Alpha Diversity Metrics Before and After TAVI (n = 38 Paired Samples)
2. Table S2. PERMANOVA Results: Bray-Curtis Dissimilarity and Metabolite Class-Level Tests
3. Table S3. Circulating short-chain fatty acid concentrations before and after TAVI.
4. Table S4. Circulating inflammatory marker concentrations before and after TAVI.
5. Table S5. Sensitivity analyses for the ΔButyrate–ΔIL-18 association across pre-specified covariates.
6. Table S6. Baseline clinical characteristics by SCFA Responder status.

**Table S1. Circulating short-chain fatty acid concentrations before and after TAVI.**

*Paired SCFA metabolomics showed class-level remodeling after TAVI, driven by significant post-procedural declines in butyric acid and isovaleric acid, while acetic, propionic, and isobutyric acids remained unchanged after FDR correction.*

| **Metabolite** | **N (pairs)** | **PRE Median** | **PRE Q1** | **PRE Q3** | **POST Median** | **POST Q1** | **POST Q3** | **Δ Median (POST-PRE)** | **W** | **p (raw)** | **FDR q** | **Sig.** |
| --- | --- | --- | --- | --- | --- | --- | --- | --- | --- | --- | --- | --- |
| **Acetic acid** | 39 | 54'280.7 | 34'504.0 | 86'127.7 | 53'442.1 | 32'147.3 | 107'920.1 | -838.6 | 349 | 0.567 | 0.922 | ns |
| **Propionic acid** | 39 | 1'089.9 | 667.1 | 1'538.1 | 1'105.7 | 893.2 | 1'297.5 | 15.8 | 378 | 0.867 | 0.922 | ns |
| **Butyric acid** | 39 | 547.3 | 347.0 | 1'463.2 | 457.6 | 380.7 | 585.3 | -89.7 | 200 | 0.008 | 0.020 | * ↓ |
| **Isobutyric acid** | 39 | 398.1 | 342.8 | 502.4 | 416.7 | 349.4 | 468.5 | 18.6 | 383 | 0.922 | 0.922 | ns |
| **Valeric acid** | 1 | — | — | — | — | — | — | — | — | — | — | n/a |
| **Isovaleric acid** | 39 | 983.0 | 661.9 | 1'272.4 | 681.1 | 530.7 | 821.7 | -301.9 | 173 | 0.003 | 0.012 | * ↓ |

* ↓ = FDR q < 0.05 within the SCFA class; significant post-TAVI decline. ns = not significant after FDR correction. n/a = not tested because valid paired data were insufficient. Values are shown in nmol/L; Δ Median is POST minus PRE.

**Table S2. Circulating inflammatory marker concentrations before and after TAVI.**

*Cytokine concentrations were stable at the cohort level after TAVI after FDR correction; IL-1β showed only a nominal decrease and did not remain significant after correction.*

| **Cytokine** | **n pairs** | **PRE Median** | **PRE Q1** | **PRE Q3** | **POST Median** | **POST Q1** | **POST Q3** | **Δ median** | **p (Wilcoxon)** | **q (BH-FDR)** | **Sig.** |
| --- | --- | --- | --- | --- | --- | --- | --- | --- | --- | --- | --- |
| **IL-18** | 36 | 629.66 | 448.2 | 785.13 | 593.37 | 401.08 | 808.88 | -51.02 | 0.7059 | 0.9827 | NS |
| **IL-1β** | 36 | 0.38 | 0.35 | 0.44 | 0.35 | 0.32 | 0.38 | -0.02 | 0.0175 | 0.1225 | * (p only) |
| **IFN-γ** | 36 | 10.6 | 7.94 | 18.25 | 11.99 | 8.42 | 17.79 | -0.15 | 0.9827 | 0.9827 | NS |
| **TNF-α** | 36 | 2.55 | 2.14 | 3.5 | 2.86 | 1.95 | 3.75 | 0.29 | 0.3682 | 0.6444 | NS |
| **IL-17A** | 36 | 3.3 | 2.76 | 5.88 | 3.06 | 2.31 | 4.9 | -0.35 | 0.2641 | 0.6444 | NS |
| **IL-17B** | 36 | 0.65 | 0.5 | 0.86 | 0.7 | 0.46 | 0.98 | 0.06 | 0.3425 | 0.6444 | NS |
| **IL-5** | 35 | 0.88 | 0.65 | 1.23 | 0.92 | 0.47 | 1.35 | 0.03 | 0.9741 | 0.9827 | NS |

BH-FDR correction was applied across the cytokine panel. * (p only) indicates nominal p < 0.05 without FDR significance. NS = not significant after correction. Values are reported in the assay units used in the source table; Δ median is POST minus PRE.

**Table S3. Alpha Diversity Metrics Before and After TAVI (n = 38 Paired Samples)**

*Paired Wilcoxon signed-rank test (n = 38 complete pairs; 1 patient missing POST sample excluded). Δ = POST − PRE. * p < 0.05; ns = not significant. Green highlight: metrics reaching p < 0.05 (Chao1 and Observed species). Shannon and Simpson: not significant.*

|  | **PRE** | | **POST** | | **Δ (POST − PRE)** | | **Wilcoxon** |  |  |
| --- | --- | --- | --- | --- | --- | --- | --- | --- | --- |
| **Metric** | **Mean** | **Median** | **Mean** | **Median** | **Mean (Δ)** | **Median (Δ)** | **p-value** | **Significance** | **n (pairs)** |
| **Chao1** | 128,230 | 128,500 | 120,110 | 125,080 | -8,120 | -4,880 | 0,0214 | * | 38 |
| **Observed species** | 127,390 | 128,000 | 119,240 | 125,000 | -8,160 | -6,000 | 0,0248 | * | 38 |
| **Shannon** | 3,047 | 3,076 | 2,952 | 3,119 | -0,095 | -0,073 | 0,1470 | ns | 38 |
| **Simpson** | 0,875 | 0,903 | 0,860 | 0,907 | -0,014 | -0,007 | 0,2400 | ns | 38 |

**Table S4. PERMANOVA Results: Bray-Curtis Dissimilarity and Metabolite Class-Level Tests**

*All PERMANOVA analyses use 9,999 permutations. Bray-Curtis dissimilarity computed on rarefied 16S rRNA amplicon data. Metabolite class tests computed on paired z-scored log₂ values using squared Euclidean distance (paired PERMANOVA stratified within patients). * p < 0.05; ** p < 0.01; *** p < 0.001; ns = not significant.*

| **Analysis** | **Distance metric** | **Design** | **n (pairs)** | **Df** | **pseudo-F** | **R²** | **p-value** |
| --- | --- | --- | --- | --- | --- | --- | --- |
| **A. Microbiome beta-diversity (16S rRNA amplicon sequencing, n = 38 paired samples)** | | | | | | | |
| Bray-Curtis — unpaired (PRE vs POST group comparison) | Bray-Curtis | Unpaired (group) | 38 | 1 | 0,363 | 0,0048 | 1.000 (ns) |
| Bray-Curtis — paired (stratified permutation by patient ID) | Bray-Curtis | Paired (strat. by patient) | 38 | 2 | 0,929 | 0,0248 | 0.692 (ns) |
| **B. Metabolite class-level PERMANOVA (paired, stratified permutation; n = paired samples per class)** | | | | | | | |
| SCFA (short-chain fatty acids) Included: Acetic acid, Propionic acid, Butyric acid, Isobutyric acid, Isovaleric acid Excluded: Valeric acid (84.6% < LLOQ) | Sq. Euclidean (z-scored log₂) | Paired (strat. by patient) | 39 |  | 2,971 | 0,038 | 0.014 * |
| Cytokines (inflammatory markers) Included: IFN-γ, IL-17A, IL-17B, IL-18, IL-1β, IL-5, TNF-α Excluded: IL-13 (72.9% missing) | Sq. Euclidean (z-scored log₂) | Paired (strat. by patient) | 35 |  | 0,788 | 0,011 | 0.630 (ns) |

**Table S5. Sensitivity analyses for the ΔButyrate–ΔIL-18 association across pre-specified covariates.**

*The inverse ΔButyrate–ΔIL-18 association remained robust after adjustment for aortic valve calcification, peri-procedural haemoglobin change, cardiovascular medications, and baseline demographics, supporting independence from major measured confounders.*

| **Covariate / Model** | **Adjustment** | **n** | **Spearman ρ** | **95% CI low** | **95% CI high** | **p-value** | **Supp. figure** |
| --- | --- | --- | --- | --- | --- | --- | --- |
| **A. Reference** |  |  |  |  |  |  |  |
| **Crude (unadjusted)** | None | 36 | -0.668 | -0.82 | -0.412 | < 0.0002 *** | — |
| **B. Aortic valve calcification burden (Ca score)** |  |  |  |  |  |  |  |
| **Calcium score (CT)** | Calcium score (continuous) | 34 | -0.732 | — | — | < 0.001 *** | Figure S11 |
| **C. Peri-procedural haematological stress (ΔHb)** |  |  |  |  |  |  |  |
| **ΔHaemoglobin — partial correlation (all patients)** | ΔHb (continuous) | 36 | -0.602 | -0.843 | -0.462 | < 0.0002 *** | Figure S12 |
| **ΔHaemoglobin — low-stress stratum (ΔHb ≤ −1.00 g/dL)** | ΔHb stratum (n = 18) | 18 | -0.335 | -0.744 | 0.253 | 0.171 (ns) | Figure S12 |
| **ΔHaemoglobin — high-stress stratum (ΔHb > −1.00 g/dL)** | ΔHb stratum (n = 18) | 18 | -0.734 | -0.905 | -0.325 | 0.0008 *** | Figure S12 |
| **D. Cardiovascular medications** |  |  |  |  |  |  |  |
| **Statin use** | Statin (binary) | 36 | -0.686 | -0.846 | -0.436 | < 0.001 *** | Figure S13 |
| **ACE inhibitor (ACEI) use** | ACEI (binary) | 36 | -0.678 | -0.836 | -0.405 | < 0.001 *** | Figure S13 |
| **Aspirin use** | Aspirin (binary) | 36 | -0.715 | -0.849 | -0.474 | < 0.001 *** | Figure S13 |
| **Joint medications (statin + ACEI + aspirin)** | Statin + ACEI + Aspirin | 36 | -0.74 | -0.87 | -0.51 | < 0.001 *** | Figure S13 |
| **E. Baseline demographics** |  |  |  |  |  |  |  |
| **BMI** | BMI (kg/m², continuous) | 36 | -0.613 | -0.808 | -0.266 | 0.0004 *** | Figure S14 |
| **Age** | Age (years, continuous) | 36 | -0.68 | -0.856 | -0.409 | < 0.0002 *** | Figure S14 |
| **Sex** | Sex (0 = female, 1 = male) | 36 | -0.655 | -0.822 | -0.389 | < 0.0002 *** | Figure S14 |
| **Joint demographics (BMI + age + sex)** | BMI + Age + Sex | 36 | -0.624 | -0.845 | -0.253 | 0.0002 *** | Figure S14 |

Partial Spearman ρ was computed by residual-rank adjustment. Bootstrap confidence intervals and permutation p-values are shown where available. Reference model: crude ρ = −0.668, 95% CI [−0.820, −0.412], p < 0.0002, n = 36. * p < 0.05; ** p < 0.01; *** p < 0.001.

**Supplementary Table S6. Baseline clinical characteristics by SCFA Responder status.**

| **Variable** | **Responders (n=19)** | **Non-Responders (n=20)** | **p-value** | **Sig.** |
| --- | --- | --- | --- | --- |
| **Age (years)** | 81.0 (78.0-85.0), n=19 | 81.5 (78.5-85.2), n=20 | 0,682 | ns |
| **BMI (kg/m2)** | 24.4 (22.7-26.8), n=19 | 26.6 (23.2-30.9), n=20 | 0,095 | ns |
| **eGFR, Cockcroft-Gault (mL/min)** | 55.7 (49.5-66.3), n=19 | 62.8 (54.5-70.5), n=20 | 0,279 | ns |
| **Hemoglobin (g/dL)** | 13.0 (12.1-14.1), n=19 | 13.1 (12.2-14.0), n=20 | 1,000 | ns |
| **CRP (mg/L)** | 2.0 (1.0-4.0), n=11 | 1.0 (1.0-2.2), n=12 | 0,340 | ns |
| **LVEF (%)** | 60.0 (52.0-68.0), n=19 | 61.5 (60.0-66.0), n=20 | 0,591 | ns |
| **Mean aortic gradient (mmHg)** | 38.0 (31.5-46.5), n=19 | 41.5 (31.8-50.8), n=20 | 0,555 | ns |
| **Aortic valve area (cm2)** | 0.8 (0.7-0.9), n=19 | 0.8 (0.7-1.0), n=20 | 0,933 | ns |
| **Indexed AVA (cm2/m2)** | 0.4 (0.4-0.5), n=19 | 0.4 (0.4-0.5), n=20 | 0,474 | ns |
| **Aortic valve calcium score (AU)** | 2740.0 (2347.0-3220.0), n=17 | 2875.5 (1599.5-3493.5), n=20 | 0,915 | ns |
| **NYHA class** | 2.0 (1.0-2.0), n=19 | 2.0 (1.8-2.0), n=20 | 0,740 | ns |
| **QRS duration (ms)** | 95.0 (85.5-118.5), n=19 | 92.5 (84.0-112.0), n=20 | 0,536 | ns |
| **STS risk of mortality (%)** | 3.0 (2.3-4.3), n=17 | 2.5 (1.7-3.8), n=20 | 0,369 | ns |
| **EuroSCORE II (%)** | 1.9 (1.7-3.7), n=17 | 1.8 (1.3-2.8), n=20 | 0,175 | ns |
| **Female sex** | 11/19 (57.9%) | 9/20 (45.0%) | 0,527 | ns |
| **Diabetes mellitus** | 6/19 (31.6%) | 3/20 (15.0%) | 0,273 | ns |
| **Hypertension** | 15/19 (78.9%) | 17/20 (85.0%) | 0,695 | ns |
| **Dyslipidemia** | 9/19 (47.4%) | 12/20 (60.0%) | 0,527 | ns |
| **Atrial fibrillation (any)** | 5/19 (26.3%) | 6/20 (30.0%) | 1,000 | ns |
| **Pre-existing pacemaker** | 5/19 (26.3%) | 3/20 (15.0%) | 0,451 | ns |
| **Coronary artery disease** | 5/19 (26.3%) | 3/20 (15.0%) | 0,451 | ns |
| **Chronic kidney disease (any stage)** | 2/19 (10.5%) | 1/20 (5.0%) | 0,605 | ns |
| **COPD** | 2/19 (10.5%) | 1/20 (5.0%) | 0,605 | ns |
| **Sinus rhythm (baseline ECG)** | 13/19 (68.4%) | 15/20 (75.0%) | 0,731 | ns |
| **AV block present (baseline ECG)** | 5/19 (26.3%) | 3/19 (15.8%) | 0,693 | ns |
| **QRS morphology: BBB vs Normal** | 5/18 (27.8%) | 3/16 (18.8%) | 0,693 | ns |
| **Diabetes mellitus (SwissTAVI cross-check)** | 6/17 (35.3%) | 2/20 (10.0%) | 0,109 | ns |
| **Valve type: self- vs balloon-expandable** | 8/17 (47.1%) | 5/20 (25.0%) | 0,188 | ns |
| **Access route: right vs left femoral** | 15/17 (88.2%) | 19/20 (95.0%) | 0,584 | ns |

**Table S6.** SCFA Responders (n=19) were defined as patients with a composite standardised z-score above zero for the pre-to-post TAVI change in butyrate and isovalerate, the two SCFAs that declined significantly at the cohort level; Non-Responders (n=20) had a composite z-score at or below zero. One patient with no post-TAVI SCFA sample was excluded (total N=39). Continuous variables are expressed as median (interquartile range) and compared using the Mann-Whitney U test, two-sided. Categorical variables are expressed as n (%) and compared using Fisher’s exact test, two-sided. No correction for multiple comparisons was applied; comparisons are descriptive. Variables with more than 20% missing data are indicated with a dagger symbol (CRP: available in 23/39 patients).

*Abbreviations: SCFA, short-chain fatty acid; TAVI, transcatheter aortic valve implantation; IQR, interquartile range; BMI, body mass index; eGFR, estimated glomerular filtration rate (Cockcroft-Gault); LVEF, left ventricular ejection fraction; AVA, aortic valve area; NYHA, New York Heart Association; STS, Society of Thoracic Surgeons; EuroSCORE, European System for Cardiac Operative Risk Evaluation; CT, computed tomography; AU, Agatston units; LBBB, left bundle branch block; RBBB, right bundle branch block; AVB, atrioventricular block; BBB, bundle branch block; AF, atrial fibrillation; CAD, coronary artery disease; CKD, chronic kidney disease; COPD, chronic obstructive pulmonary disease; CRP, C-reactive protein.*
