## Supplementary Figures for "Gut microbiome-derived metabolic remodeling and the butyrate–IL-18 inflammatory axis after transcatheter aortic valve implantation"

**Supplementary Tables and Figures**

**Table of Contents**

**Supplementary Figures**

1. Figure S1. Alpha diversity before and after TAVI
   a. 1.1 Figure S1A. Chao1 diversity: PRE vs POST TAVI
   b. 1.2 Figure S1B. Observed diversity: PRE vs POST TAVI
2. Figure S2. Per-patient changes in alpha diversity after TAVI
   a. 2.1 Figure S2A. Per-patient ΔShannon diversity
   b. 2.2 Figure S2B. Per-patient ΔSimpson diversity
   c. 2.3 Figure S2C. Per-patient ΔObserved diversity
3. Figure S3. Bray–Curtis PCoA of microbial community structure before and after TAVI
4. Figure S4. Within-patient versus between-patient Bray–Curtis distance
5. Figure S5. Refined metabolic functional capacity before and after TAVI
   a. 5.1 Figure S5A. Functional guild overview
   b. 5.2 Figure S5B. True butyrate synthesis: PRE vs POST TAVI
   c. 5.3 Figure S5C. Proteolytic degradation: PRE vs POST TAVI
6. Figure S6. Cross-omics correlations between functional guild scores and plasma SCFA changes
7. Figure S7. Correlations between metabolite changes and haemodynamic changes
8. Figure S8. Association analysis of ΔButyrate, ΔIL-18, and ΔIFN-γ
9. Figure S9. Inflammatory marker profiles before and after TAVI
   a. 9.1 Figure S9A. PCA of inflammatory markers: PRE vs POST TAVI
   b. 9.2 Figure S9B. ΔPCA of inflammatory marker changes
10. Figure S10. Tryptophan-pathway metabolites and ΔIL-18
    a. 10.1 Figure S10A. TRP metabolite screen against ΔIL-18
    b. 10.2 Figure S10B. Parallel covariate analysis of ΔButyrate and ΔKynurenine
11. Figure S11. Partial Spearman correlation between ΔButyrate and ΔIL-18 adjusted for aortic calcium score
12. Figure S12. Sensitivity analyses for peri-procedural haemoglobin change
    a. 12.1 Figure S12A. Crude versus partial correlation controlling for ΔHaemoglobin
    b. 12.2 Figure S12B. High ΔHb-drop stratum
    c. 12.3 Figure S12C. Low ΔHb-drop stratum
13. Figure S13. Medication-adjusted sensitivity analyses
    a. 13.1 Figure S13A. Adjustment for statin, ACEI, and aspirin use
    b. 13.2 Figure S13B. Stratified analysis by medication use
14. Figure S14. Demographic-adjusted sensitivity analysis for baseline BMI, age, and sex
15. Figure S15. Falsification tests for biologically implausible outcomes
16. Figure S16. Baseline serum SCFA concentrations by 1-month adjudicated event status.
17. Figure S17. Baseline serum isovalerate concentration and 1-month adjudicated adverse events.
18. Figure S18. Inflammatory adverse event rates at 3-month follow-up by SCFA Responder status.

**Figure S1A. Chao1 richness before and after TAVI**

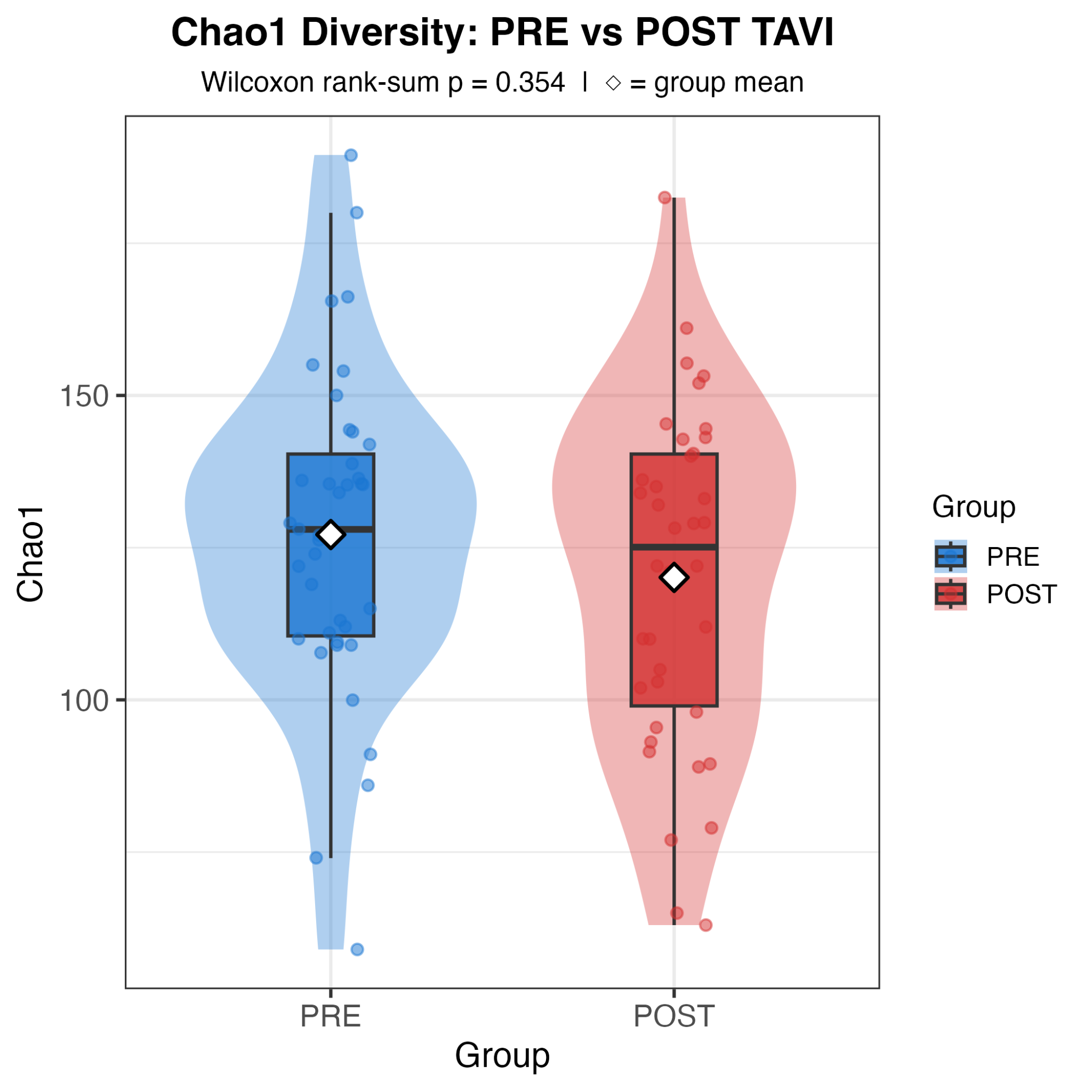

**Figure S1A.** Paired violin and boxplots show that Chao1 richness decreased after TAVI, supporting the manuscript finding of post-procedural contraction of microbial richness.

**Figure S1B. Observed species before and after TAVI**

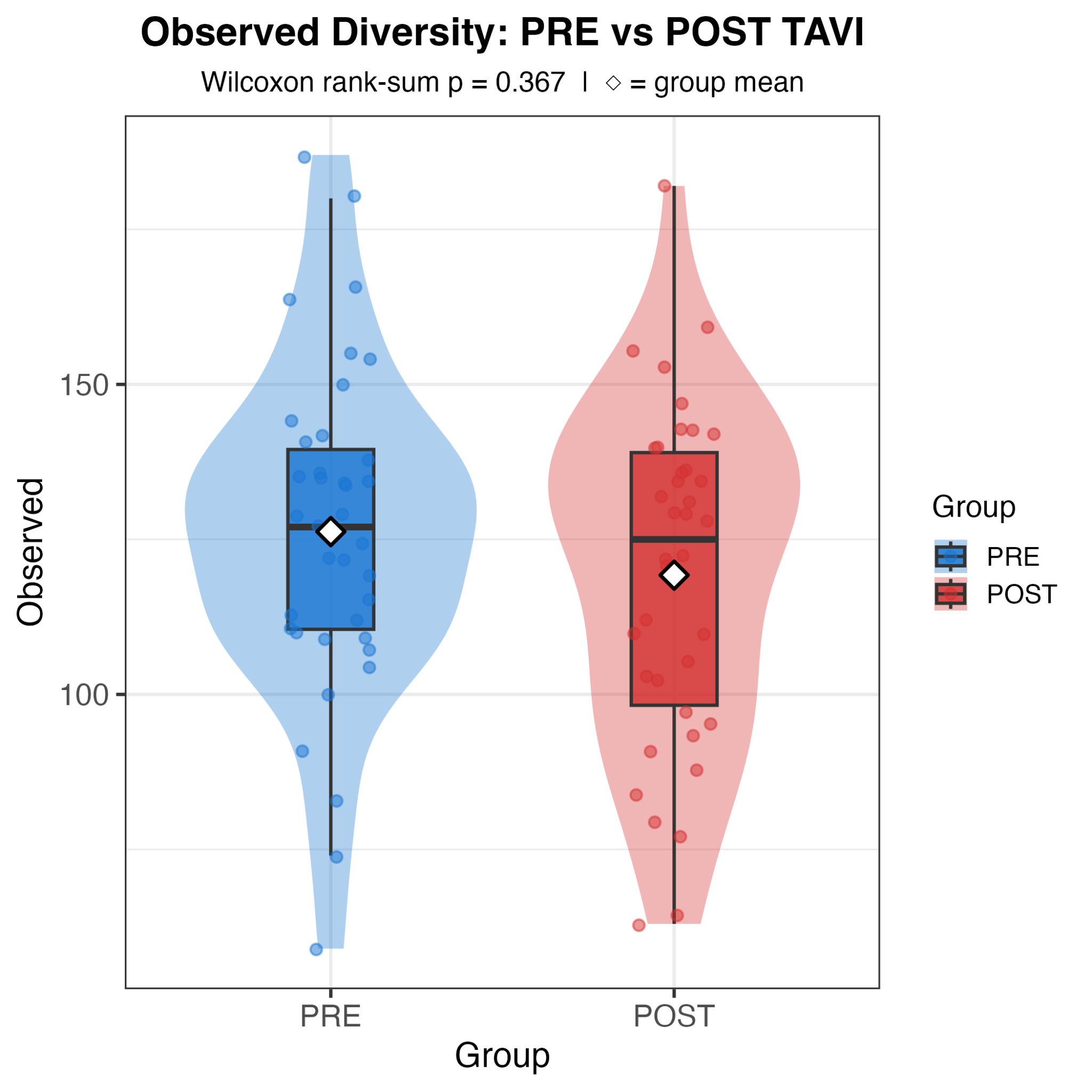

**Figure S1B.** Paired violin and boxplots show a reduction in observed species counts after TAVI, consistent with reduced microbial richness rather than broad dysbiosis.

**Figure S2A. Per-patient Shannon entropy changes after TAVI**

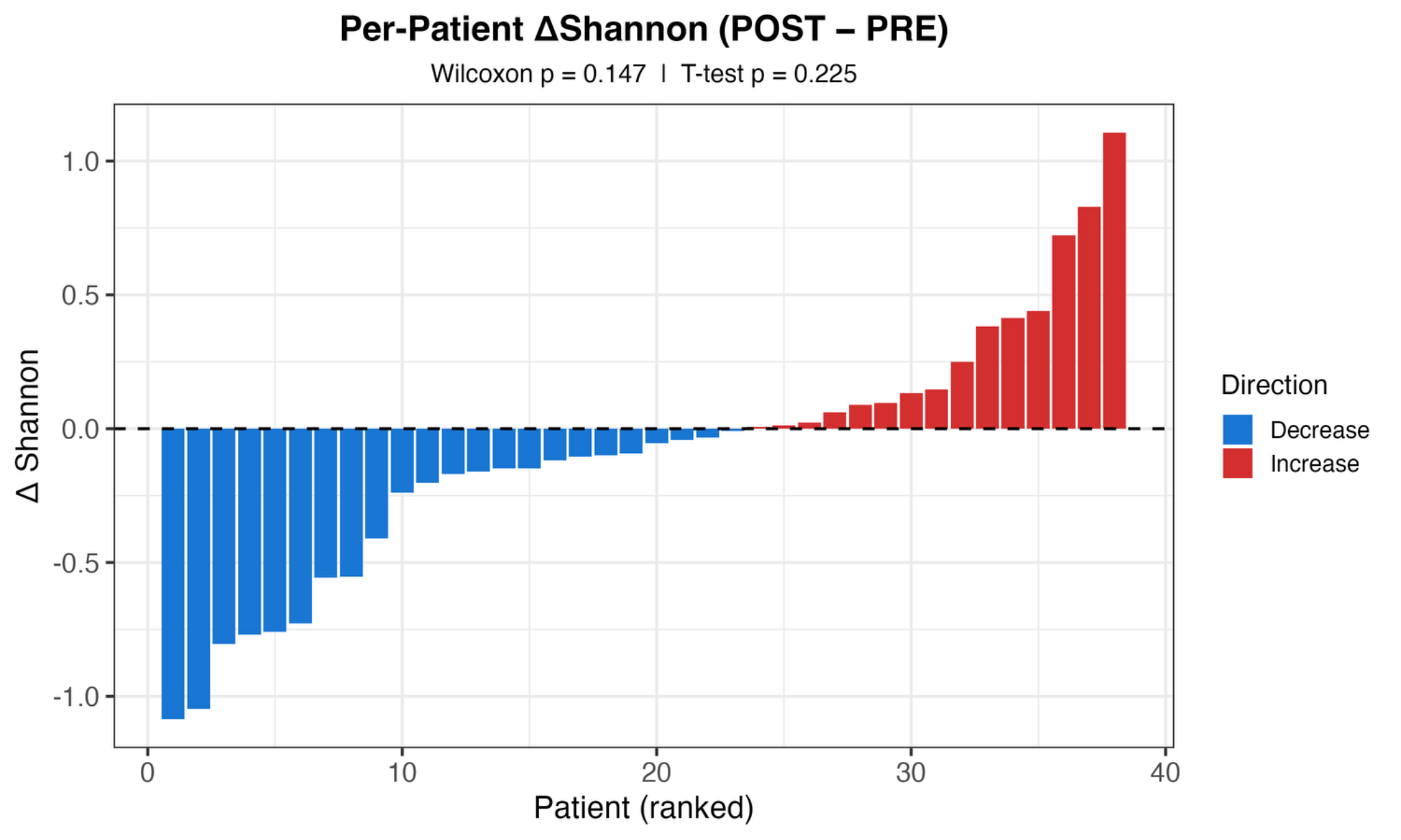

**Figure S2A.** The waterfall plot displays individual Shannon entropy changes from PRE to POST. Changes are heterogeneous and do not indicate a significant cohort-level shift in Shannon diversity.

**Figure S2B. Per-patient Simpson index changes after TAVI**

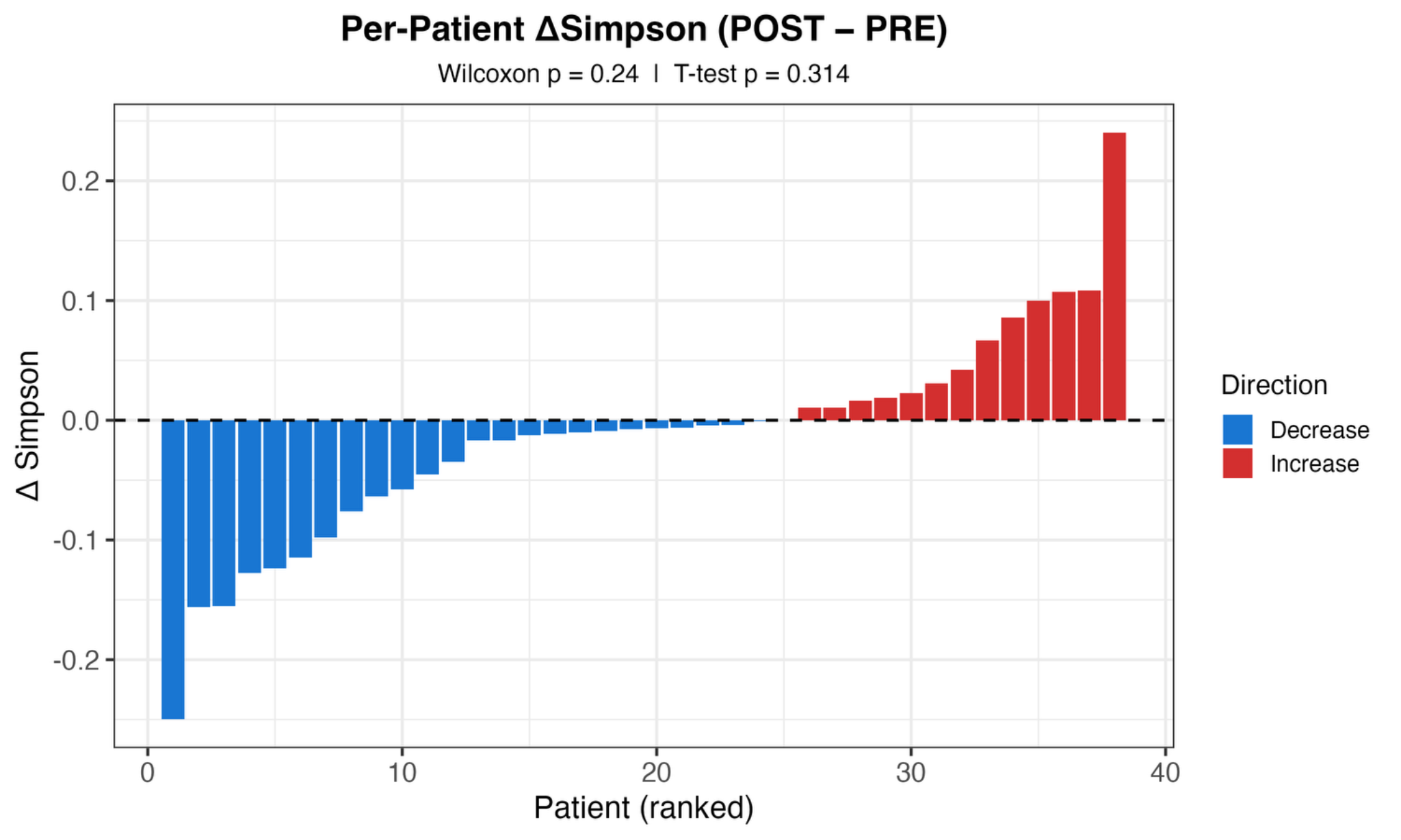

**Figure S2B.** The waterfall plot shows patient-level Simpson index changes after TAVI. The absence of a uniform directional pattern supports preservation of overall alpha diversity.

**Figure S2C. Per-patient observed-species changes after TAVI**

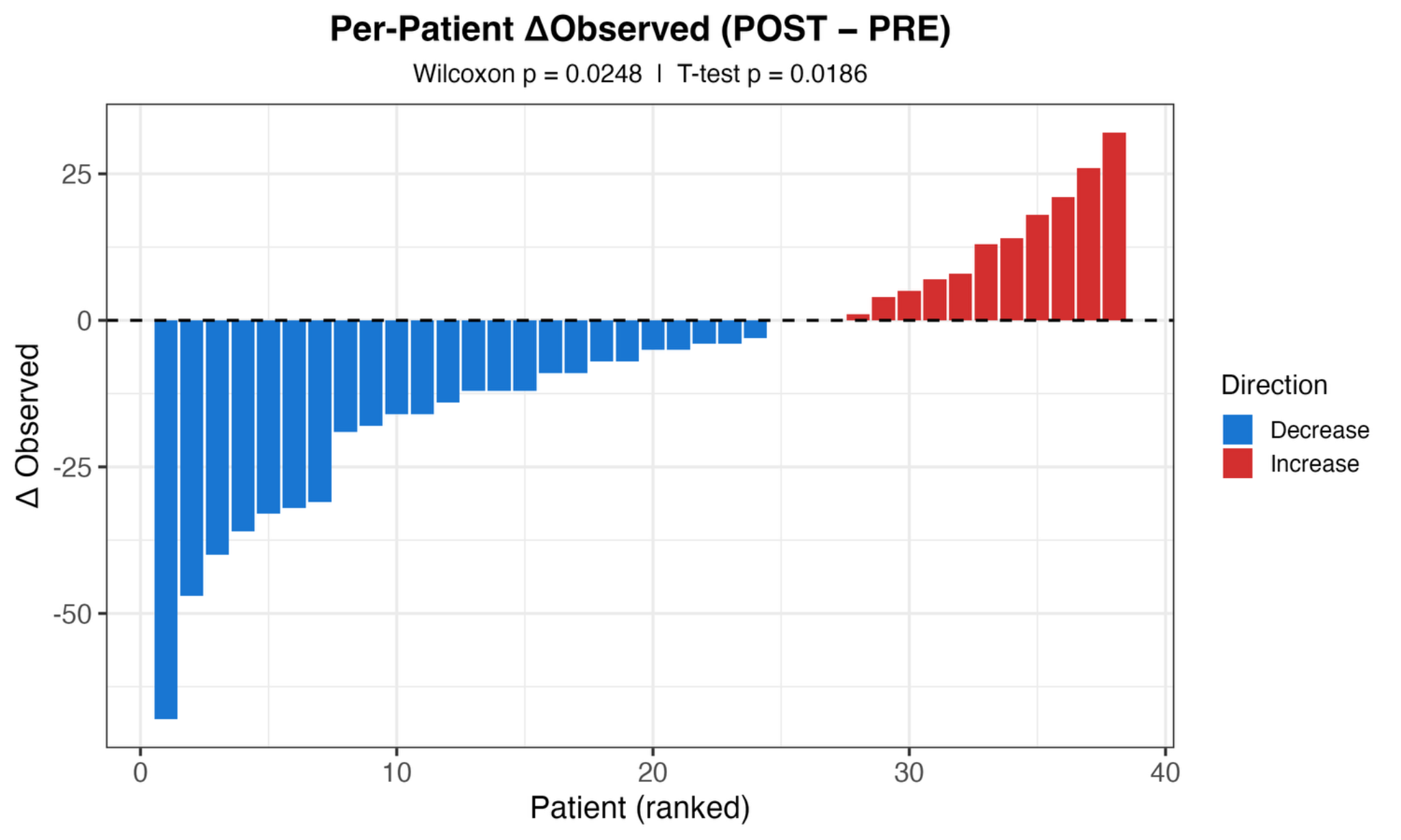

**Figure S2C.** The ranked individual delta plot illustrates heterogeneous patient trajectories in observed species counts, while the overall cohort pattern supports reduced richness after TAVI.

**Figure S3. Bray-Curtis community composition before and after TAVI**

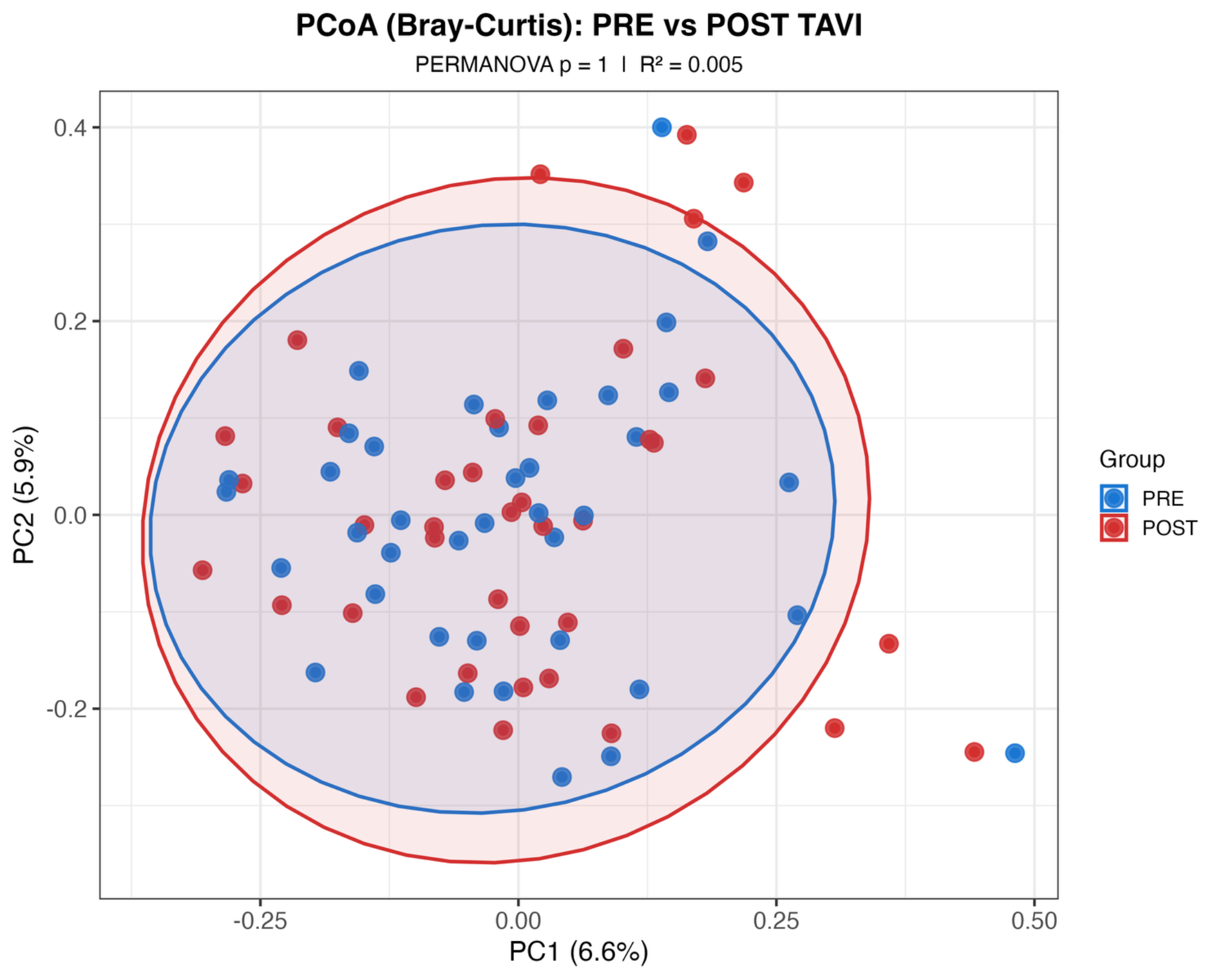

**Figure S3.** Principal coordinate analysis of Bray-Curtis dissimilarity shows that global microbiome community composition was not significantly restructured after TAVI.

**Figure S4. Within-patient and between-patient Bray-Curtis distances**

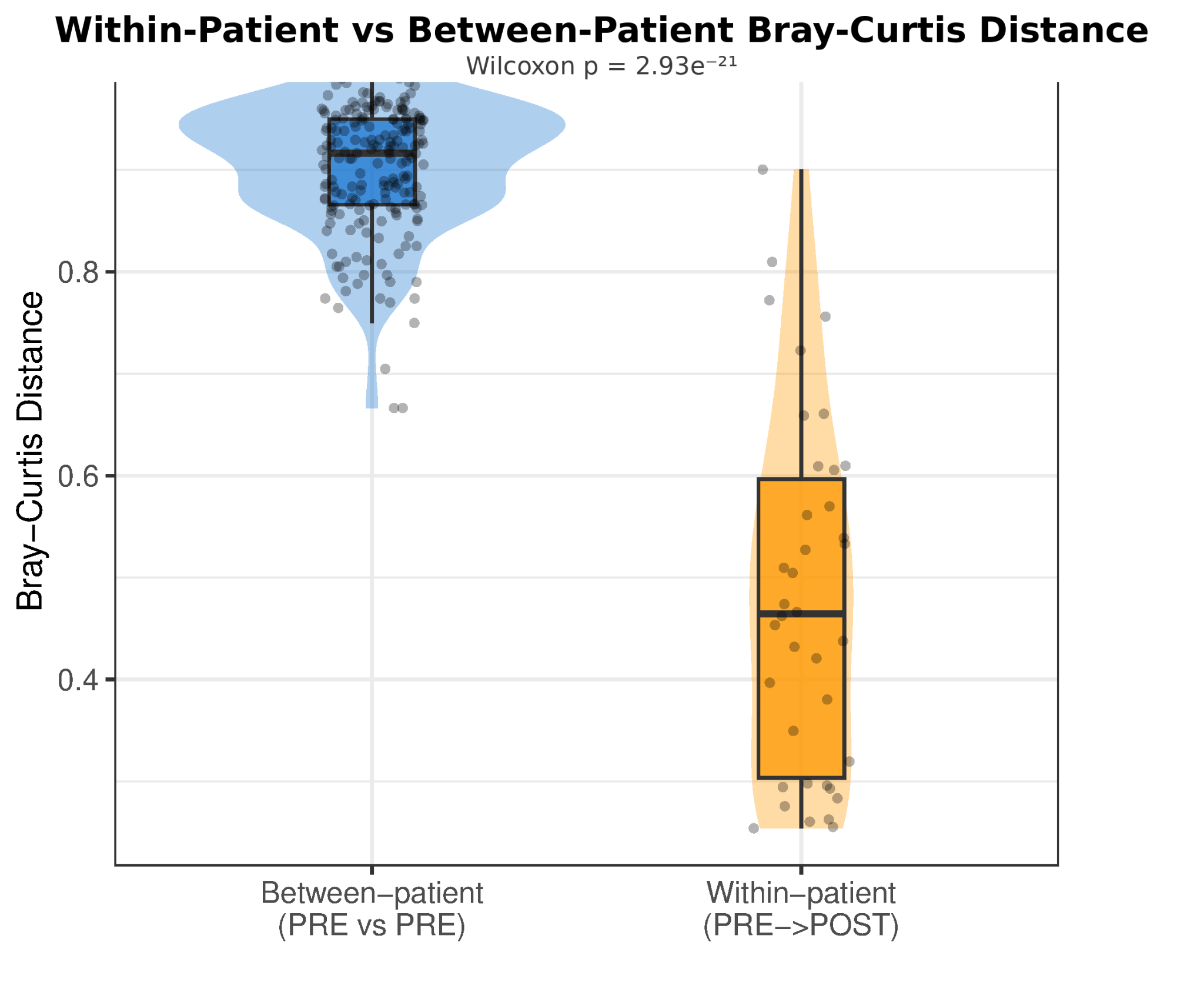

**Figure S4.** The distance comparison shows that within-patient PRE-to-POST shifts are smaller than between-patient distances, indicating that inter-individual microbiome structure remains dominant.

**Figure S5A. Functional guild trajectories after TAVI**

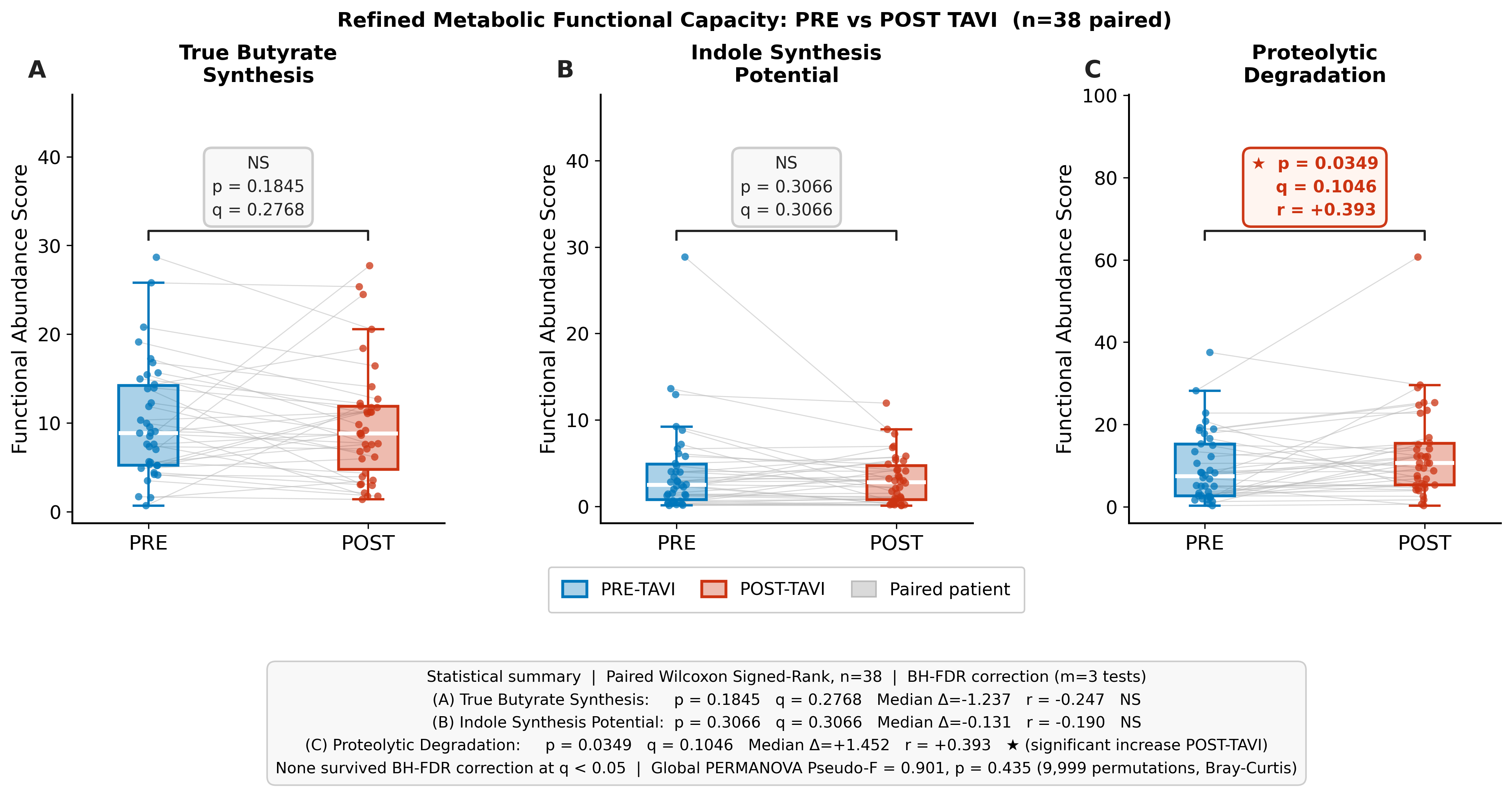

**Figure S5A.** Boxplots summarize pre-to-post changes across inferred microbial functional guilds. Guild scores varied across patients but did not show significant cohort-level change after false-discovery correction.

**Figure S5B. True butyrate-synthesis guild before and after TAVI**

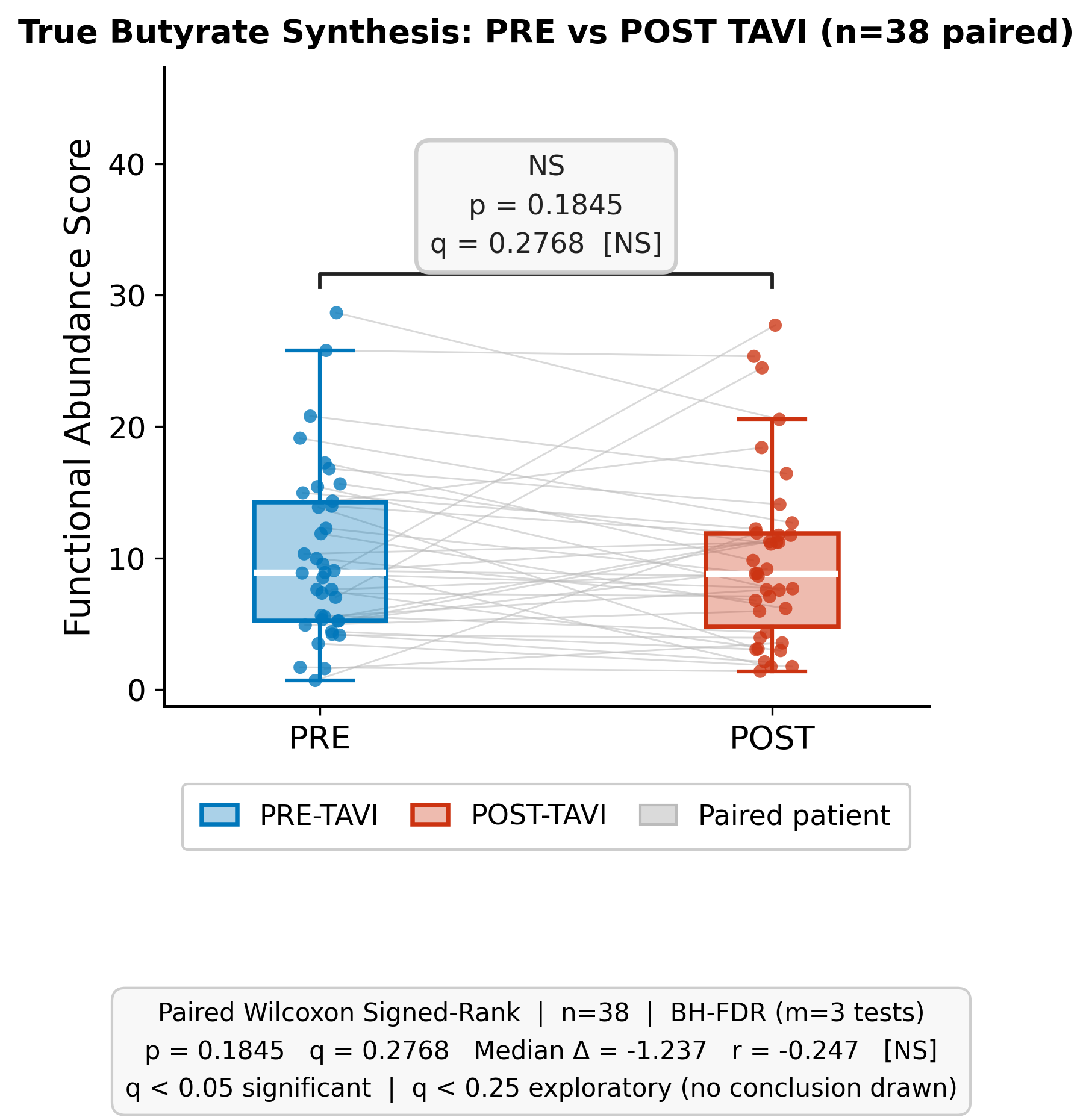

**Figure S5B.** The paired comparison focuses on the true butyrate-synthesis guild and shows no significant cohort-level change after TAVI.

**Figure S5C. Proteolytic degradation guild before and after TAVI**

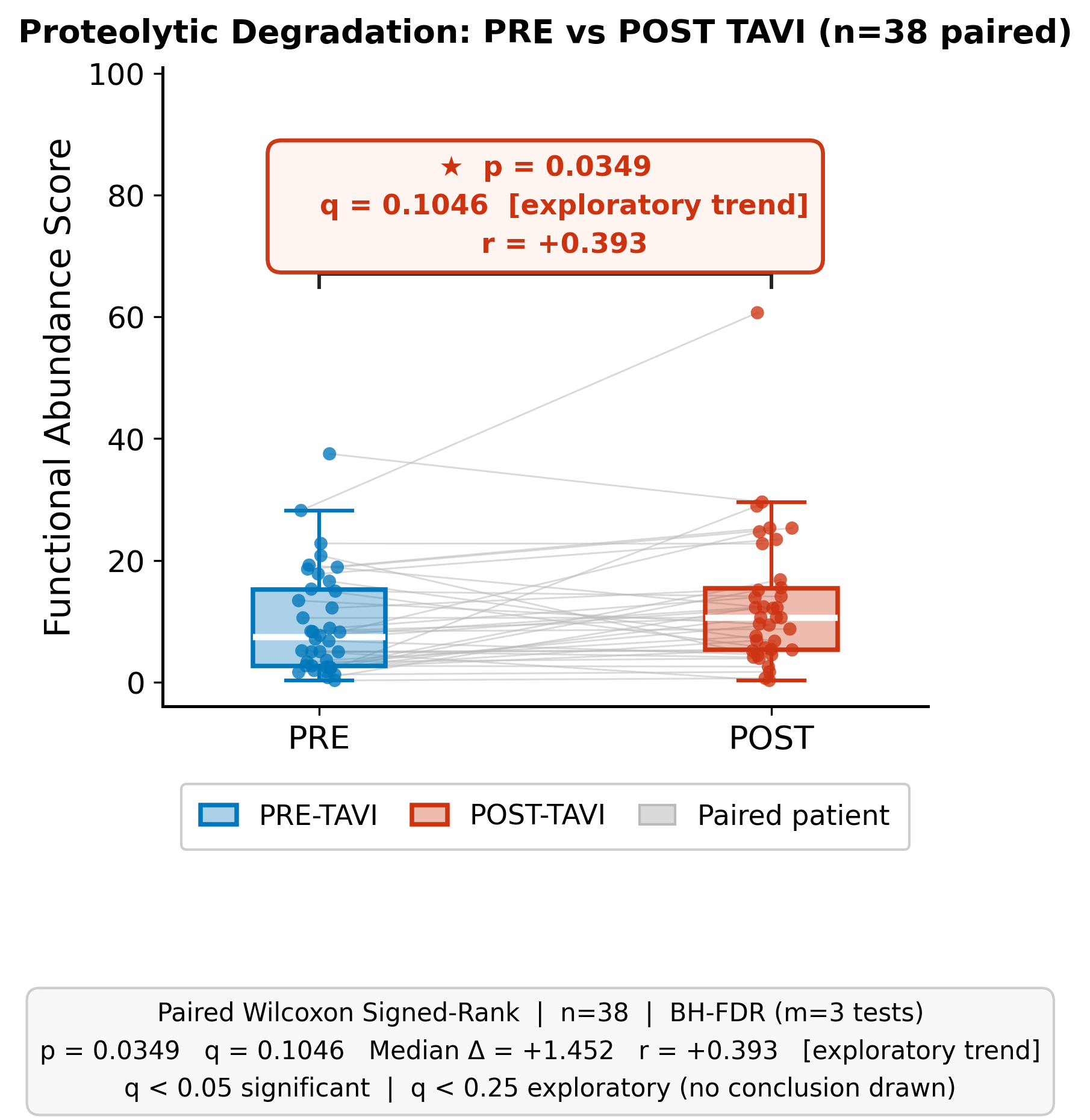

**Figure S5C.** The paired comparison of the proteolytic degradation guild shows no significant cohort-level pre-to-post change after TAVI.

**Figure S6. Butyrate-producing guild score and circulating butyrate**

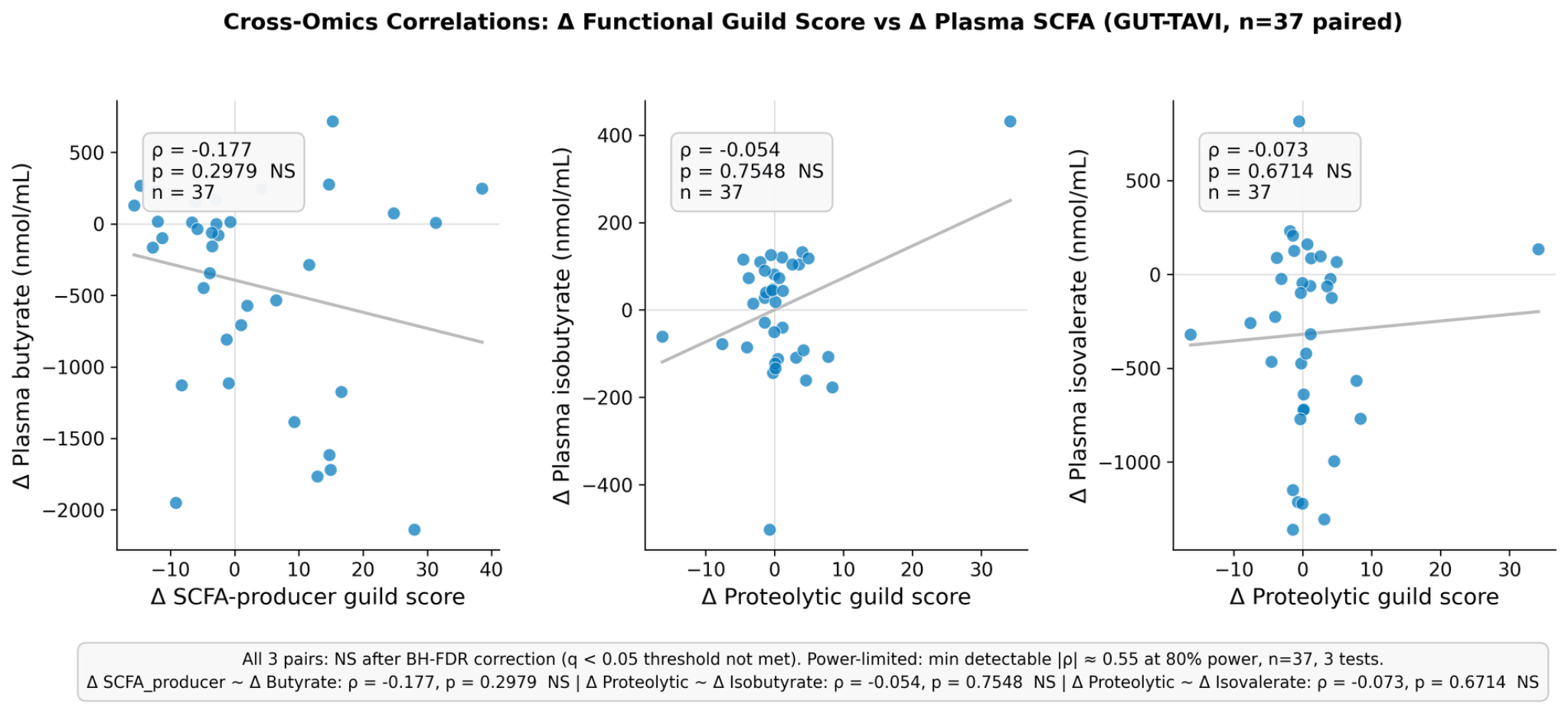

**Figure S6.** The cross-omics analysis shows that the 16S-derived butyrate-producing guild score did not correlate with circulating butyrate, indicating that plasma butyrate dynamics are not captured by this guild-level proxy.

**Figure S7. SCFA changes and haemodynamic improvement**

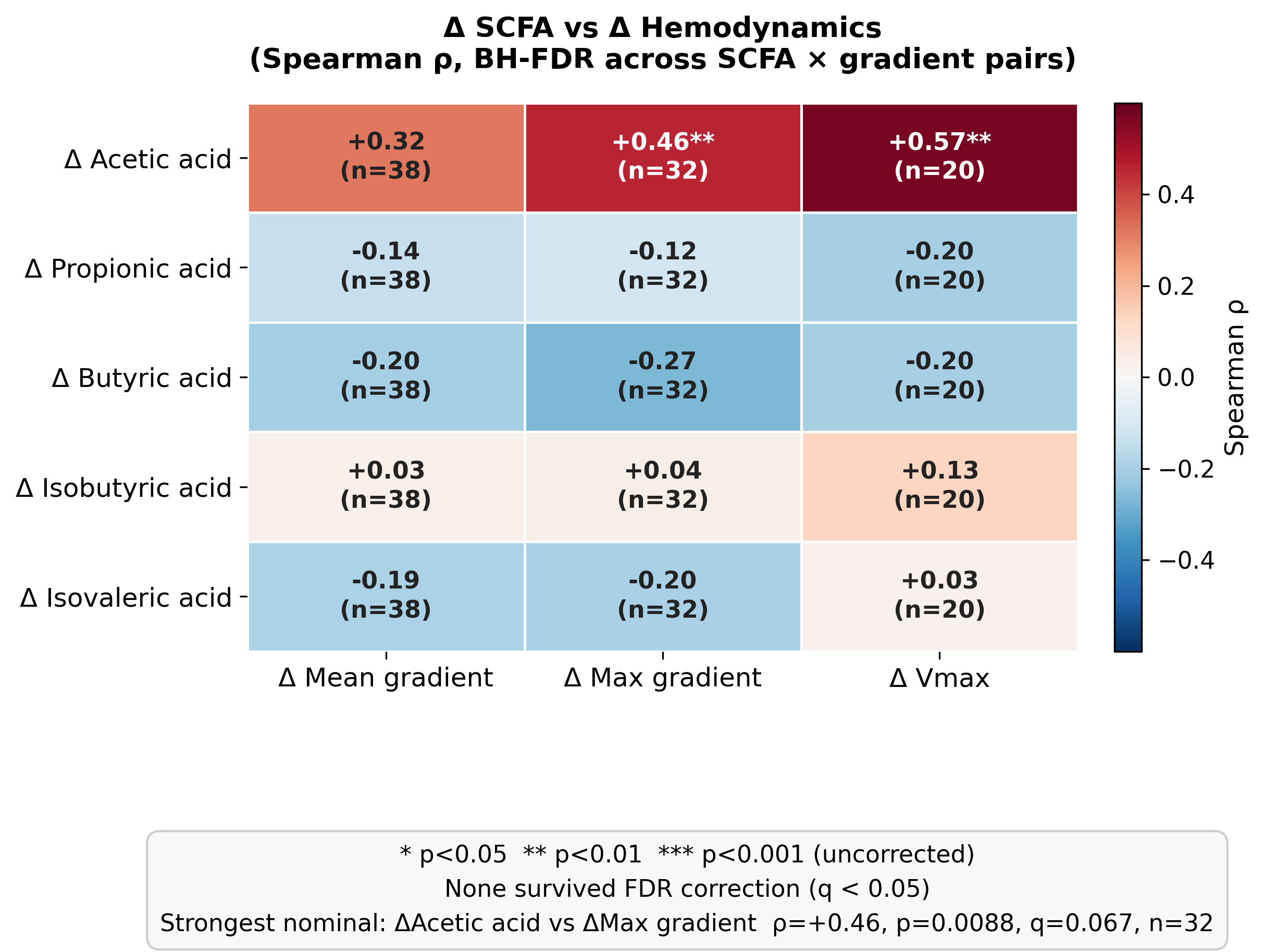

**Figure S7.** The correlation heatmap compares SCFA deltas with changes in aortic mean gradient, maximum gradient, and Vmax. No SCFA delta remained significantly associated with haemodynamic change after FDR correction.

**Figure S8. Delta-butyrate sequential association with IL-18 and IFN-γ**

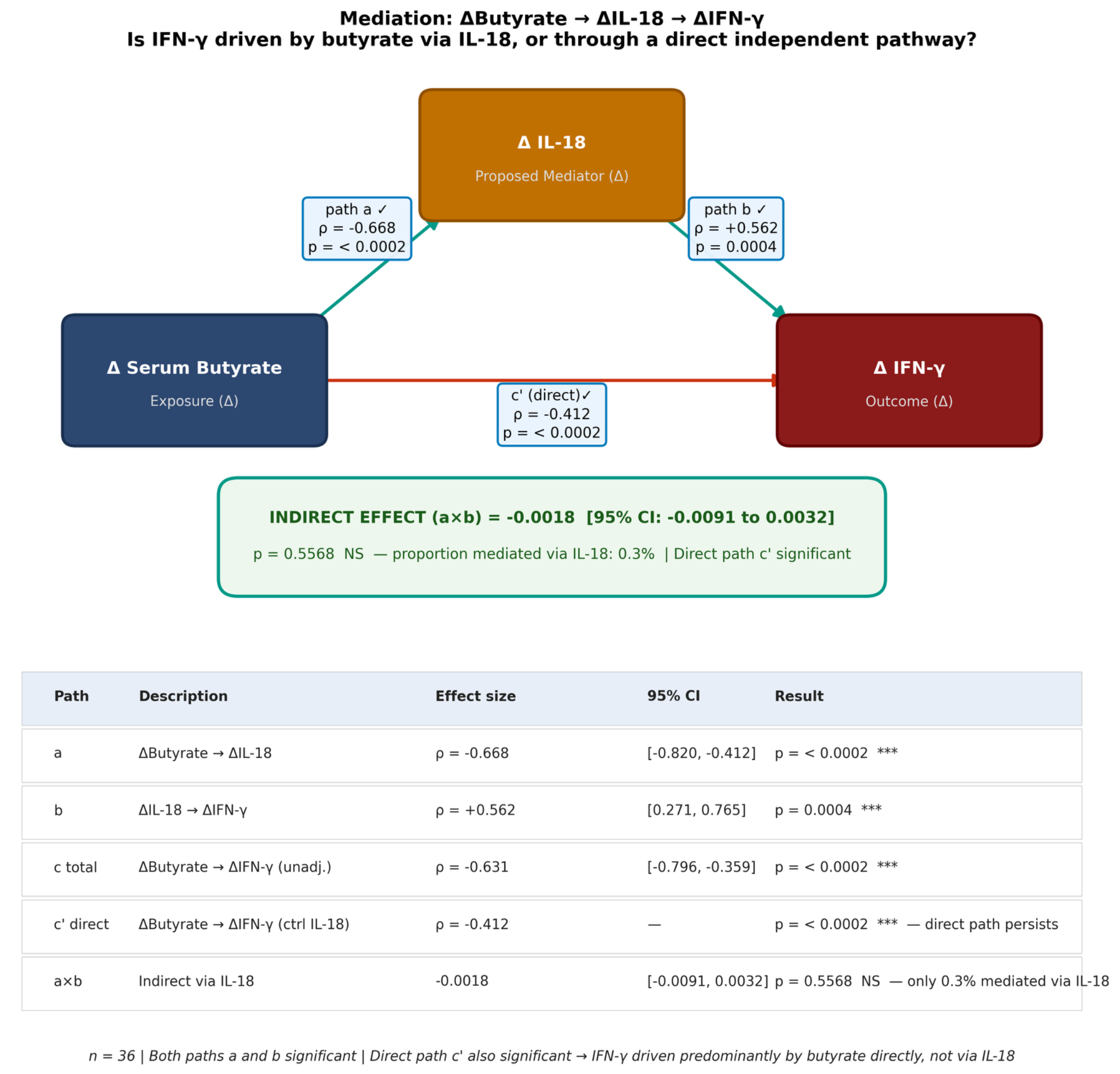

**Figure S8. The analysis shows that post-TAVI butyrate decline was also inversely associated with IFN-γ change, representing a secondary immune signal alongside the primary IL-18 association. Both IFN-γ and IL-18 were simultaneously tested as downstream variables in a three-step sequential association model; because all variables derive from the same PRE-to-POST interval, the directional interpretation remains associative.**

**Figure S9A. Inflammatory cytokine profile before and after TAVI**

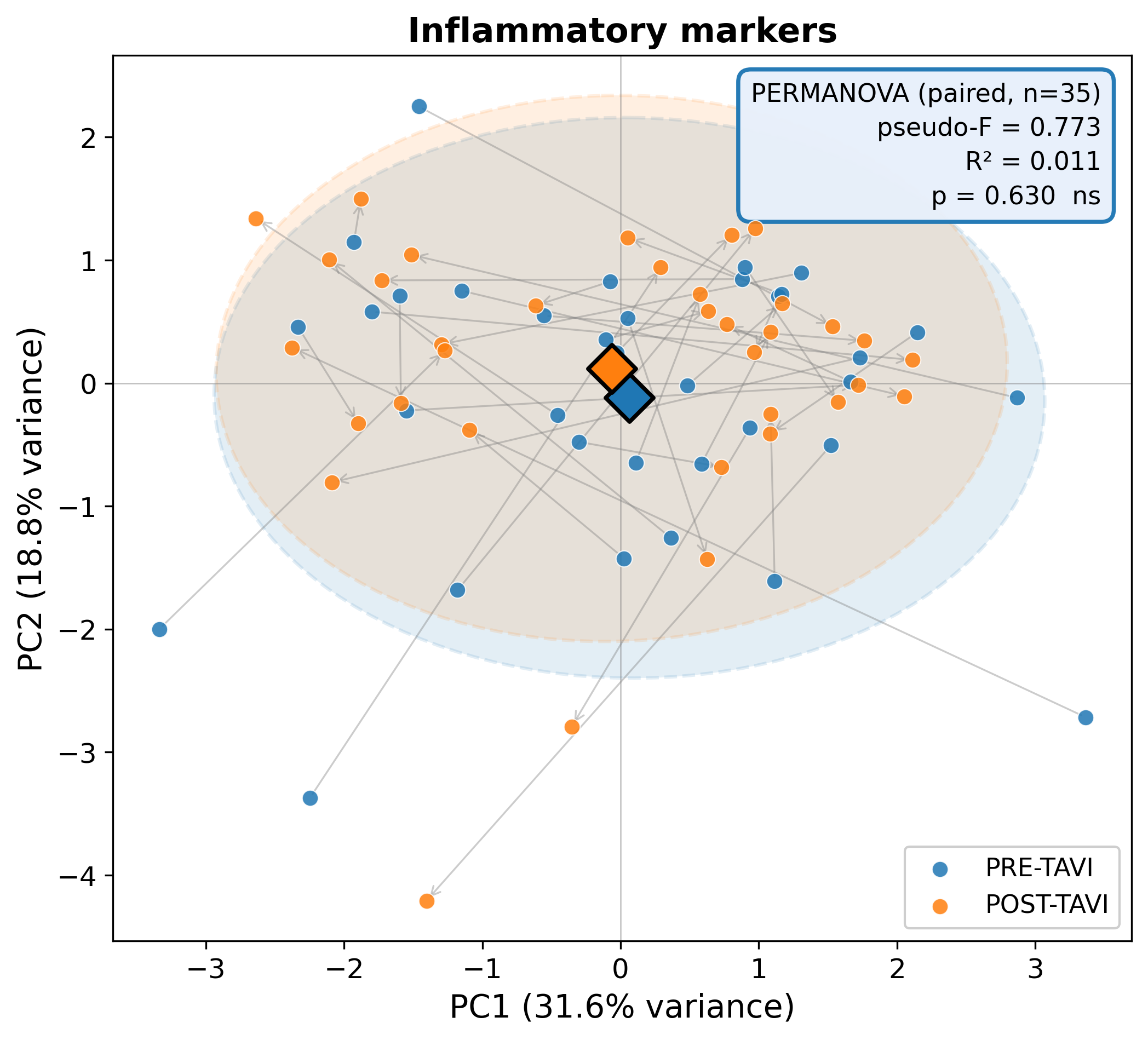

**Figure S9A.** The cytokine-panel ordination visualizes inflammatory mediator profiles before and after TAVI and supports the absence of a strong global cohort-level cytokine shift.

**Figure S9B. Delta inflammatory cytokine profile after TAVI**

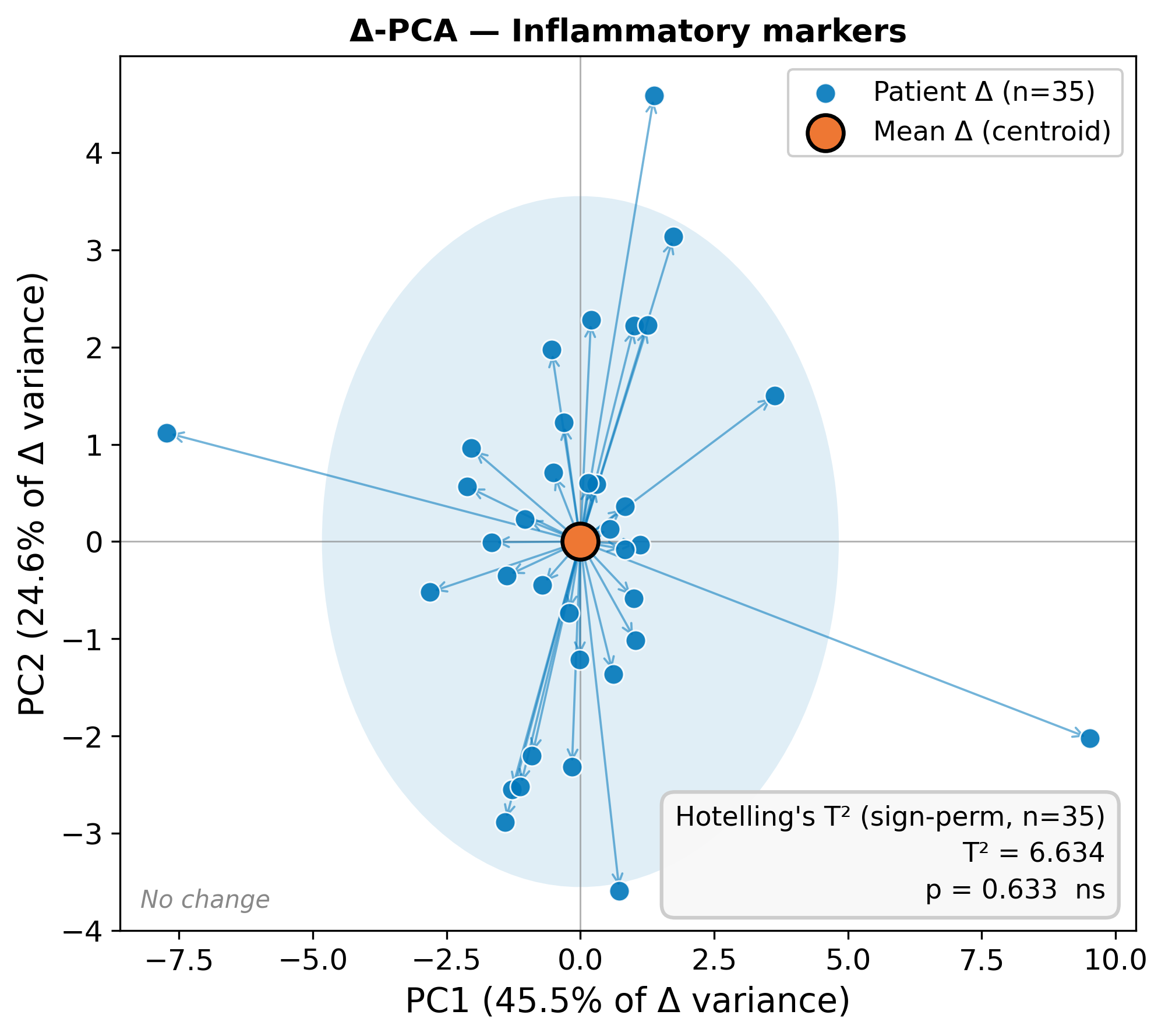

**Figure S9B.** The delta-profile analysis illustrates post-procedural cytokine variability. At the cohort level, no cytokine changed significantly after correction for multiple testing, and IL-18 remained stable on average.

**Figure S10A. Kynurenine-screen analysis for IL-18 association**

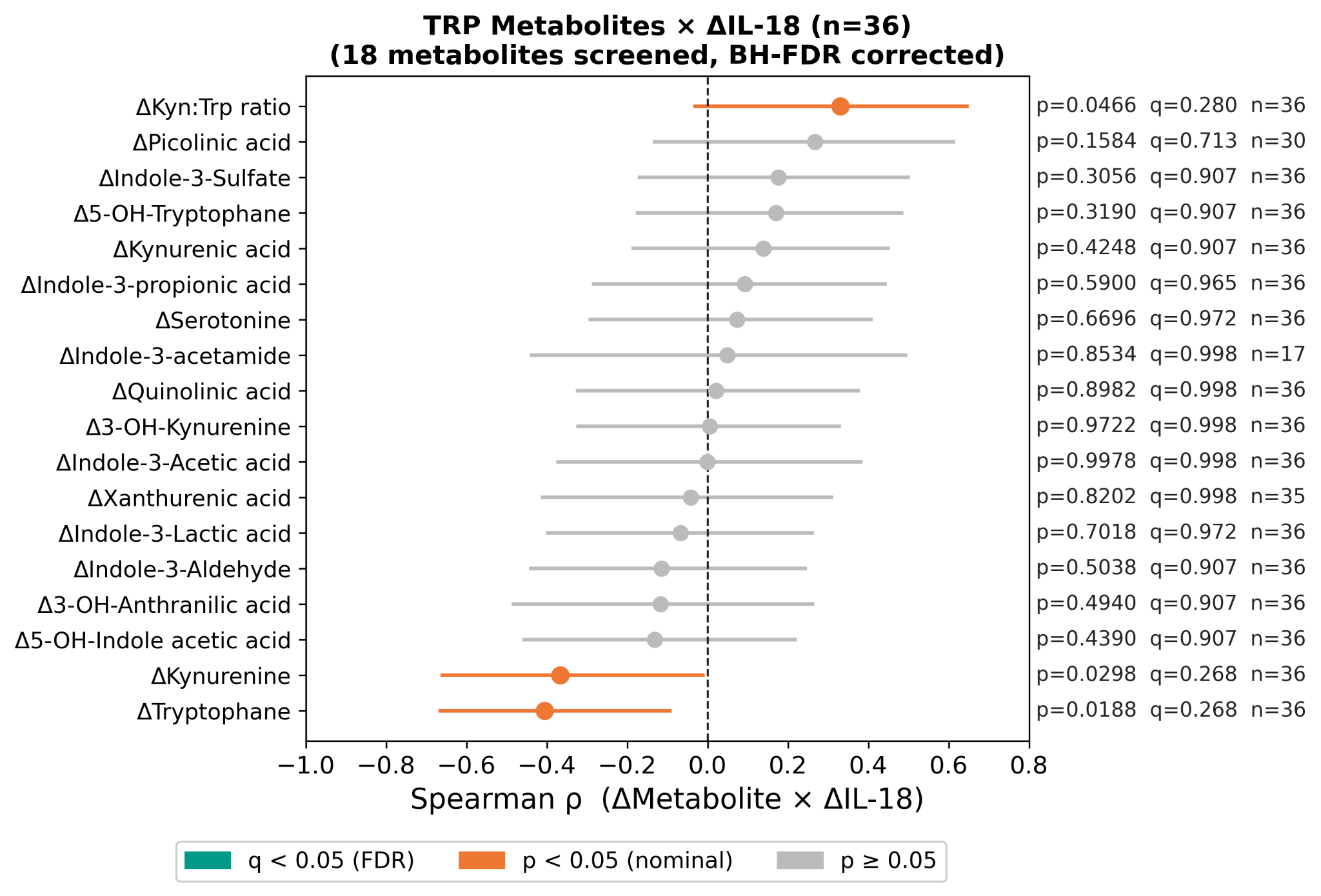

**Figure S10A.** The screen evaluates whether tryptophan-pathway changes explain IL-18 dynamics. Kynurenine did not independently account for the delta-IL-18 signal after considering butyrate.

**Figure S10B. Parallel adjustment of butyrate and kynurenine**

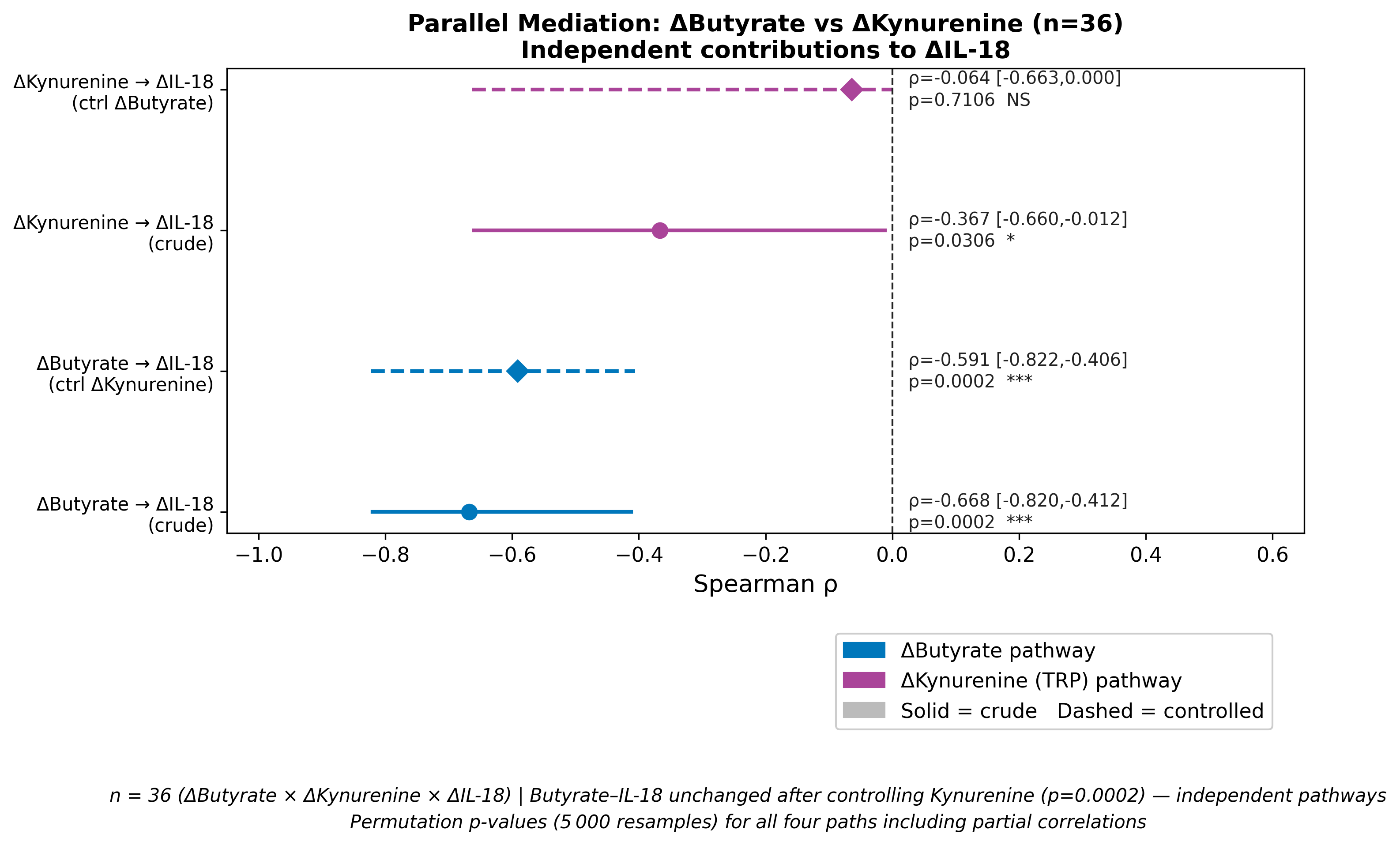

**Figure S10B. In the parallel adjustment model, delta-butyrate remains inversely associated with delta-IL-18, whereas delta-kynurenine is not independently associated after adjustment for delta-butyrate, supporting the specificity of the SCFA pathway.**

**Figure S11. Calcium-score-adjusted butyrate-IL-18 association**

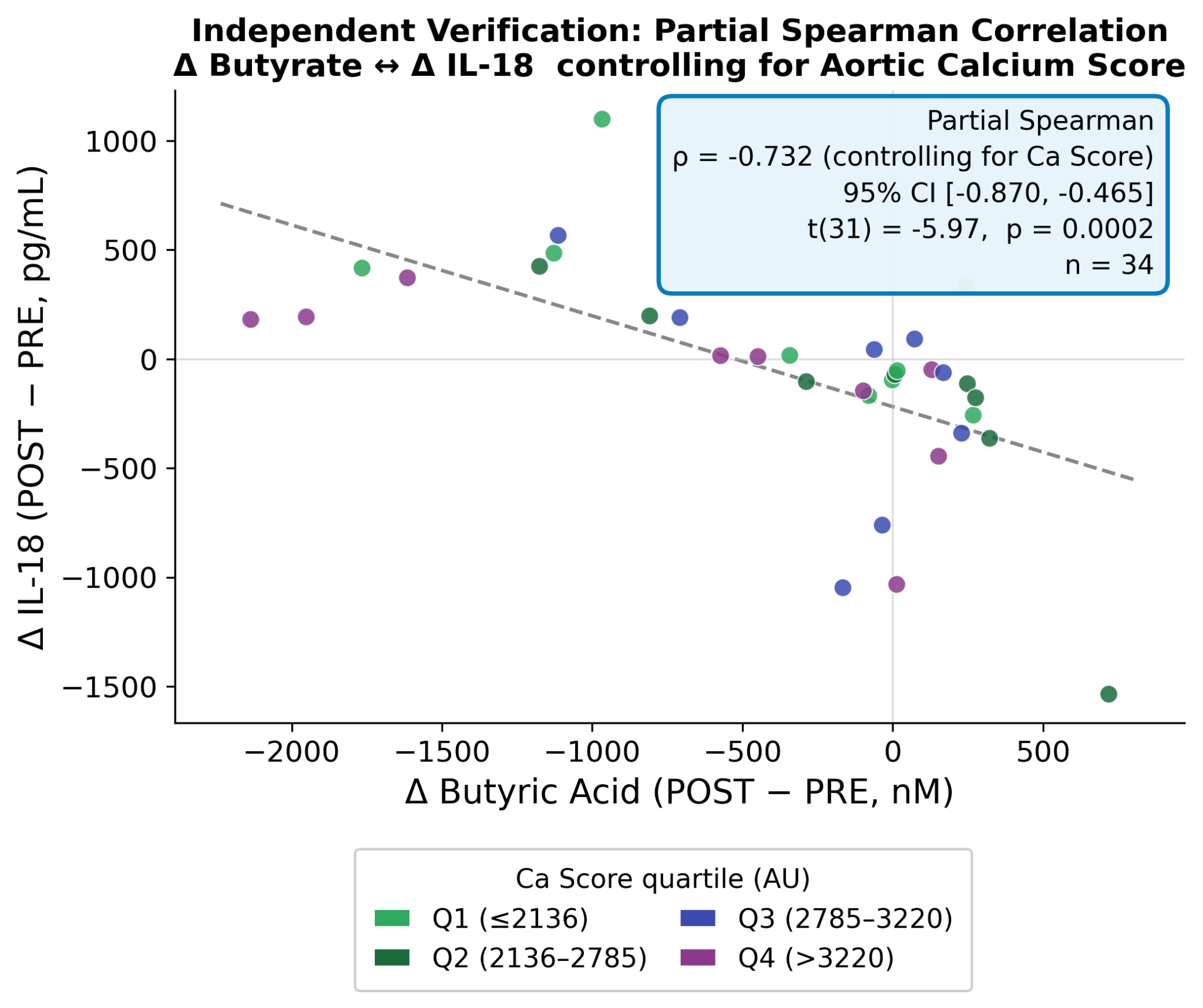

**Figure S11.** Partial Spearman analysis shows that the inverse association between delta-butyrate and delta-IL-18 persists after adjustment for aortic valve calcification burden.

**Figure S12A. Haemoglobin-decline adjustment sensitivity analysis**

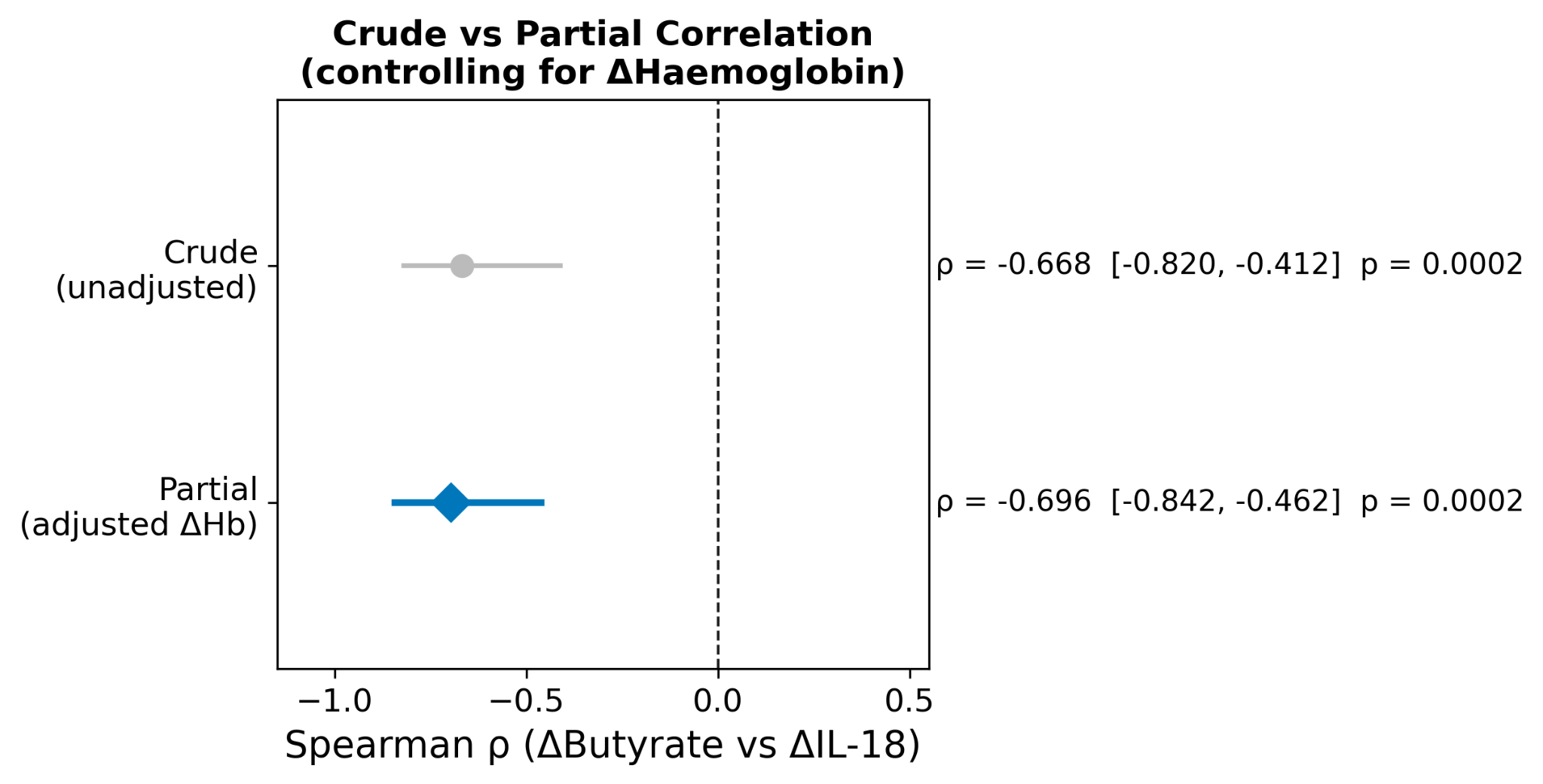

**Figure S12A.** The crude and partial estimates show that the delta-butyrate to delta-IL-18 association remains robust after accounting for peri-procedural haemoglobin decline.

**Figure S12B. High haemoglobin-drop stratum**

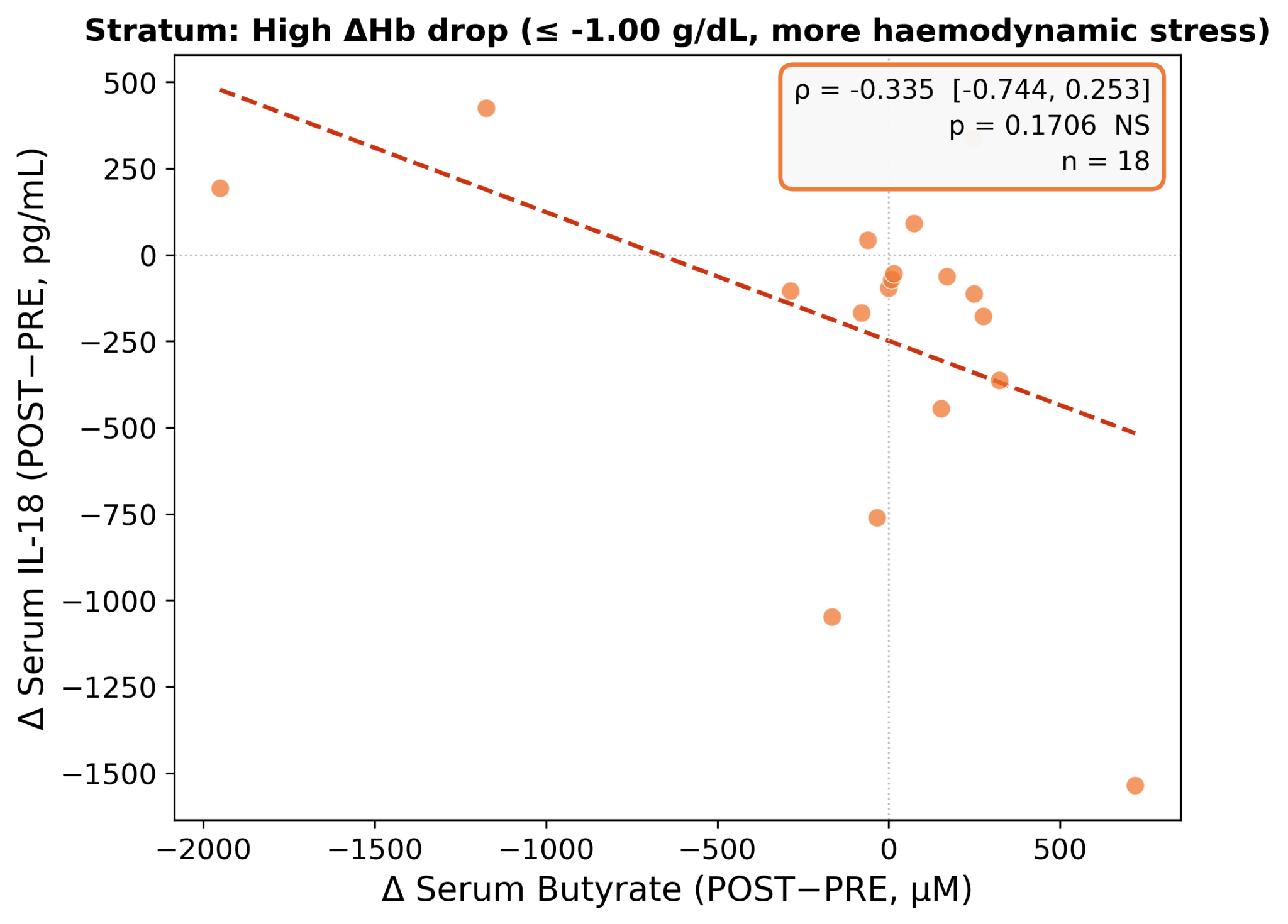

**Figure S12B.** The stratified analysis evaluates patients with greater haemoglobin decline and shows that the butyrate-IL-18 relationship is not explained by this peri-procedural stress marker.

**Figure S12C. Low haemoglobin-drop stratum**

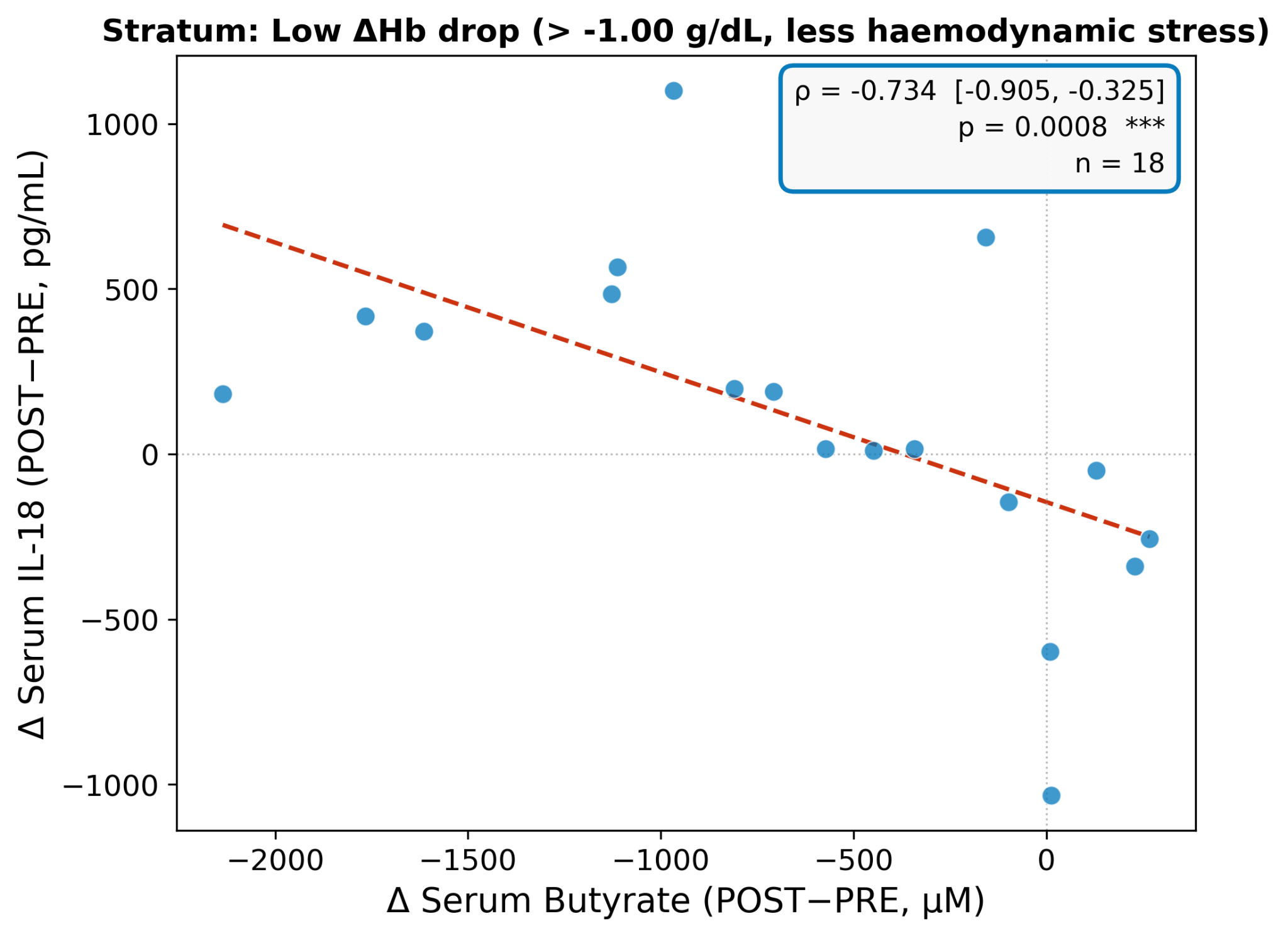

**Figure S12C.** The complementary low haemoglobin-drop stratum supports that the inverse butyrate-IL-18 association is not restricted to patients with larger haemoglobin decreases.

**Figure S13A. Medication-adjusted sensitivity analysis**

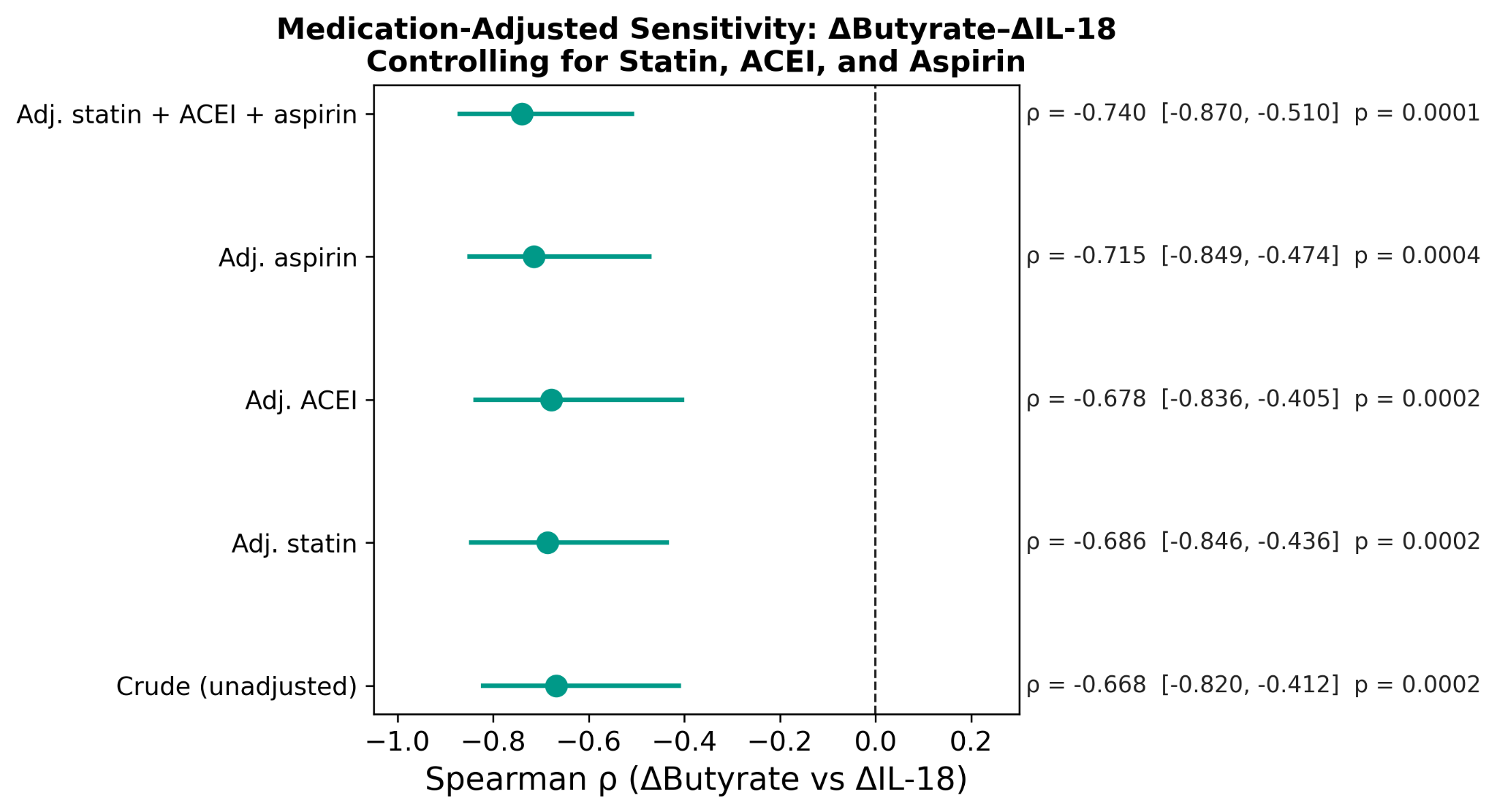

**Figure S13A.** Adjusted analyses for cardiovascular medications, including statin, ACE inhibitor, and aspirin use, preserve the inverse association between delta-butyrate and delta-IL-18.

**Figure S13B. Medication-subgroup sensitivity analysis**

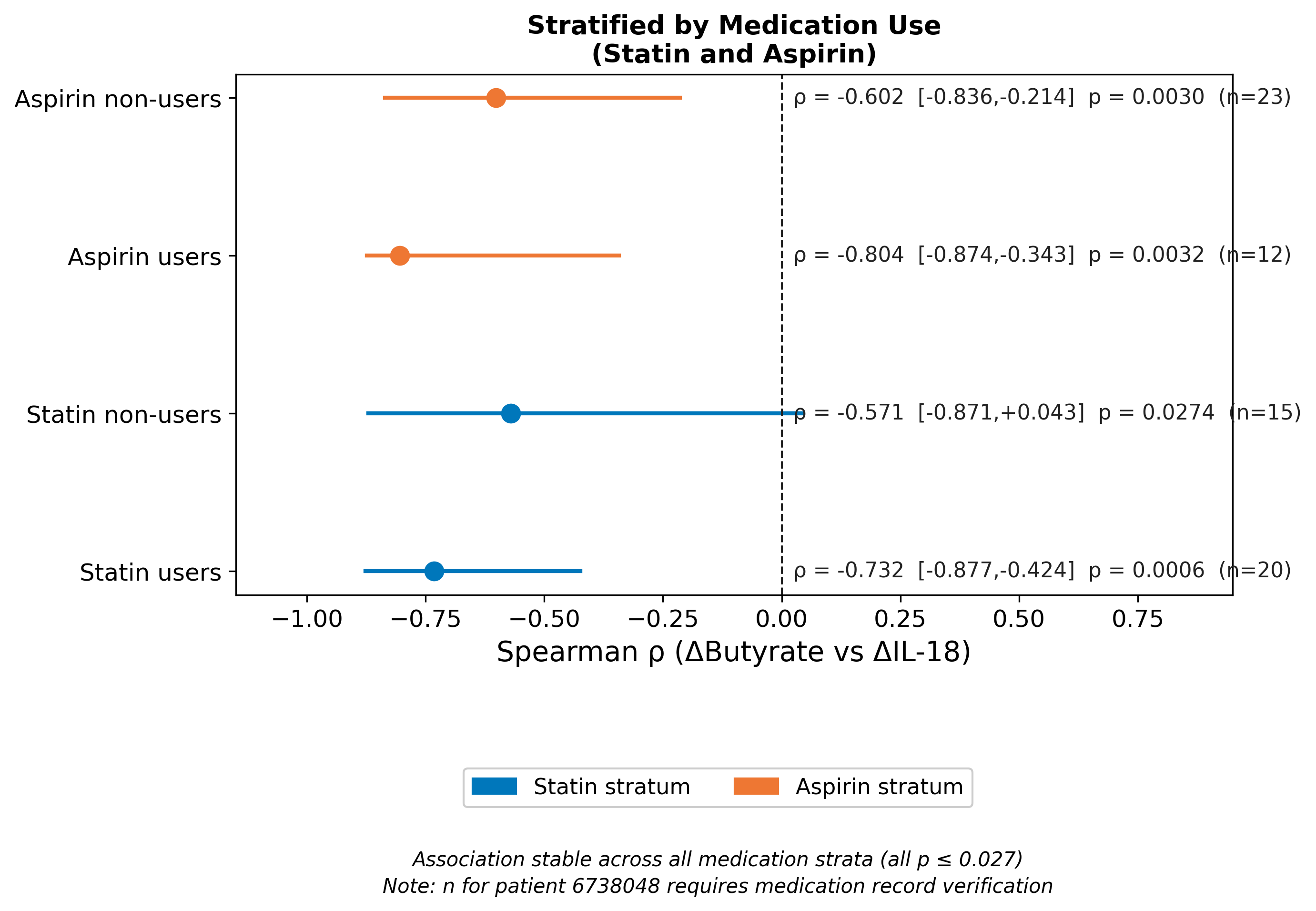

**Figure S13B.** Subgroup analyses across medication strata support that the butyrate-IL-18 axis is not driven by baseline cardiovascular medication exposure.

**Figure S14. Demographic-adjusted sensitivity analysis**

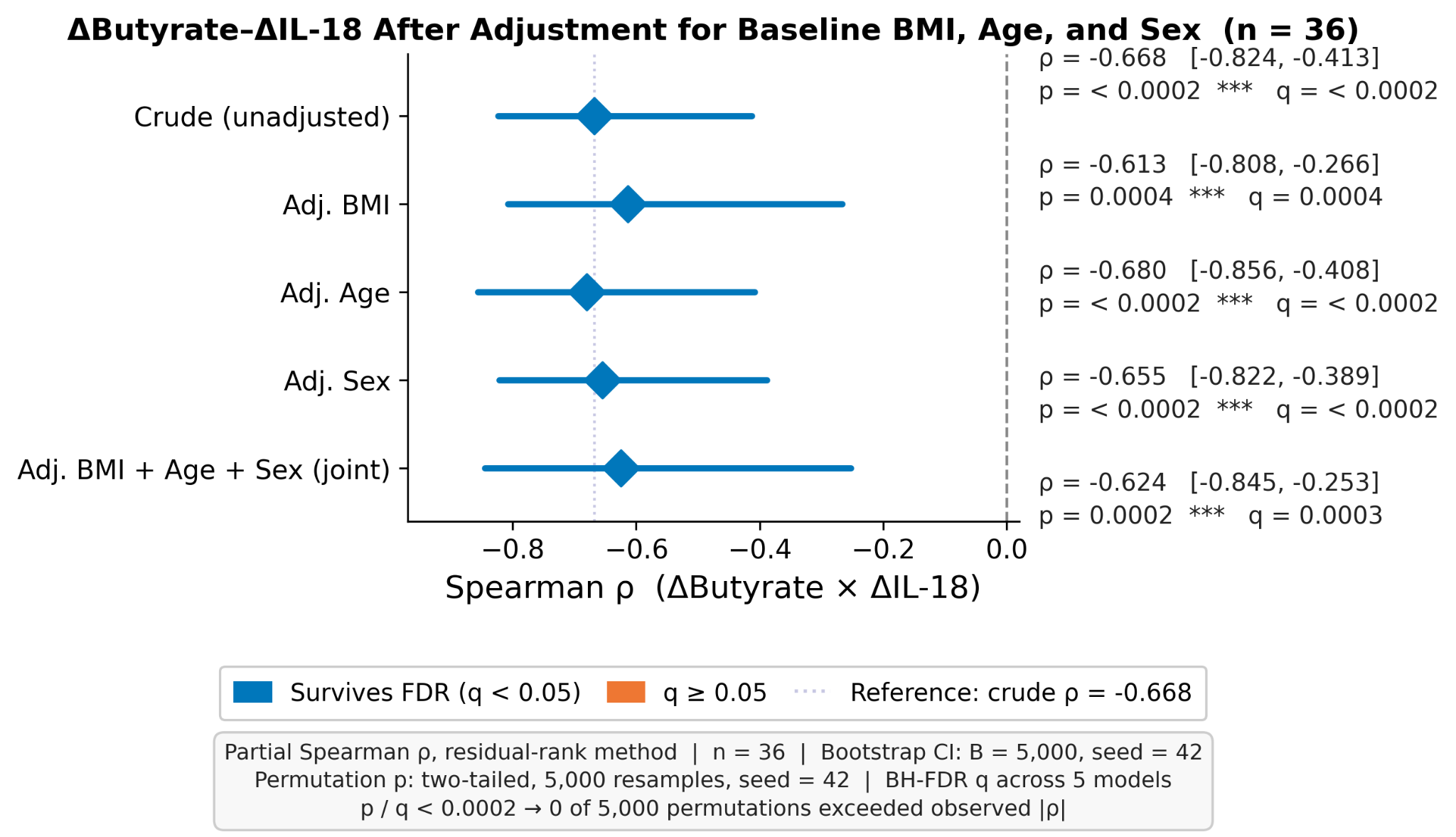

**Figure S14.** Adjustment for demographic variables including BMI, age, and sex preserves the delta-butyrate to delta-IL-18 association, supporting independence from baseline patient profile.

**Figure S15. Falsification analysis for non-mechanistic procedural markers**

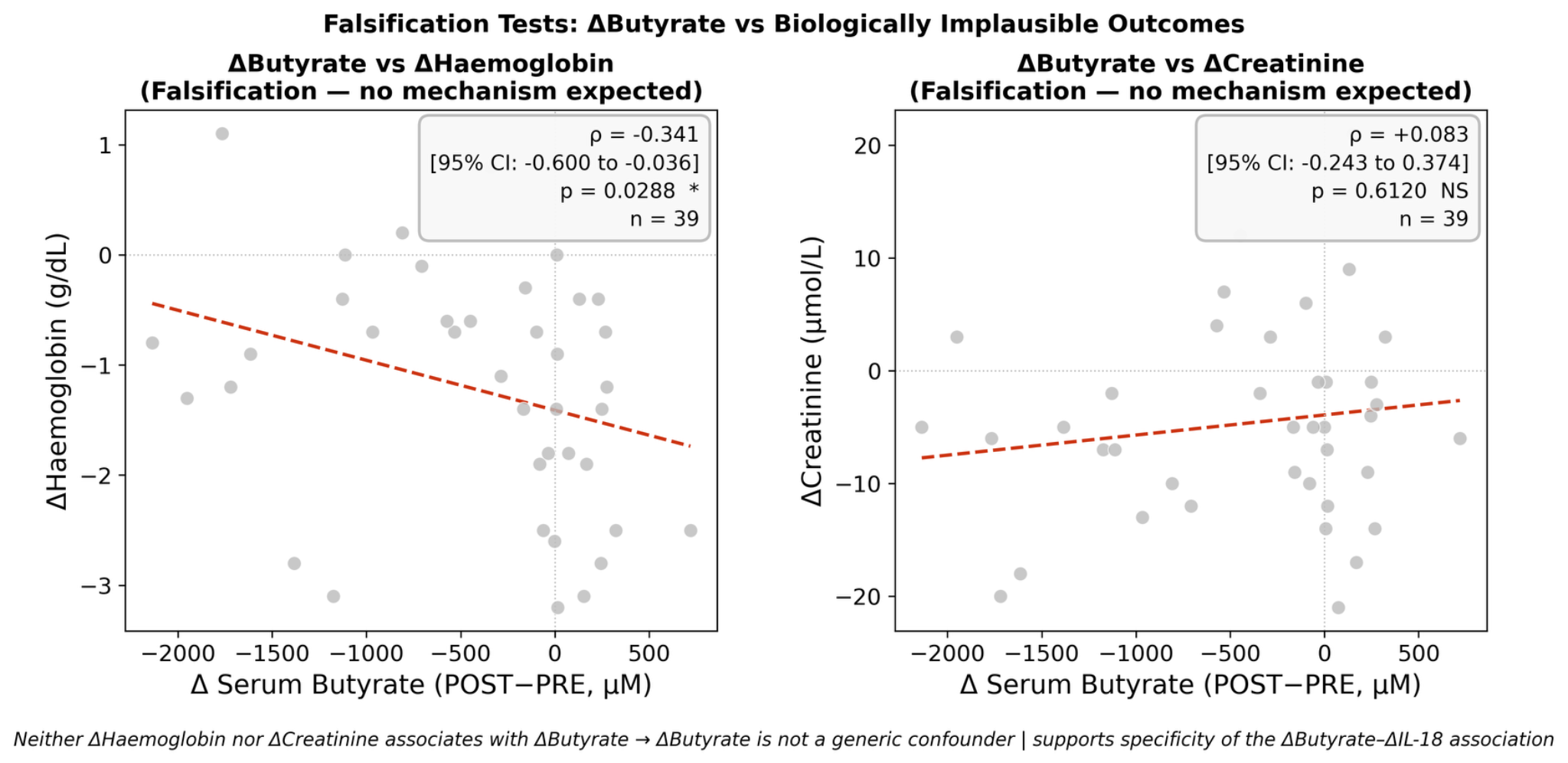

**Figure S15.** The falsification analysis shows that delta-butyrate was not associated with peri-procedural haemoglobin or creatinine changes, arguing against a non-specific procedural stress explanation.

**Figure S16. Baseline serum SCFA concentrations by 1-month adjudicated event status.**

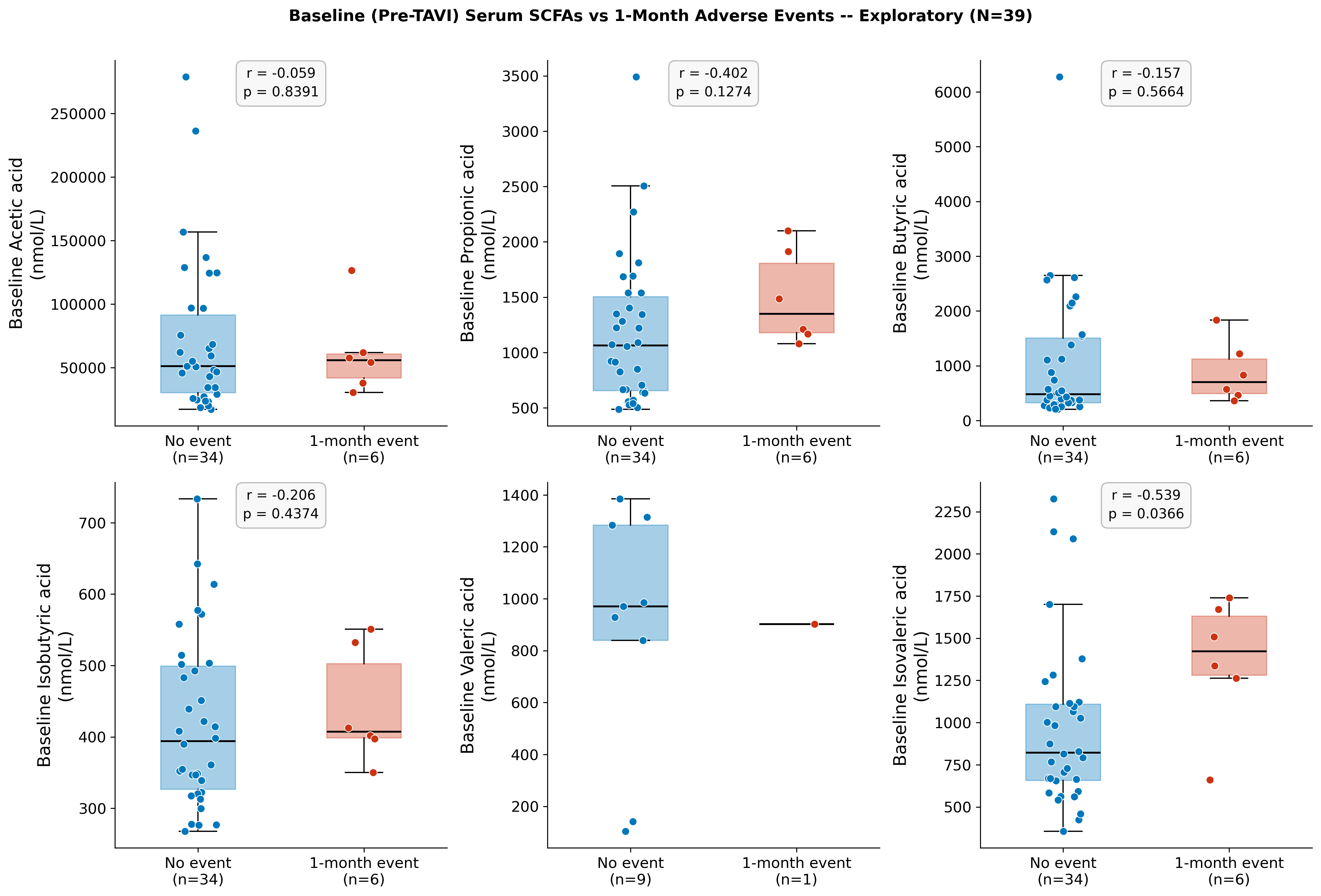

**Figure S16.** Serum concentrations of six SCFAs (acetic, propionic, isobutyric, butyric, isovaleric, and valeric acid) measured at baseline (pre-TAVI) in patients who experienced a 1-month adjudicated inflammatory or conduction event (n=6) versus those who did not (n=34). Individual data points are overlaid on box-and-whisker plots showing median, interquartile range, and 1.5×IQR whiskers. Statistical comparisons by Mann-Whitney U test, two-sided. Nominal p-values are shown without multiplicity correction (six tests, exploratory). Isovaleric acid (isovalerate) was the only SCFA with a nominal p-value below 0.05 (see Figure 5).

*SCFA, short-chain fatty acid; TAVI, transcatheter aortic valve implantation; IQR, interquartile range; nM, nanomolar.*

**Figure S17. Baseline serum isovalerate concentration and 1-month adjudicated adverse events.**

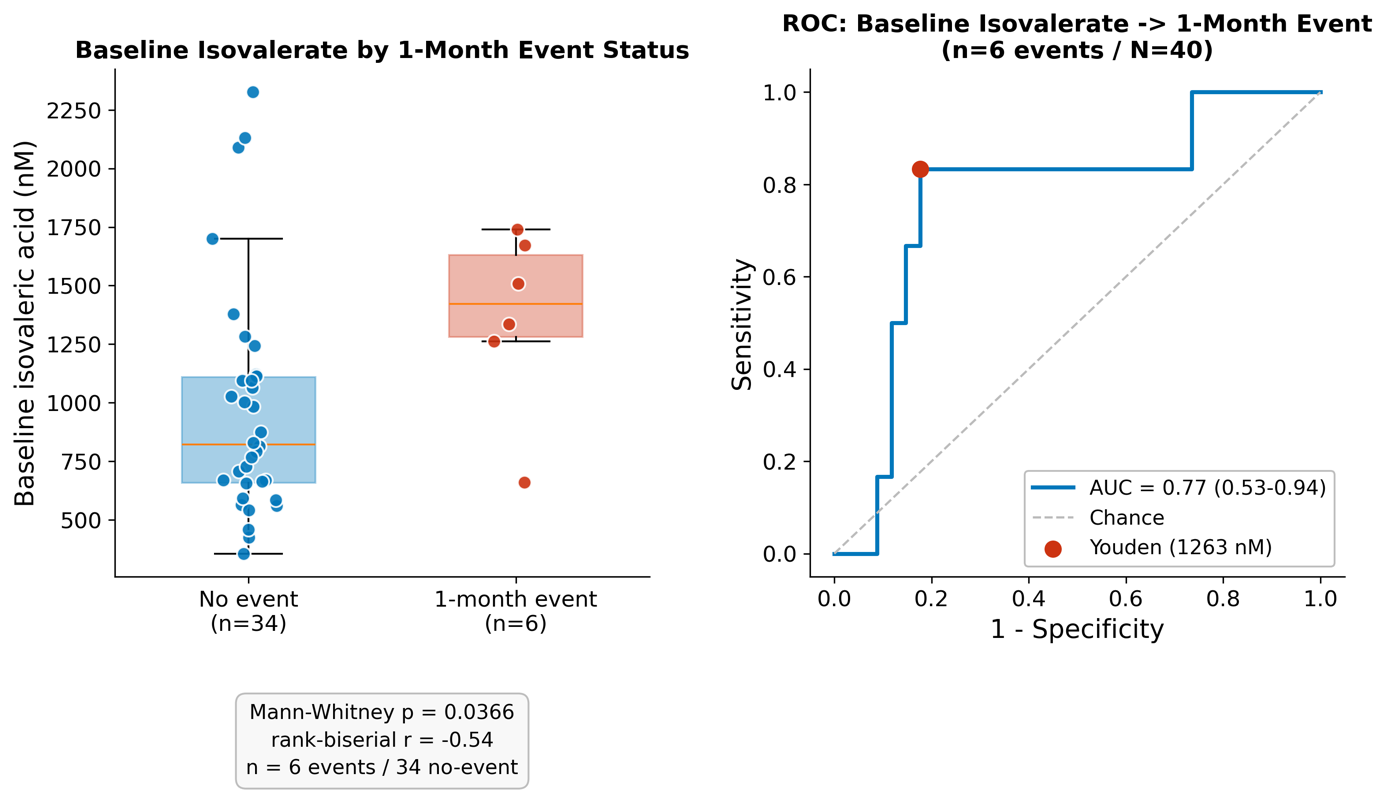

**Figure S17.** Left panel: Serum isovalerate concentrations measured at baseline (pre-TAVI) in patients who subsequently experienced a 1-month adjudicated inflammatory or conduction event (n=6) versus those who did not (n=34). Individual data points are overlaid on box-and-whisker plots showing median, interquartile range, and 1.5×IQR whiskers. Statistical comparison by Mann-Whitney U test, two-sided (p=0.037; rank-biserial r=−0.54). Right panel: Receiver operating characteristic (ROC) curve for baseline serum isovalerate as a predictor of 1-month adjudicated event status. The area under the curve (AUC) was estimated by bootstrap resampling (B=5,000, stratified by outcome; 95% bootstrap CI 0.53-0.94). The Youden-optimal classification threshold (1,263 nM; sensitivity 0.83, specificity 0.82) is indicated as a filled circle. This analysis is exploratory and based on 6 events; the association should be regarded as hypothesis-generating and requires prospective validation. TAVI, transcatheter aortic valve implantation; SCFA, short-chain fatty acid; AUC, area under the curve; ROC, receiver operating characteristic; nM, nanomolar; IQR, interquartile range.

**Figure S18. Inflammatory adverse event rates at 3-month follow-up by SCFA Responder status.**

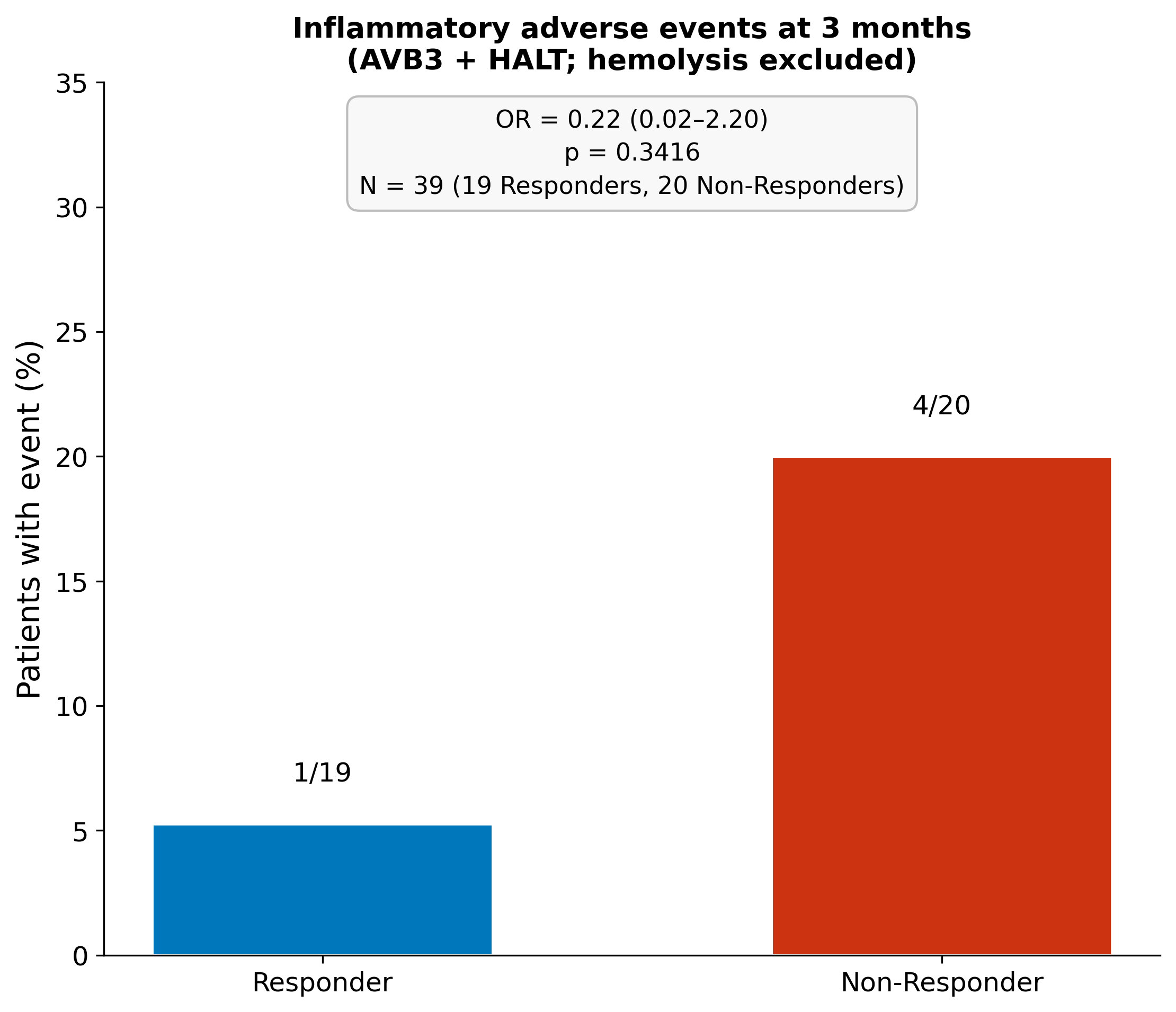

**Figure S18.** Inflammatory adverse event rates at 3-month follow-up in SCFA Responders (n=19) and Non-Responders (n=20). SCFA Responders were defined as patients with a composite standardised z-score above zero for the pre-to-post TAVI change in butyrate and isovalerate; Non-Responders had a composite z-score at or below zero. One patient without a post-TAVI SCFA sample was excluded (total N=39). The inflammatory composite comprised complete atrioventricular block (AVB3) and haemodynamic leaflet thrombosis (HALT); haemolysis was excluded. p-value from Fisher’s exact test, two-sided. Odds ratio and 95% confidence interval were computed with Haldane–Anscombe continuity correction (+0.5 to all cells) because one cell contained zero observations; p-value from the uncorrected exact test. This comparison is exploratory; no multiplicity correction was applied.

*SCFA, short-chain fatty acid; TAVI, transcatheter aortic valve implantation; AVB3, complete atrioventricular block; HALT, haemodynamic leaflet thrombosis; OR, odds ratio; CI, confidence interval.*
