## Supplementary material for "Gut microbiome-derived metabolic remodeling and the butyrate–IL-18 inflammatory axis after transcatheter aortic valve implantation": Main Tables

**TABLE 1: Baseline Characteristics**

| **Characteristic** | **N** | **Value** |
| --- | --- | --- |
| **Demographics** | | |
| Age, years | 40 | 81 [78.5–85.3] |
| Female sex | 40 | 21 (52.5%) |
| Body mass index, kg/m² | 40 | 26.1 [23.1–29.1] |
| **Symptoms** | | |
| NYHA functional class I | 40 | 13 (32.5%) |
| NYHA functional class II | 40 | 22 (55.0%) |
| NYHA functional class III | 40 | 4 (10.0%) |
| NYHA functional class IV | 40 | 1 (2.5%) |
| Angina (CCS ≥ I) | 40 | 7 (17.5%) |
| Dyspnoea | 40 | 28 (70.0%) |
| Syncope | 40 | 3 (7.5%) |
| Acute heart failure | 40 | 0 (0.0%) |
| Hospitalisation for heart failure | 40 | 0 (0.0%) |
| **Cardiovascular history** | | |
| Hypertension | 40 | 33 (82.5%) |
| Dyslipidaemia | 40 | 21 (52.5%) |
| Diabetes mellitus | 40 | 9 (22.5%) |
| – Insulin-dependent diabetes | 40 | 2 (5.0%) |
| – Diabetes with complications | 40 | 1 (2.5%) |
| Current smoker | 40 | 4 (10.0%) |
| Former smoker | 40 | 8 (20.0%) |
| Coronary artery disease | 40 | 8 (20.0%) |
| Prior PCI | 40 | 9 (22.5%) |
| Prior myocardial infarction | 40 | 3 (7.5%) |
| Myocardial infarction within 90 days | 40 | 1 (2.5%) |
| Atrial fibrillation | 40 | 3 (7.5%) |
| Permanent pacemaker | 40 | 8 (20.0%) |
| Prior stroke/TIA | 40 | 5 (12.5%) |
| Peripheral artery disease | 40 | 1 (2.5%) |
| Porcelain aorta | 40 | 1 (2.5%) |
| Active/prior cancer | 40 | 6 (15.0%) |
| COPD | 40 | 3 (7.5%) |
| Renal failure | 40 | 2 (5.0%) |
| Dialysis | 40 | 0 (0.0%) |
| Elective procedure | 40 | 40 (100.0%) |
| **Pre-procedural medications** | | |
| Aspirin | 40 | 13 (32.5%) |
| Clopidogrel | 40 | 5 (12.5%) |
| Vitamin K antagonist | 40 | 2 (5.0%) |
| Statin | 40 | 20 (50.0%) |
| ACEI | 40 | 7 (17.5%) |
| ARB | 40 | 18 (45.0%) |
| Beta-blocker | 40 | 19 (47.5%) |
| Diuretics | 40 | 19 (47.5%) |
| **Pre-procedural vital signs** | | |
| Systolic blood pressure, mmHg | 40 | 133 [123–146.5] |
| Diastolic blood pressure, mmHg | 40 | 72 [62.8–84.0] |
| Heart rate, bpm | 40 | 70.5 [65–82.3] |
| **Pre-procedural laboratory values** | | |
| Haemoglobin, g/dL | 40 | 12.95 [12.2–14.0] |
| Leucocytes, ×10⁹/L | 40 | 7.26 [6.09–8.41] |
| Platelets, ×10⁹/L | 40 | 224 [163–246] |
| Creatinine, µmol/L | 40 | 84.5 [70.8–94.0] |
| eGFR (CKD-EPI), mL/min/1.73 m² | 40 | 60.3 [50.4–69.3] |
| Sodium, mmol/L | 31 | 140 [139–141] |
| Potassium, mmol/L | 30 | 4.05 [3.90–4.20] |
| BNP, pg/mL | 27 | 1170 [366–2015] |
| CK, U/L | 40 | 97 [66.8–126.3] |
| CK-MB, µg/L | 22 | 3.30 [2.20–4.92] |
| Albumin, g/L | 18 | 35 [34–37] |
| CRP, mg/L | 23 | 1.00 [1.00–3.00] |
| Fasting glucose, g/L | 39 | 0.99 [0.92–1.29] |
| HbA1c, % | 22 | 5.90 [5.50–6.20] |
| Total cholesterol, mmol/L | 34 | 4.06 [3.07–5.07] |
| LDL cholesterol, mmol/L | 34 | 1.92 [1.36–3.25] |
| HDL cholesterol, mmol/L | 34 | 1.29 [1.12–1.69] |
| Triglycerides, mmol/L | 34 | 1.20 [1.03–1.46] |
| Lipoprotein(a), nmol/L | 37 | 20 [20–106] |
| **Echocardiography** | | |
| LVEF, % | 40 | 60.5 [54.8–66.8] |
| Mean aortic gradient, mmHg | 40 | 40.0 [32.3–47.0] |
| Peak (max) aortic gradient, mmHg | 33 | 60.0 [43.0–70.0] |
| Peak aortic velocity (Vmax), m/s | 21 | 4.00 [3.60–4.11] |
| Aortic valve area, cm² | 40 | 0.80 [0.70–0.98] |
| Aortic valve area index, cm²/m² | 40 | 0.43 [0.37–0.52] |
| Aortic valve calcification grade | 25 | Severe: 20/25 (80.0%) |
| Aortic valve morphology | 40 | Tricuspid: 26 (65.0%); native unknown: 14 (35.0%) |
| Mitral regurgitation | 37 | Trace/mild: 22 (59.5%) |
| Tricuspid regurgitation | 37 | Trace/mild: 26 (70.3%) |
| Right ventricular function | 38 | Normal: 33/38 (86.8%) |
| PASP, mmHg | 5 | 40 [31–45] |
| **CT — aortic valve calcium score** | | |
| Aortic valve calcium score, AU | 38 | 2819 [2129–3342] |
| Calcium semiquantitative grade | 33 | Severe (grade 3): 29/33 (87.9%) |
| **Surgical risk scores** | | |
| EuroSCORE II, % | 39 | 1.9 (1.5–3.3) |
| STS predicted mortality, % | 39 | 2.8 (2.0–4.1) |

**Table 1. Baseline characteristics of the GUT-TAVI cohort.**

Values are median [interquartile range] or n (%). N denotes valid observations. Rows with N < 40 reflect available data. **Abbreviations:** ACEI = angiotensin-converting enzyme inhibitor; ARB = angiotensin receptor blocker; AU = Agatston units; AV = aortic valve; BNP = B-type natriuretic peptide; CCS = Canadian Cardiovascular Society; CK = creatine kinase; COPD = chronic obstructive pulmonary disease; CRP = C-reactive protein; CT = computed tomography; eGFR = estimated glomerular filtration rate; HbA1c = glycated haemoglobin; HDL = high-density lipoprotein; IQR = interquartile range; LDL = low-density lipoprotein; LVEF = left ventricular ejection fraction; MI = myocardial infarction; NYHA = New York Heart Association; PASP = pulmonary artery systolic pressure; PCI = percutaneous coronary intervention; RV = right ventricular; STS = Society of Thoracic Surgeons; TIA = transient ischaemic attack.

**Table 2. Post-procedural adverse events across four follow-up windows.**

| **Event** | **Periprocedural (N=40) N (%)** | **1-month (N=40) N (%)** | **3-month (N=40) N (%)** | **1-year (N=38) N (%)** |
| --- | --- | --- | --- | --- |
| **All-cause mortality** | 0 (0.0%) | 0 (0.0%) | 0 (0.0%) | 0 (0.0%) |
| **Any stroke** | 1 (2.5%) | 0 (0.0%) | 0 (0.0%) | 0 (0.0%) |
| Disabling stroke | 0 (0.0%) | 0 (0.0%) | 0 (0.0%) | 0 (0.0%) |
| Non-disabling stroke | 1 (2.5%) | 0 (0.0%) | 0 (0.0%) | 0 (0.0%) |
| **Myocardial infarction** | 0 (0.0%) | 0 (0.0%) | 0 (0.0%) | 1 (2.6%) |
| **Major or life-threatening bleeding** | 0 (0.0%) | 0 (0.0%) | 0 (0.0%) | 1 (2.6%) |
| **Major vascular complication** | 4 (10.0%) | 1 (2.5%) | 0 (0.0%) | 0 (0.0%) |
| **Unplanned aortic valve reintervention** | 0 (0.0%) | 0 (0.0%) | 0 (0.0%) | 1 (2.6%) |
| **New permanent pacemaker implantation** | 4 (10.0%) | 0 (0.0%) | 0 (0.0%) | 1 (2.6%) |
| **New left bundle branch block** | 0 (0.0%) | 3 (7.5%) | 0 (0.0%) | 3 (7.9%) |
| **New right bundle branch block** | 0 (0.0%) | 0 (0.0%) | 0 (0.0%) | 4 (10.5%) |
| **First-degree atrioventricular block** | 0 (0.0%) | 1 (2.5%) | 0 (0.0%) | 3 (7.9%) |
| **Second-degree atrioventricular block** | 0 (0.0%) | 0 (0.0%) | 1 (2.5%) | — |
| **Third-degree atrioventricular block** | 0 (0.0%) | 0 (0.0%) | 1 (2.5%) | 0 (0.0%) |
| **New-onset atrial fibrillation** | 0 (0.0%) | 1 (2.5%) | 0 (0.0%) | 1 (2.6%) |
| **Hypoattenuated leaflet thickening** | 0 (0.0%) | 0 (0.0%) | 4 (10.0%) | 0 (0.0%) |
| **Hypoattenuated leaflet thickening with reduced leaflet motion** | 0 (0.0%) | 0 (0.0%) | 1 (2.5%) | 0 (0.0%) |
| **Hemolysis** | 0 (0.0%) | 0 (0.0%) | 1 (2.5%) | 0 (0.0%) |
| **COMPOSITE ENDPOINTS** |  |  |  |  |
| **VARC-3 Technical Success †** | 37 (92.5%) | — | — | — |
| **VARC-3 Early Safety (≤30 days) ‡** | 5 (12.5%) | 1 (2.5%) | — | — |
| **VARC-3 Clinical Efficacy (1-year) §** | — | — | — | 1 (2.6%) |
| **Any inflammatory event** | 8 (20.0%) | 6 (15.0%) | 5 (12.5%) | 11 (28.9%) |

**Table 2.** Adjudicated events (VARC-3) were obtained from the SwissTAVI adjudicated events database; all other events are from discharge forms, 30-day follow-up CRF, 3-month visit data, and SwissTAVI 1-year registry. p-values from Fisher's exact test, two-sided. Odds ratios and 95% confidence intervals for cells with zero observations computed with Haldane-Anscombe continuity correction; all comparisons are exploratory.

† VARC-3 Technical Success assessed at procedure exit using discharge echocardiography (N=40).

§ Partial composite only — registry does not include KCCQ/NYHA or hospitalisation data. Components available: all-cause mortality (0), all stroke (0), and unplanned aortic valve reintervention (1).

*Abbreviations: SCFA, short-chain fatty acid; PPM, permanent pacemaker; SVD, structural valve deterioration; LBBB, left bundle branch block; RBBB, right bundle branch block; AVB, atrioventricular block; AF, atrial fibrillation; HALT, haemodynamic leaflet thrombosis; SAE, serious adverse event; OR, odds ratio; CI, confidence interval; n/a, not applicable; VARC, Valve Academic Research Consortium.*
