## Supplementary data metagenomics for "Gut microbiome-derived metabolic remodeling and the butyrate–IL-18 inflammatory axis after transcatheter aortic valve implantation"

**Metabolomics detailed methods**

**Bile acids**(1)

Serum samples were thawed at room temperature and vortexed before processing. For absolute quantification of bile acids, 50 µL of serum, calibration standards, and quality controls prepared in methanol/water (1:1) were mixed with 400 µL of internal standard solution in isopropanol in a 96-deepwell plate. After incubation at −20 °C for 30 min, samples were centrifuged at 4 °C (3000 rpm, 30 min). A volume of 350 µL of supernatant was transferred and evaporated under nitrogen at 40 °C, then reconstituted in 100 µL methanol/water (1:1), centrifuged again, and 80 µL transferred to a 96-well plate for LC-MS/MS injection (5 µL). For highly concentrated samples, an additional 1:10 dilution step was performed prior to extraction. Chromatographic separation was achieved on a UPLC I-Class system using an Acquity BEH C8 column, and detection was performed on a Xevo TQ-XS mass spectrometer in MRM mode with ESI ±. Quantification was based on metabolite-to–internal standard peak area ratios using external calibration curves, validated by quality controls and expressed within the established LLOQ–ULOQ ranges.

**Short-chain fatty acids**(2)

SCFAs were quantified in serum using a derivatization-based LC-MS/MS method adapted from Monteiro et al. Briefly, 50 µL of serum was mixed with 150 µL methanol, shaken, incubated at −20 °C, and centrifuged. An aliquot of supernatant (or a 1:10 diluted extract for highly concentrated samples) was subjected to derivatization with internal standards, pyridine, EDC, and 3-nitrophenylhydrazine, followed by incubation for 1 h at 4 °C. The reaction was quenched with formic acid–containing water and the plate was shaken prior to LC-MS/MS analysis (15 µL injection). Separation was performed on a C18 column using a water/acetonitrile gradient with formic acid, and detection was carried out on a Xevo TQ-XS in negative ESI MRM mode. Concentrations were calculated from calibration curves built on metabolite-to–internal standard response ratios and expressed in nM, with batch validity ensured by quality control samples within predefined acceptance limits. Values below the lower limit of quantification were imputed as LLOQ/√2. Valerate was excluded from downstream analyses as pre-specified because 84.6% of measurements were below the lower limit of quantification. Complete paired SCFA metabolomics data were available for 39 patients.

**TMA, TMAO, choline, and L-carnitine**(3)

Serum levels of TMA, TMAO, choline, and L-carnitine were measured after ethyl bromoacetate derivatization. Fifty microliters of serum, calibration standards, and quality controls were spiked with internal standards, derivatized with ethyl bromoacetate, and incubated at room temperature under agitation. The reaction was quenched with acetonitrile/water/formic acid, followed by centrifugation, and 2 µL of supernatant were injected into the LC-MS/MS system. Chromatographic separation was performed on a Cortecs HILIC column using ammonium formate/formic acid buffers, and detection was achieved on a Xevo TQ-XS in positive ESI MRM mode. Absolute concentrations were obtained from external calibration curves using metabolite-to–internal standard response ratios, with quality controls used to validate analytical performance.

**Tryptophan metabolites**(4)

Samples were allowed to return to room temperature and vortexed prior to processing. Calibration standards and quality control samples were prepared in PBS containing 4% (w/v) BSA. For extraction, 200 µL of internal standard solution in methanol were added to 50 µL of sample, calibration standard, or quality control in a 96-deepwell plate. Plates were sealed and centrifuged (4 °C, 3000 rpm, 30 min). A total of 175 µL of supernatant was transferred to a new plate and evaporated under a nitrogen stream at 40 °C for 30 min. The dried residue was reconstituted in 100 µL of methanol/water (1:9, v/v), followed by centrifugation (4 °C, 3000 rpm, 10 min). Finally, 80 µL of each sample was transferred to a 96-well plate, and 5 µL was injected into the LC-MS/MS system.

Tryptophan and its downstream metabolites, including kynurenine pathway and indole derivatives, were quantified using a targeted LC-MS/MS method following methanol extraction and addition of isotope-labeled internal standards. Serum samples, calibration standards, and quality controls were spiked with internal standards, derivatized with ethyl bromoacetate, quenched, centrifuged, and analyzed by UPLC-MS/MS on a HILIC column with positive ESI in MRM mode. Quantification relied on metabolite-to–internal standard peak area ratios and external calibration curves covering the validated LLOQ–ULOQ ranges. Batch accuracy and precision were monitored using quality control samples, and concentrations were reported as absolute values after correction for recovery and instrumental variability.

**Sequential Association Analysis**

For associations identified in the correlation analysis and surviving BH-FDR correction, a three-step sequential association model was applied to assess whether a third variable's change pattern was consistent with an intermediate step between the exposure and the outcome; because the intermediate and outcome variables were both changes computed over the same PRE-to-POST interval, this framework cannot establish temporal precedence (see Limitations). Association 1 (exposure→intermediate), association 2 (intermediate→outcome), and the residual association were each estimated by Spearman ρ. Indirect-pathway estimates were quantified by bootstrap resampling (B=5,000), and the pattern was considered consistent with an indirect pathway when both association 1 and association 2 were statistically significant and the bootstrapped 95% confidence interval of the indirect estimate excluded zero.

**Sensitivity Analyses**

To assess the robustness of significant associations, covariate adjustment was performed using partial Spearman correlation, whereby the covariate was regressed out of both variables in rank space (ordinary least squares on rank-transformed values) prior to computing Spearman ρ on the residuals. Permutation p-values (5,000 resamples) and bootstrap 95% confidence intervals (B=5,000) were obtained on the residual ranks. Pre-specified sensitivity analyses were conducted sequentially for clinically relevant covariates including valvular calcification burden (aortic valve calcium score), peri-procedural hematological changes (hemoglobin decline), concomitant cardiovascular medications (statin, ACEI, and aspirin, individually and jointly), and demographic variables (BMI, age, and sex, individually and jointly). Stratified analyses by hemoglobin decline and sex were also performed.

A falsification analysis was performed by correlating the exposure variable of interest with non-mechanistically related peri-procedural outcomes using the same Spearman framework, to verify that the signal is not attributable to a non-specific procedural stress response.

**Metagenomics detailed methods**

**DNA extraction and sample preparation**

Total DNA was extracted using the ZymoBIOMICS DNA MagBead Kit (Zymo Research, D4311) on a KingFisher Apex automated platform (Thermo Fisher Scientific), following the manufacturer’s instructions.

For tissue samples, approximately 15 mg of material was transferred into ZR BashingBead Lysis Tubes (2.0 mm) containing 750 µL DNA/RNA Shield. For fecal samples, 100 mg of material was processed using ZR BashingBead Lysis Tubes (0.1 and 0.5 mm) with 750 µL DNA/RNA Shield.

Samples were homogenized using a FastPrep-96 instrument (MP Biomedicals) with five bead-beating cycles (1 min at 6.5 m/s, followed by 5 min rest).

**DNA extraction and quality control**

After homogenization, 200 µL of lysate was transferred to a 96-deepwell plate for automated extraction. DNA quantity and quality were assessed using a Qubit 4.0 fluorometer (dsDNA HS assay), FEMTO Pulse system (Agilent), and Denovix DS-11 spectrophotometer.

**Library preparation and sequencing**

Full-length 16S rRNA gene libraries (V1–V9 regions) were prepared using the PacBio Kinnex workflow (PacBio, Rev05, Jan 2025). Amplification was performed using universal primers targeting the 27F and 1492R regions with sample-specific barcodes.

Library quality and fragment size (~18 kb) were assessed using Qubit and Fragment Analyzer systems. SMRTbell libraries were prepared according to PacBio protocols and sequenced on the PacBio Revio platform using 30-hour movie acquisition and adaptive loading at 160 pM.

**Controls**

Positive controls included commercially available microbial community standards (ZymoBIOMICS DNA standards and ATCC MSA-3001). Negative controls included extraction blanks and PCR no-template controls to monitor contamination.

**16S rRNA Amplicon Sequencing, Diversity Analysis, and Differential Abundance**

Genomic DNA was extracted from one to two fecal pellets collected at defined time points using the QIAamp PowerFecal Pro DNA Kit (Qiagen, Cat. No. 51804) according to the manufacturer’s instructions. DNA concentration was quantified with a Qubit fluorometer. Prior to amplification of the bacterial 16S rRNA gene with barcoded primers, DNA quantity, integrity, and purity were evaluated using a Qubit 4.0 fluorometer (Qubit dsDNA HS Assay Kit, Q32851, Thermo Fisher Scientific), a FEMTO Pulse system (Genomic DNA 165 kb Kit, FP-1002-0275, Agilent), and a DeNovix DS-11 UV-Vis spectrophotometer, respectively. Thereafter, Kinnex libraries from full-length V1-V9 16S rRNA amplicons were prepared exactly as outlined in manufacturer documentation: Preparing Kinnex libraries from 16S rRNA amplicons (PacBio, 103-238-800 REV05, Jan2025).

For the amplification of bacterial full-length 16S rRNA genes the following primers were used: Kinnex16S_Fwd CTACACGACGCTCTTCCGATCT/barcode/AGRGTTYGATYMTGGCTCAG (27F primer), Kinnex16S_Rev AAGCAGTGGTATCAACGCAGAG/barcode/RGYTACCTTGTTACGACTT (1492R primer). Barcode is 10nt.

Sequencing was performed on the PacBio Revio system. High-fidelity reads underwent quality filtering and denoising, with amplicon sequence variant inference performed using DADA2 within the QIIME2 framework.(5,6) Taxonomic assignment was performed against the SILVA reference database using QIIME2-compatible classifiers.

Microbial community analyses were performed using established microbiome analysis packages, including phyloseq, vegan, and microbiome.(7,8) Alpha diversity was assessed using richness and diversity metrics including observed amplicon sequence variants, Chao1 richness, Shannon entropy, and Simpson index. Beta diversity was assessed using Bray-Curtis dissimilarity and visualized by principal coordinate analysis. Pre-to-post procedural changes in microbial community composition were tested using PERMANOVA, with paired analyses stratified by patient as the primary approach. Complete paired microbiome datasets were available for 38 patients.

Differential abundance analysis was performed using MaAsLin3, applying multivariable linear modeling to identify taxa associated with pre-to-post TAVI status and relevant clinical variables.(9) Multiple testing was addressed using Benjamini-Hochberg false discovery rate correction. Functional guild analyses based on genus-level profiles were performed in 33 patients with complete paired data available for these analyses.

**Functional Guild Inference**

Unlike computational metagenome-prediction tools such as PICRUSt2, which infer whole predicted gene content from 16S phylogenetic placement (18), guild scores here were derived from a pre-specified, literature-curated genus-to-function assignment matrix, restricted to genera with direct genomic or biochemical evidence for the relevant pathway, conceptually analogous to curated metabolic module frameworks previously applied to the human gut microbiome.(10)

To translate 16S taxonomic profiles into metabolic functional capacity, genus-level relative abundances were mapped onto a pre-specified binary genus-function matrix. Each genus was assigned a binary score (1/0) for each of seven functional guilds based on genomic and biochemical evidence from the literature, independent of the study data. Guild scores were computed as the sum of relative abundances of all genera assigned to each guild, yielding a continuous score per sample reflecting the aggregate functional potential of the community.

Three guilds were selected a priori as primary axes of interest based on their mechanistic relevance to the SCFA–inflammation hypothesis. The True Butyrate Synthesis guild was restricted to genera strictly possessing the butyryl-CoA:acetate CoA-transferase terminal enzyme (including *Faecalibacterium, Roseburia, Coprococcus, Agathobacter, Anaerobutyricum*, and *Butyricicoccus*). Genera with general saccharolytic capacity but lacking this terminal enzyme (including Bacteroides, which produces acetate and propionate via distinct pathways, and Blautia, an acetogen via the Wood-Ljungdahl pathway) were explicitly excluded, as supported by genomic mapping of the butyryl-CoA:acetate CoA-transferase gene across human gut commensals.(11,12) The Indole Synthesis Potential guild was mapped to tryptophanase (tnaA)-positive genera responsible for protective tryptophan catabolism into indole derivatives.(13,14) The Proteolytic Degradation guild was restricted to facultative anaerobes and pathobionts equipped with extensive extracellular protease machinery (Enterococcus, Escherichia, Fusobacterium, Anaerococcus, Campylobacter), which utilize amino acid putrefaction as an alternative energy source, yielding branched-chain fatty acids and pro-inflammatory lipopolysaccharide.(15–17) Additional guilds (Saccharolytic, Mucin_degrader, Barrier_support, Pathobiont) were used for exploratory analyses.

**Cytokine panel assessment**

Serum samples were diluted 1:2-1:10 in MSD Diluent Buffer, depending on the assay panel, and added to V-PLEX plates together with serially diluted cytokine standards for calibration. Plates were incubated for 2 h at room temperature with shaking, followed by three washes with PBS containing 0.05% Tween-20. A detection antibody solution containing the commercial MSD SULFO-TAG secondary antibodies, diluted 1:50 in Diluent Buffer, was then added to each well and incubated for 1 h at room temperature with shaking. Following three additional washes with PBS containing 0.05% Tween-20, 150 μL of MSD Read Buffer was added to each well, and electrochemiluminescence signals were acquired using the MESO QuickPlex SQ 120 instrument (Meso Scale Discovery). Cytokine concentrations were calculated from assay-specific standard curves using Discovery Workbench software (Meso Scale Discovery). Assay performance was monitored using standard platform-specific quality-control procedures, including assessment of calibration performance, lower limit of quantification behavior, and intra- and inter-assay variability. Eight cytokines were measured using a multiplex immunoassay (MSD 8-plex). IL-13 was excluded from all analyses because concentrations were below the lower limit of quantification in 72.9% of samples, yielding seven cytokines for statistical analysis. The global SCFA × cytokine cross-correlation screen comprised 35 tests (5 SCFAs × 7 cytokines; Figure 4A); BH-FDR correction was applied within the butyrate row only (7 tests) for the cytokine specificity analysis (Figure 4C).

Adverse events

An inflammatory composite was defined at each timepoint: periprocedural (new permanent pacemaker implantation [PPM] + early structural valve deterioration [SVD]); 1-month (persistent left bundle branch block [LBBB] at 30-day follow-up + first-degree atrioventricular block [AVB1] + new-onset atrial fibrillation + SVD on Days 1–35); 3-month (complete AV block [AVB3] or any hemodynamic leaflet thrombosis [HALT, REDCap codes 3, 6, 7]); 1-year (new PPM since last contact + LBBB or right BBB [RBBB] present at 1-year + AVB1 present at 1-year + new-onset AF).

SVD at 1-month was classified as an inflammatory event on the basis that early SVD within 35 days of TAVI predominantly represents a thrombotic-inflammatory process rather than structural leaflet failure. A pre-specified sensitivity analysis tested the 1-month composite excluding SVD.

For comparisons between SCFA Responders and Non-Responders, Fisher’s exact test (two-sided) was used. When any cell of the 2×2 table contained zero observations, odds ratios and 95% confidence intervals were computed with Haldane–Anscombe continuity correction (+0.5 to all cells); p-values were taken from the uncorrected exact test. No multiplicity correction was applied to adverse event comparisons, which were pre-specified as exploratory. One-year analyses were restricted to patients with available 1-year follow-up.

Major adverse events were prospectively adjudicated by an independent Clinical Events Committee according to the Valve Academic Research Consortium-3 (VARC-3) criteria (18) within the SwissTAVI registry. Adjudicated endpoints included cardiovascular death, stroke or transient ischemic attack, myocardial infarction, structural valve deterioration, vascular access-site complications, VARC bleeding, and repeat unplanned intervention.

Conduction abnormalities and arrhythmias were ascertained from systematic ECG assessments performed at each scheduled follow-up visit (discharge, 30 days, 1 year). New permanent pacemaker implantation was recorded as a clinician-reported outcome at each visit. Bundle branch block (LBBB/RBBB) and atrioventricular block (degree recorded per standard ECG classification) were extracted from the 'Intraventricular conduction delay' and 'Degree' fields respectively. New-onset atrial fibrillation was captured via a dedicated registry variable ('New-Onset of Atrial Fibrillation since baseline') present at each visit. Conduction events were only attributed to a given timepoint if present on the ECG recorded at that visit.
